# Evaluating synthetic-data fidelity in two-group biomedical studies: a multidimensional validation framework

**DOI:** 10.64898/2026.08.31.26361878

**Authors:** Tony Tran, Mohammad Sajjad Ghaemi, Chapin S. Korosec

## Abstract

Synthetic data increasingly support model development and privacy-conscious sharing in biomedicine. Two-group studies require synthetic data to reproduce within-group structure and between-group differences, yet marginal agreement or predictive performance may obscure multivariate and conditional-dependence changes. We present a multidimensional validation framework for class-conditional synthetic data and apply it to three datasets spanning sample-size and dimensionality regimes. Two controls and four generators spanning mixture, interpolation, hybrid, and latent-variable architectures (GMM, SMOTE, GMM-SMOTE, and CVAE, respectively) were assessed using predictive utility, real–synthetic distinguishability, marginal agreement, PCA and t-SNE geometry, pairwise dependence, and Graphical LASSO networks. Noise perturbation, within-class permutation, and reverse ablation probed the sources of real–synthetic distinguishability. Across 18 dataset–method comparisons, discriminator AUC ranged from 0.55 to 1.00, while mean feature-level KS statistics ranged from 0.027 to 0.282. Thus, strong performance under individual criteria coexisted with detectable differences and lost or synthetic-only dependencies. Rather than assigning a single fidelity score, the framework supports multidimensional fidelity reporting as a minimum standard for shared synthetic biomedical data.

## 1 Introduction

Synthetic data are rapidly becoming integral to biomedical and clinical research (*1*), enabling privacy-preserving data sharing (*2–4*), dataset augmentation (*5*), AI model development and validation (*6–8*), and the construction or augmentation of data-derived control arms for clinical studies (*9–11*). The broader demand for high-quality, AI-ready biomedical data is reflected in NIH Common Fund initiatives: Bridge2AI (*12*) supports FAIR, interoperable, AI-ready biomedical and behavioral datasets, while PRIMED-AI (*13*) integrates multimodal clinical data and emphasizes rigorous, generalizable model validation. Beyond improving data access and sample size, synthetic data support scientific inference (*14*) and the development and validation of mechanistic and computational modeling frameworks; when generated from known mechanisms and parameters, they provide ground truth for assessing parameter recovery, identifiability, and model-selection performance (*15–17*). Although clinical applications remain emergent, synthetic datasets are also being investigated for the development and evaluation of clinical decision-support systems (*18, 19*). Across these applications, fidelity is inherently multidimensional and application-dependent: synthetic data may reproduce marginal distributions while altering global data structure, preserving some feature dependencies while creating or destroying others. A generalizable, multidimensional fidelity-evaluation framework that jointly assesses statistical agreement, global data structure, and feature dependencies preserved, lost, or synthetic-only during generation would therefore represent a significant advance toward the trustworthy, fit-for-purpose use of synthetic data in biomedical discovery, AI-enabled research, and regulatory science.

Before synthetic data can be responsibly used in downstream applications, fidelity must be evaluated against the properties required for the intended use. However, fidelity has no single standardized operational definition, and available measures interrogate different, and potentially discordant, dimensions of similarity (*19–21*). Targeted approaches assess specific dimensions through propensity- or classifier-based real–synthetic distinguishability (*22, 23*), replication of selected statistical analyses (*24, 25*), downstream predictive performance (*26*), and sample-level fidelity, coverage, and authenticity (*27*). Distributional fidelity has similarly been quantified using feature-level chi-squared, Kolmogorov–Smirnov, and marginal divergence measures (*7*), as well as global multivariate measures such as multivariate Hellinger distance (*28*) and classifier-estimated Kullback–Leibler and Jensen–Shannon divergences between joint real and synthetic distributions (*29*). Broader evaluations combine marginal, pairwise, population-level, predictive, and privacy criteria (*30–32*), while frameworks such as SynthEval integrate many of these measures within configurable evaluation pipelines (*33, 34*). Healthcare-specific frameworks have further incorporated statistical quality, privacy, machine-learning usability, computational complexity, and checks for duplicated, outlying, or out-of-range observations (*35*). Consistent with this multidimensional view, a recent benchmark of 11 generators across nine healthcare datasets found that no single model performed best across fidelity, utility, and privacy, further demonstrating that assessment outcomes depend on both the source dataset and the evaluation criterion (*36*). More recently, TabStruct formalized structural fidelity as a distinct evaluation dimension, assessing conditional-independence preservation when expert-validated causal structures are available and introducing a global-utility measure for real-world settings in which ground-truth causal structure is unavailable (*37*). TabStruct also highlights that causal-discovery-based comparisons are difficult to validate when the inferred graphs cannot be compared with a known ground truth. Collectively, these studies demonstrate that fidelity is criterion-dependent: strong performance under one measure does not guarantee strong performance under another. However, existing multidimensional frameworks typically summarize structural fidelity globally or evaluate conditional-independence preservation when causal structure is known, rather than localizing which conditional-dependence relationships are preserved, lost, or synthetic-only in observational biomedical data. Fit-for-purpose validation therefore requires complementary measures that resolve these relationship-specific structural changes without presuming a ground-truth causal model.

The structural and statistical properties of multivariate synthetic data are shaped by the assumptions of the generating model. Because generator families represent and sample the joint distribution through fundamentally different architectures (*19, 21*), they may preserve, attenuate, eliminate, or create distinct multivariate relationships. For example, univariate synthesis can reproduce marginal distributions without preserving joint or conditional structure (*38, 39*).Recent dependency-aware synthesis work has similarly shown that functional and logical inter-attribute dependencies can be poorly retained across generator families, and that explicitly reconstructing these relationships can improve both structural fidelity and downstream utility (*40*). Independent feature resampling exemplifies this limitation by breaking observation-level alignment and systematic cross-feature dependence. Finite Gaussian mixture models (GMMs) approximate a joint density through weighted multivariate Gaussian components whose fitted means and covariance matrices determine the locations and geometry of the modeled regions (*41, 42*). Because GMM approximation depends on the number of components and covariance constraints, fitted mixtures may merge or subdivide non-Gaussian structure and alter the dispersion or orientation of generated observations (*43, 44*). Interpolation-based methods generate observations along line segments between sampled observations and their neighbors (*45*); because these interpolants are drawn inward relative to the source distribution, they may contract its support or populate low-density regions between observations (*46, 47*). Latent-variable models encode observations into a learned latent representation and generate samples through a decoder (*48, 49*); the resulting compression– reconstruction trade-off and potential loss of informative latent dimensions may attenuate variation and alter the within and between-group feature-dependence structure represented in the synthetic data (*50, 51*). Taken together, these generator-specific distortions expose a gap in existing multidimensional assessments: marginal agreement, low-dimensional overlap, pairwise association, and global distinguishability characterize complementary dimensions of fidelity without resolving which conditional-dependence relationships have been preserved, lost, or synthetic-only.

Here, we introduce a multidimensional fidelity framework tailored to class-conditional synthetic data generated from two-group biomedical and clinical studies. We apply the framework to three openly available biomedical datasets spanning contrasting sample-size and dimensionality regimes, comparing two sampling-based controls with four synthetic-data generators representing mixture, interpolation, hybrid, and latent-variable architectures while matching sample size and class composition to the source data. The framework jointly evaluates downstream predictive utility, held-out real–synthetic distinguishability, marginal distributional agreement, PCA, t-SNE-based multivariate geometry, pairwise covariance and correlation, and sparse conditional-dependence structure estimated using Graphical Lasso (*52*). Our central advance is the integration of these complementary diagnostics with an explicit network-level classification of feature dependencies as preserved, lost, or synthetic-only. Noise perturbation, progressive within-class permutation, and reverse ablation further determine whether real–synthetic distinguishability arises from insufficient dispersion, disrupted feature dependence, or a limited subset of features. Across datasets and generators, we show that strong marginal agreement or predictive utility can coexist with high distinguishability and with structural distortions in the synthetic data, including the loss of conditional dependencies present in the source data and the creation of synthetic-only dependencies absent from it. The purpose of the framework is therefore not to certify synthetic data as universally high quality, rather to determine the degree and dimensions of fidelity achieved, and consequently the level of trust warranted for a specified use, supporting transparent, fit-for-purpose decisions about the analysis, reuse, and sharing of synthetic biomedical data.

## 2 Methods

We evaluated synthetic-data fidelity using three open biomedical binary-classification datasets deliberately selected to span contrasting sample-size and dimensionality regimes (Table 1). For each dataset, multiple synthetic-data generators were assessed using complementary measures of predictive utility, real–synthetic distinguishability, marginal fidelity, low-dimensional geometry, and conditional-dependence structure. This framework was designed to determine not only whether synthetic data reproduced individual variables or broad statistical patterns, but also whether relationships between biomarkers were preserved, lost, or present only in the synthetic data. A schematic illustrating the full process of input, synthetic data generation, and fidelity assessment is shown in Figure 1.

**Figure 1:**
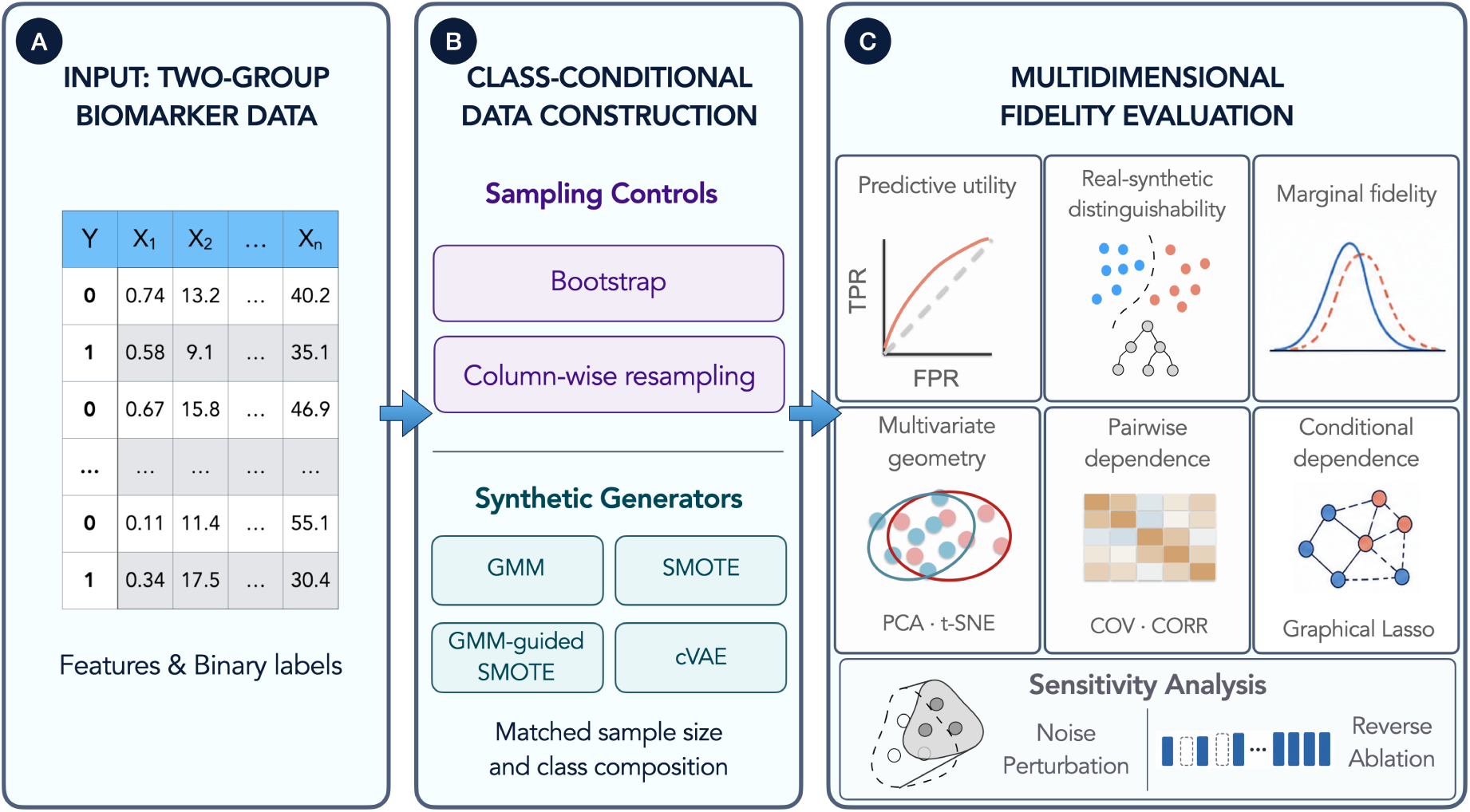
Multidimensional synthetic-data fidelity assessment. (A) Real biomarker features and binary outcome labels serve as the input data. (B) Data are constructed separately within each outcome class using two sampling controls and four synthetic-data generators while preserving sample size and class composition. (C) Fidelity is evaluated across predictive utility, real–synthetic distinguishability, marginal distributions, multivariate geometry, pairwise and conditional dependence, with robustness examined through noise perturbation, reverse ablation, and within-class permutation.

**Table 1:** Binary classification datasets used for synthetic-data evaluation. *n* denotes the number of observations, and *n*_0_ and *n*_1_ denote the class counts. All analyzed features were continuous.

| **Dataset** | **n** | **n<sub>0</sub>** | **n<sub>1</sub>** | **Features, $p$** | **Source** |
| --- | --- | --- | --- | --- | --- |
| HIV cohort | 91 | 23 | 68 | 63 | Korosec et al. (53) |
| Breast cancer | 569 | 212 | 357 | 30 | Street et al. (55) |
| Diabetes | 768 | 500 | 268 | 8 | Smith et al. (57) |

### 2.1 Datasets and preprocessing

Three openly accessible biomedical datasets were selected to represent contrasting sample-size and dimensionality settings while retaining a common binary-classification structure (Table 1).

The HIV vaccine-response dataset was obtained from the public data release accompanying Korosec et al. (*53*) which is an extended dataset originally published in Matveev et al. (*54*). It contains 91 participants who received up to 5 SARS-CoV-2 vaccinations, comprising 23 HIV-negative controls and 68 people living with HIV receiving antiretroviral therapy. Participants were represented by 63 longitudinal immunological features derived from repeated SARS-CoV-2 vaccination, including serum and saliva antibody responses, cytokine-producing T-cell responses, the CD4/CD8 ratio, viral neutralization, and ACE2-displacement measurements. The binary outcome distinguished HIV-negative participants from participants living with HIV.

The Wisconsin Diagnostic Breast Cancer dataset was originally developed from digitized images of fine-needle aspirates of breast masses (*55*) and was accessed using the load breast cancer function from the Python scikit-learn library (*56*). The dataset contains 569 observations and 30 quantitative features describing properties of cell nuclei, including their size, texture, shape, smoothness, and concavity. The binary outcome identifies malignant and benign diagnoses.

The Pima Indians Diabetes dataset contains 768 observations and eight clinical features, including measures of glucose concentration, blood pressure, body mass index, insulin, age, and diabetes pedigree (*57*). The dataset was accessed from the Penn Machine Learning Benchmarks repository using the Python pmlb library (*58*). Its binary outcome indicates whether an individual met the diagnostic criteria for diabetes.

Together, the datasets provided a deliberately varied evaluation setting: a small, high-dimensional immunological dataset; an intermediate-sized dataset with moderate dimensionality; and a larger dataset with comparatively few features. Synthetic datasets are generated to reproduce the number of observations and binary class composition of the corresponding real datasets. When an evaluation required features on a common scale, preprocessing parameters were estimated from the real dataset and then applied unchanged to the corresponding synthetic datasets. Model-specific transformations used during synthetic-data generation are described in Section 2.2.

### 2.2 Synthetic-data generation

Generator selection was designed to span distinct data-construction mechanisms under the small- to moderate-sample regimes represented by the three datasets, rather than to exhaustively benchmark contemporary tabular synthesizers. GAN- and diffusion-based generators, including CTGAN, CTAB-GAN+, and TabDDPM, were not evaluated because their inclusion would require a broader neural-generator benchmarking design with architecture-specific optimization, particularly for the small-(n), high-(p) data sets used in this work (*59*). We therefore prioritized two sampling controls and four generators representing mixture, interpolation, hybrid, and conditional latent-variable approaches that could be applied consistently across datasets. The generators represented distinct mechanisms for constructing novel tabular observations, whereas the controls were included to establish interpretable reference cases for the preservation or disruption of the empirical data structure. To support reproducibility and comparison across datasets, each procedure used a fixed, prespecified configuration rather than being exhaustively tuned for each dataset. Observations were produced separately by outcome class until the corresponding real-data class count was reproduced. The nonparametric bootstrap was included as a sampling-based control rather than as a synthetic-data-generation technique (*60*). Complete observations were sampled with replacement within each outcome class. Because all features belonging to an observation were resampled together, the bootstrap retained the empirical joint structure and cross-feature relationships present in the real data, subject to variation arising from finite resampling. It therefore provided a reference for performance when no new feature combinations were generated and the observed multivariate structure was preserved.

Column-wise random resampling was included as a second control designed to represent the opposite structural condition. Within each outcome class, every feature was sampled independently with replacement from its empirical distribution. This procedure preserved the class-conditional univariate marginal distribution of each feature while deliberately breaking the observed alignment, covariance, and interdependence among features. Comparison with this control therefore helped determine whether an evaluation metric was sensitive to multivariate structure rather than only to marginal fidelity.

Gaussian mixture models (GMMs) were fitted separately within each outcome class using expectation–maximization (*61*). For the primary analysis, each class-specific model used two components, full covariance matrices, covariance regularization of (10^−4^), and a maximum of 100 fitting iterations. When necessary, the number of components was reduced so that each fitted component was supported by at least two observations. Synthetic observations were sampled directly from the fitted mixtures to reproduce the corresponding real-data class counts. To assess sensitivity to the fixed two-component specification, candidate models containing (K=2,. . . ,5) components were also fitted separately within each class using otherwise identical settings. The component number was selected independently for each class by minimizing either the Akaike information criterion (AIC) or Bayesian information criterion (BIC), producing separate AIC- and BIC-selected synthetic datasets. The selected component numbers are reported in Supplementary Table S5.

SMOTE datasets were generated separately within each outcome class (*45*). For each synthetic observation, an anchor observation was sampled, one of its five nearest within-class neighbours was selected, and a point was generated at a uniformly sampled position along the line segment connecting the anchor and neighbour. Nearest-neighbour distances and interpolation were calculated in the original feature space.

GMM-guided SMOTE combined GMM-based partitioning with local interpolation, motivated by the proposed hybrid of GMM and SMOTE for preserving complementary global and local characteristics (*53*). A three-component GMM with full covariance matrices and covariance regularization of 10^−4^ was fitted separately within each outcome class. Each real observation was assigned to its most likely mixture component. For each synthetic observation, a real anchor was sampled, and one of its five nearest within-class neighbours assigned to the same component was selected. When the anchor’s component contained no other observations, neighbour selection was performed across the full outcome class. The synthetic observation was generated as

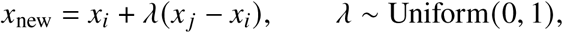

where *x_i_* and *x _j_* denote the real anchor and selected neighbour, respectively. Nearest-neighbour distances and interpolation were calculated in the original feature space.

The conditional variational autoencoder (CVAE) followed the variational latent-variable framework of (*48*) and its conditional extension by (*62*). The encoder and decoder each contained two fully connected hidden layers of 128 units with rectified linear-unit activations, and the latent dimension was 16. The objective combined squared reconstruction error with a KL-divergence term weighted by *β* = 0.5. The model was trained for 200 epochs using the Adam optimizer (*63*), a learning rate of 10^−3^, and a batch size of 64. A stratified 20% validation partition was held out, and the model parameters associated with the lowest validation loss were retained.

For CVAE training, the features were standardized using statistics estimated from the training partition, and outcome classes were represented by one-hot condition vectors. During generation, latent vectors were sampled from a 16-dimensional standard multivariate normal distribution and decoded conditional on the requested outcome class. Generated observations were then inverse- standardized to the original feature scales.

All stochastic procedures used deterministic seeds derived from master seed 42.

### 2.3 Synthetic-data evaluation

Each synthetic dataset was evaluated relative to its corresponding real dataset using the complementary analyses described in the following subsections. Downstream predictive utility was assessed using supervised classification across real and synthetic training and testing conditions. Real– synthetic distinguishability was evaluated by training classifiers to predict whether observations originated from the real or synthetic dataset, with feature-importance analyses used to identify the variables contributing to this discrimination. Feature-wise comparisons of marginal distributions and summary statistics were used to assess univariate fidelity, while principal component analysis (PCA), and *t*-distributed stochastic neighbour embedding (*t*-SNE), were used to compare low-dimensional representations of the multivariate data. Finally, covariance and correlation matrices were compared to assess pairwise feature relationships, and Graphical Lasso was used to evaluate preservation of sparse conditional-dependence structure. These analyses were applied separately to each synthetic dataset.

#### 2.3.1 Real–synthetic distinguishability

For each synthetic dataset, the real and synthetic observations were combined and assigned binary source labels, with 0 denoting real observations and 1 denoting synthetic observations. The real and synthetic samples were equal in size and had identical outcome-class compositions. The original outcome label was excluded from the discriminator predictors and was used only to preserve class composition during partitioning.

The combined data were divided into 75% training and 25% testing partitions, stratified jointly by source and outcome class. A random-forest discriminator (*64*) containing 500 trees was trained to predict whether each observation was real or synthetic. Discrimination was evaluated on the heldout observations using the area under the receiver operating characteristic curve (ROC AUC) (*65*), calculated from the predicted probability that an observation was synthetic. An AUC of 0.5 indicated chance-level discrimination, whereas values approaching 1 indicated increasingly systematic differences between the real and synthetic observations. This procedure was repeated across 50 independently sampled stratified train–test partitions to characterize variability attributable to data partitioning.

Feature contributions to real–synthetic discrimination were quantified using permutation importance on the held-out test observations, measured as the decrease in ROC AUC produced by independently permuting each feature. Importance values were averaged across the 50 partitions to obtain an aggregate feature ranking. For reverse-ablation analyses, feature rankings were estimated using the training data within each repetition, and discrimination following ablation was evaluated using the corresponding held-out test data. A separate 500-tree discriminator was fitted to the complete combined dataset using random seed 42 solely to obtain the descriptive feature ranking used in the network-overlay analysis.

#### 2.3.2 Noise-sensitivity analysis

To test whether the concentration of synthetic observations accounted for real–synthetic distinguishability, independent Gaussian noise was added as

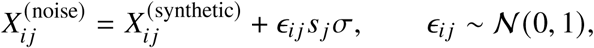

where *s _j_* was the corresponding real-feature standard deviation and *σ* controlled the noise magnitude. The discriminator was reevaluated at *σ* = 0 to 2, with five repetitions per level.

#### 2.3.3 Predictive utility

Predictive utility was evaluated using train-on-synthetic, test-on-real (TSTR) and train-on-real, test-on-real (TRTR) protocols. For TSTR, a 300-tree random-forest classifier (*64*) with balanced class weights was trained on the complete synthetic dataset and evaluated on the corresponding real dataset. Performance was measured using the positive-class F1 score.

TRTR performance was estimated by stratified cross-validation on the real dataset, using up to five folds according to the size of the minority class. Both TSTR and TRTR were repeated 20 times, with repetition-specific random seed 42 + 1009*r* for repetition *r*. The paired utility gap was defined as

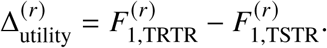

#### 2.3.4 Marginal fidelity

Marginal agreement was evaluated separately for each continuous feature. Feature means were compared using two-sided Welch unequal-variance *t*-tests (*66*), and complete empirical distributions were compared using the two-sample Kolmogorov–Smirnov statistic (*67*).

Distributional discrepancy was additionally measured using Kullback–Leibler divergence (*68*). For each feature, real and synthetic values were divided into 30 aligned histogram bins spanning their pooled range. A smoothing constant of 10^−10^ was added to each bin before normalization. Dataset-level KLD was summarized as the mean feature-level divergence. Feature-level statistics and distribution plots are reported in the Supplementary Tables S3, S4, S2, S1.

#### 2.3.5 Multivariate geometry

Principal component analysis (*69*) was fitted to the standardized real feature matrix for each dataset. Synthetic observations were standardized using the corresponding real-data parameters and projected into the fitted real-data component space using the same component loadings.

Real and synthetic PCA scores were summarized using covariance ellipses centred at their respective sample means. Under an approximate bivariate-normal assumption, each ellipse encloses a region expected to contain 95% of the observations. The ellipses summarize the location, spread, and orientation of the PCA scores and should not be interpreted as confidence intervals for the mean.

#### 2.3.6 Pairwise correlation structure

Pairwise linear dependence was evaluated by calculating the Pearson correlation coefficient, *r*, between every pair of features in each real and synthetic dataset. For each generation method, the resulting synthetic correlation matrix, *R_S_*, was compared with the corresponding real-data correlation matrix, *R_R_*. Feature ordering was held constant across the real and synthetic matrices.

Overall disagreement was quantified using the off-diagonal Frobenius distance,

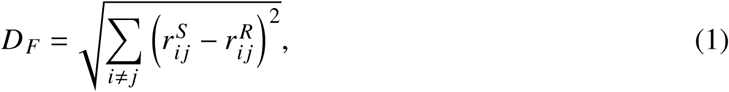

where 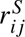 and 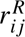 denote the synthetic- and real-data correlations, respectively. Diagonal elements were excluded because they are equal to one by definition. Smaller values indicate closer preservation of the real-data pairwise correlation structure.

Because the number of feature pairs differed among datasets, a dimension-normalized correlation error was also calculated as

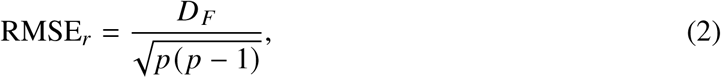

where *p* is the number of features. This quantity represents the root-mean-square difference across all off-diagonal correlation coefficients and permits comparison of correlation error across datasets with different numbers of features.

#### 2.3.7 Feature distribution of real–synthetic distinguishability

Features were ranked from highest to lowest mean-decrease-in-impurity importance in the full-data real–synthetic discriminator. Increasing numbers of top-ranked features were removed, and the discriminator was refitted using the remaining features.

Eight approximately evenly spaced removal levels were evaluated, ending with two features remaining. At each level, test AUC was estimated over 20 repeated stratified train–test splits. The feature order was fixed throughout the analysis.

#### 2.3.8 Conditional-dependence structure

Graphical Lasso (*52*) was used to compare conditional relationships in the real and synthetic data. Models were fitted separately to the standardized real and synthetic observations. For each dataset, the regularization parameter, *λ*, was selected from the real data using scikit-learn’s GraphicalLassoCV implementation with five-fold cross-validation, eight candidate values at each search stage, and three grid refinements. The value maximizing the mean cross-validated Gaussian log-likelihood was retained. This procedure selected *λ* = 0.504 for HIV, *λ* = 0.502 for breast cancer, and *λ* = 0.0159 for diabetes. The selected value was then reused for every corresponding synthetic dataset, keeping the level of regularization consistent across comparisons.

Each model was fitted for up to 1000 iterations, and the resulting precision matrix was converted to a partial-correlation network. Nonzero off-diagonal entries were treated as edges and classified as preserved, lost, or synthetic-only according to whether they appeared in the real network, the synthetic network, or both.

For visualization, the feature order was fixed within each dataset using the real-data partial-correlation matrix. Average-linkage hierarchical clustering was applied with

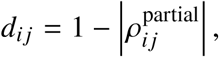

and the resulting order was reused for all corresponding synthetic networks. This ensured that differences between panels reflected changes in edge status rather than changes in feature arrangement. Feature names and plotted positions are provided in Supplementary Table S7.

Structural agreement was quantified using three complementary measures: the proportion of real edges recovered in the synthetic network, the proportion of synthetic edges absent from the real network, and the off-diagonal Frobenius distance between the real and synthetic precision matrices.

#### 2.3.9 Within-class permutation analysis

To test what happens when feature relationships are disrupted but individual feature distributions remain unchanged, we progressively shuffled values within each feature and outcome class. At each level, between 10% and 100% of the values in every feature were selected and permuted among observations without replacement, in increments of 10 percentage points. Each feature therefore retained exactly the same values within each class, while the combinations of values across features were gradually broken apart.

At each permutation level, the random-forest discriminator was used to distinguish the real observations from their permuted counterparts. Graphical Lasso was also refitted using the regularization parameter selected from the real data. Real-edge retention was calculated as the proportion of edges in the real network that also appeared in the permuted network:

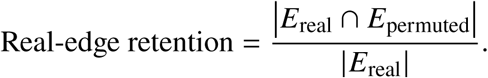

The analysis was repeated 10 times at each permutation level.

## 3 Results

Synthetic-data fidelity was evaluated across the HIV, breast cancer, and diabetes datasets, which span contrasting sample-size and dimensionality regimes. The same class-conditional generation and validation pipeline was applied to each dataset, incorporating real–synthetic distinguishability, predictive utility, marginal-distribution agreement, multivariate geometry, reverse feature ablation, and conditional-dependence structure. This common evaluation design allowed us to examine how the dimensions of fidelity achieved varied with both the characteristics of the source data and the synthetic-data generator.

### Real–synthetic AUC and predictive utility across datasets

Mean discriminator AUC, ⟨AUC⟩, quantified how readily held-out real and synthetic observations could be separated by source. An ⟨AUC⟩ near 0.5 indicates chance-level discrimination, meaning that the discriminator cannot reliably distinguish synthetic observations from real observations and therefore suggesting greater fidelity under this criterion. Conversely, values approaching 1.0 indicate increasingly complete real–synthetic separation. Predictive utility was evaluated alongside distinguishability to determine whether synthetic data retained the class-related information required for downstream prediction. Results across all six generation methods are summarized in Table 2 and Figure 2A–C and G–I.

**Figure 2:**
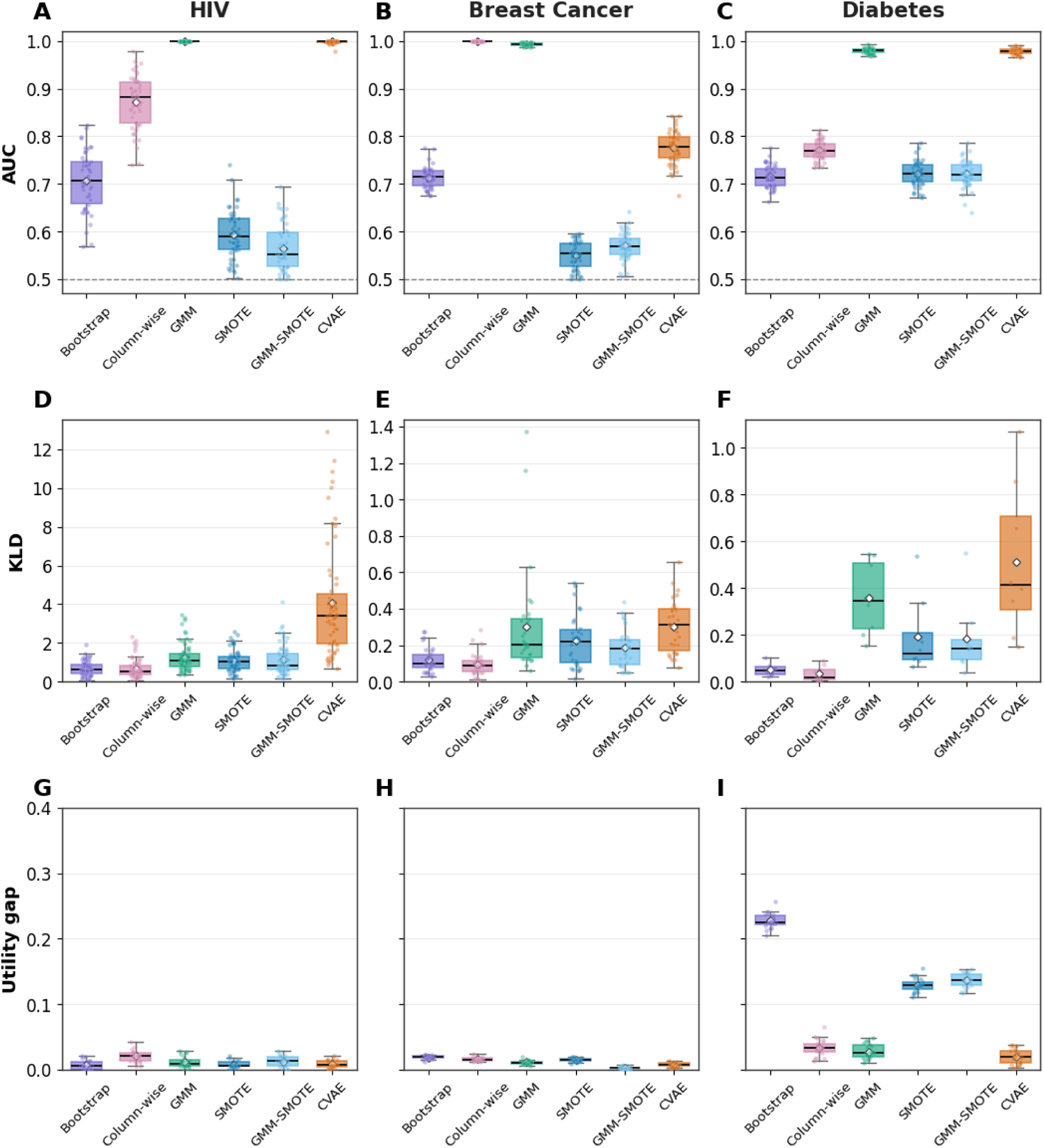
Real–synthetic distinguishability, marginal discrepancy, and predictive utility across datasets and generation methods. Panels A–C show real–synthetic discriminator AUC for the HIV, breast cancer, and diabetes datasets, respectively. Panels D–F show mean feature-level Kullback– Leibler divergence, and panels G–I show the absolute utility gap between train-on-real, test-on-real and train-on-synthetic, test-on-real F1 scores. Boxes represent the interquartile range centre lines indicate medians, diamonds indicate means, and points show individual repetitions. The dashed line in panels A–C denotes chance-level discrimination at AUC = 0.5.

**Table 2:** Main summary of discriminator and utility metrics across datasets and methods. AUC values are means across 50 discriminator repetitions; utility metrics are means across 20 repetitions.

| Dataset | Method | $\langle \text{AUC} \rangle$ | TSTR F1 | $\vert \text{TRTR} - \text{TSTR}\vert $ |
| --- | --- | --- | --- | --- |
| HIV | Bootstrap | 0.71 | 0.97 | 0.01 |
| HIV | Column-wise | 0.86 | 0.99 | 0.02 |
| HIV | GMM | 1.00 | 0.98 | 0.01 |
| HIV | CVAE | 1.00 | 0.98 | 0.01 |
| HIV | SMOTE | 0.59 | 0.97 | 0.01 |
| HIV | GMM-SMOTE | 0.57 | 0.98 | 0.01 |
| Breast Cancer | Bootstrap | 0.71 | 0.99 | 0.02 |
| Breast Cancer | Column-wise | 1.00 | 0.96 | 0.01 |
| Breast Cancer | GMM | 0.99 | 0.96 | 0.01 |
| Breast Cancer | CVAE | 0.82 | 0.97 | 0.01 |
| Breast Cancer | SMOTE | 0.55 | 0.98 | 0.02 |
| Breast Cancer | GMM-SMOTE | 0.57 | 0.97 | 0.00 |
| Diabetes | Bootstrap | 0.72 | 0.86 | 0.23 |
| Diabetes | Column-wise | 0.79 | 0.67 | 0.04 |
| Diabetes | GMM | 0.98 | 0.63 | 0.01 |
| Diabetes | CVAE | 0.99 | 0.63 | 0.01 |
| Diabetes | SMOTE | 0.72 | 0.76 | 0.13 |
| Diabetes | GMM-SMOTE | 0.72 | 0.77 | 0.14 |

For HIV, SMOTE and GMM–SMOTE produced the least distinguishable synthetic observations, with ⟨AUC⟩ values of 0.59 and 0.57, respectively. Bootstrap showed intermediate distinguishability at 0.71, while column-wise resampling increased separation to 0.86. GMM and CVAE samples were almost completely distinguishable from the real observations, with both reaching an ⟨AUC⟩ of 1.00. Despite this broad range in distinguishability, predictive utility remained consistently high: TSTR F1 scores ranged from 0.97 to 0.99, and absolute differences from the TRTR reference ranged from 0.01 to 0.02.

Breast cancer showed a similar separation between source distinguishability and predictive utility. SMOTE and GMM–SMOTE again produced discriminator performance closest to chance, with ⟨AUC⟩ values of 0.55 and 0.57. Bootstrap reached 0.71 and CVAE reached 0.82, whereas column-wise resampling and GMM were nearly completely distinguishable at 1.00 and 0.99, respectively. Nevertheless, all six methods retained high predictive performance, with TSTR F1 scores between 0.96 and 0.99 and absolute utility gaps no greater than 0.02.

The diabetes dataset exhibited a different utility pattern. GMM and CVAE remained highly distinguishable, with ⟨AUC⟩ values of 0.98 and 0.99, while column-wise resampling reached 0.79. Bootstrap, SMOTE, and GMM–SMOTE each produced lower but still appreciable distinguishability, with values of approximately 0.72. TSTR F1 scores varied more widely than in the other datasets: bootstrap achieved 0.86, followed by GMM–SMOTE at 0.77, SMOTE at 0.76, column-wise resampling at 0.67, and GMM and CVAE at 0.63. The larger absolute utility gaps for bootstrap, SMOTE, and GMM–SMOTE reflected TSTR performance above, rather than below, the corresponding TRTR reference.

Together, these results demonstrate that real–synthetic distinguishability and predictive utility interrogate different dimensions of fidelity. Synthetic observations could retain or even strengthen class-predictive information while remaining systematically different from the real data. Conversely, close agreement with TRTR performance did not imply low real–synthetic distinguishability. Predictive utility alone was therefore insufficient to establish whether the broader structure of the source data had been reproduced.

The noise-sensitivity analysis further tested whether insufficient dispersion alone accounted for real–synthetic distinguishability. Adding feature-scaled independent Gaussian noise generally increased discriminator AUC toward 1.0, while GMM and CVAE remained near complete separation across the evaluated noise levels (Supplementary Figure S1). Thus, adding random variation did not make the synthetic observations more similar to the real data. Their distinguishability could not be explained simply by reduced variance or overly tight clustering, but instead reflected systematic differences in the generated feature patterns.

### Marginal fidelity did not account for discriminator AUC

Marginal fidelity was evaluated to determine whether real–synthetic distinguishability could be explained by discrepancies in individual features. Mean feature-level Kullback–Leibler divergence (KLD) is shown across datasets and generation methods in Figure 2D–F, with values closer to zero indicating greater distributional agreement. Feature-level distributions for the HIV dataset are shown in Supplementary Figure S2. Corresponding KS statistics and complete feature-level comparisons are reported in Supplementary Table S1 and Supplementary Tables S2–S4.

In HIV, CVAE produced the largest and most variable marginal discrepancies, whereas GMM showed substantially lower KLD and KS statistics. Despite this difference, both methods were completely distinguishable from the real observations, with mean discriminator AUC values of 1.00. SMOTE and GMM–SMOTE produced marginal discrepancies within the range observed for several other methods but had substantially lower AUC values of 0.59 and 0.57. Marginal discrepancy therefore did not correspond directly to source distinguishability.

Column-wise resampling provided the clearest demonstration of this distinction. Because each feature was sampled independently from its class-conditional empirical distribution, marginal distributions were preserved by construction, apart from finite-sampling variation, while observation-level alignment and cross-feature dependence were disrupted. Column-wise resampling consequently produced among the lowest KLD and KS statistics across the three datasets but was consistently more distinguishable than bootstrap. This contrast was particularly pronounced in breast cancer, where column-wise resampling achieved strong marginal agreement but an AUC of 1.00.

The absence of a one-to-one relationship between marginal discrepancy and discriminator AUC extended across breast cancer and diabetes. In breast cancer, SMOTE and GMM–SMOTE remained close to chance-level discrimination despite having greater marginal discrepancy than column-wise resampling, which was completely distinguishable. In diabetes, GMM and CVAE showed both comparatively large marginal discrepancies and near-complete distinguishability, whereas bootstrap and column-wise resampling closely reproduced individual feature distributions but remained distinguishable at AUC values of 0.72 and 0.79.

Across datasets and generators, neither KLD nor KS statistics showed a consistent correspondence with real–synthetic discriminator AUC. Marginal analyses identified important feature-level discrepancies, but could not determine whether realistic combinations of feature values or the joint structure of the source data had been preserved.

Because the fixed two-component GMM remained highly distinguishable from the real data, we examined whether its component-number specification accounted for this result. Class-specific GMMs containing (K=2)–(5) components were compared using AIC and BIC (Supplementary Table S5). AIC generally favored more components than BIC, and the criteria agreed only for HIV class 0. Regenerating the synthetic datasets using the criterion-selected component numbers did not materially reduce real–synthetic distinguishability, with discriminator AUC remaining close to 1.0 across datasets and selection strategies (Supplementary Figure S3A–C). Effects on other fidelity dimensions were dataset-dependent: component selection produced little change in HIV, AIC selection reduced the predictive-utility gap in breast cancer, and both selected models improved marginal agreement and predictive utility in diabetes (Supplementary Figure S3D–I). Thus, component-number selection improved particular fidelity criteria in some settings but did not account for the consistently high distinguishability of GMM-generated data.

### Synthetic-data generators produced dataset-dependent changes in PCA geometry

PCA was used to examine whether the six synthetic-data methods reproduced the broad multivariate geometry of each source dataset. Results are shown for HIV in Figure 3A–F and for breast cancer and diabetes in Supplementary Figures S4A–F and S5A–F, respectively. PCA was fitted separately to each standardized real dataset, and all corresponding synthetic observations were projected using the same fixed PC1 and PC2 loadings. Real and synthetic coordinates are therefore directly comparable within each dataset. The percentages displayed on the axes were calculated separately for each projected dataset and indicate how much of its total variation fell along the fixed real-data component directions.

**Figure 3:**
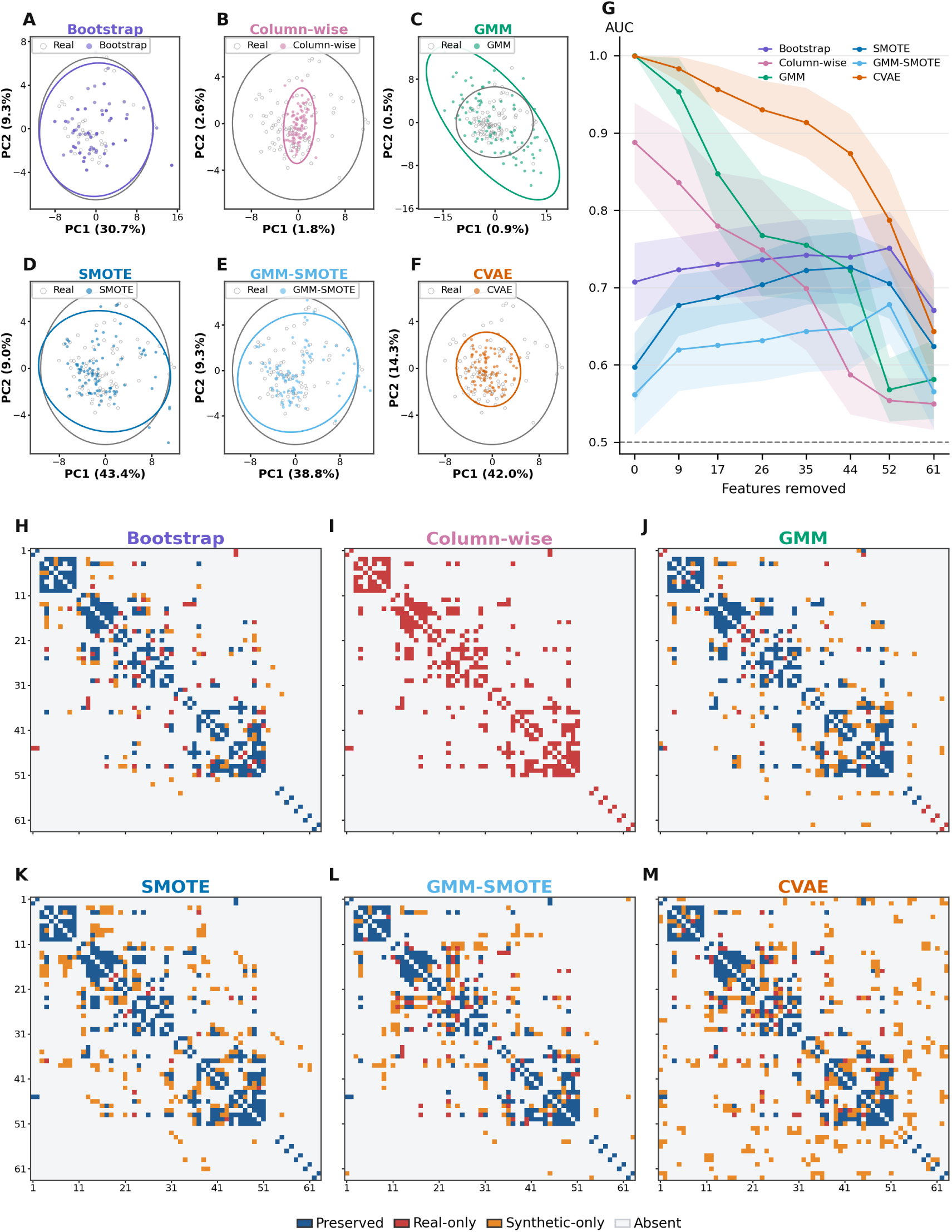
Multidimensional fidelity assessment for HIV vaccine-elicited immune responses. A– F) real and synthetic observations projected onto the first two principal components fitted to the standardized real data. Ellipses indicate 95% bivariate covariance regions for each distribution. G) Discriminator AUC as the highest-ranked features were removed; shaded regions indicate variation across repeated splits and the dashed line marks chance performance. H–M) Graphical lasso edge comparisons. Blue, red, and yellow indicate preserved, real-only, synthetic-only relationships.

In the HIV SARS-CoV-2 vaccine immunological biomarker dataset, bootstrap showed the closest visual agreement with the real data, with substantial overlap between observations and covariance ellipses (Fig. 3A). This was expected because bootstrap resampling draws complete observations with replacement from the empirical joint distribution, preserving observed feature combinations and cross-feature alignment apart from finite-sampling variation. Column-wise resampling produced the most compression for all data sets (Figures 3, S4, S5 panel B), concentrating observations near the centre of the real-data region. Unlike bootstrap, column-wise resampling retains each feature’s marginal variance but eliminates within-class covariance; because the real principal components represent directions of coordinated feature variation, this redistributes variance away from the leading real-data axes and produces a compressed PC1–PC2 projection. SMOTE and GMM–SMOTE (Fig. 3C,D) also occupied much of the same broad region, although their ellipses slightly differed from the real data in spread and orientation. CVAE, however, generated a smaller, more concentrated projection (Fig. 3F) similar to Column-wise sampling. In contrast, GMM produced a broader and differently oriented distribution (Figure 3C), indicating that the fitted mixture altered the geometry of the projected data.

Breast cancer showed related but less pronounced generator-specific differences as compared to the HIV dataset (Figure S4A–F). Bootstrap, SMOTE, and GMM-SMOTE, broadly reproduced the region occupied by the real observations, although their covariance ellipses did not align perfectly with the real-data ellipse. Column-wise resampling compressed the distribution along the principal-component directions, while CVAE generated a comparatively compact projection (albeit less than compression produced for the HIV data set). GMM retained broad overlap with the real data but altered the spread and orientation of the projected observations along PC2.

Differences between synthetic and real PCA projections were subtler in the lower-dimensional, higher population, diabetes dataset (Figure S5A–F). Here, Bootstrap, SMOTE, and GMM-SMOTE produced synthetic data with PCA ellipse overlap with real data. Similar to the HIV and Diabetes datasets, CVAE and Columnwise synthetic data produced a more concentrated ellipse projection with data that subclustered within the real-data PCA region.

Viewed across all three datasets (Table 1), distinct method-specific patterns emerged in PCA space (Figures 3, S4, S5). Bootstrap most consistently reproduced the broad location, spread, and orientation of the real observations, while SMOTE and GMM–SMOTE generally occupied the same real-data regions but showed modest differences in covariance-ellipse scale or orientation. Column-wise resampling consistently compressed the projected distributions along the leading real-data component directions. GMM produced its most pronounced expansion and reorientation in HIV, whereas its projections showed closer agreement with the real data in breast cancer and diabetes. CVAE was similarly dataset-dependent: it generated a smaller, more concentrated projection for HIV (Figure 3F), but showed substantial overlap and closer covariance-ellipse agreement in breast cancer and diabetes (Supplementary Figures S4F and S5F). Compression under CVAE was therefore most evident in HIV, which combined the smallest sample size with the greatest feature dimensionality.

Across datasets, PCA revealed that synthetic observations occupying the same broad region as the real data could nevertheless differ in their projected location, dispersion, or orientation. These differences varied jointly with the source dataset and generation method, demonstrating that marginal-distribution agreement did not ensure preservation of multivariate geometry. Conversely, close agreement in the first two principal components could not establish that pairwise or conditional-dependence structure had been preserved, emphasizing the need to interpret PCA as one component of a broader fidelity assessment.

### Pairwise correlation structure differed across generation methods

Pairwise Pearson correlation comparisons demonstrated method-dependent agreement with the real-data correlation structure within all three datasets (Table S6; Supplementary Figures S6–S8). Bootstrap showed the closest agreement for both HIV and breast cancer, with dimension-normalized correlation errors of 0.0837 and 0.0341, respectively. Column-wise resampling produced the largest discrepancy in every dataset, with RMSE*_r_* values of 0.3624 for HIV, 0.2762 for breast cancer, and 0.1828 for diabetes. Diabetes was the only dataset in which bootstrap did not have the smallest error, with GMM marginally lower than bootstrap (0.0367 versus 0.0369).

### Reverse ablation revealed distributed and generator-dependent distinguishability

Reverse feature ablation was used to determine whether real–synthetic distinguishability was concentrated in a small number of features or distributed across the feature set. The design was adapted from the feature-removal analysis used in the source immunological study (*53*) and applied across all three datasets. Features were removed in decreasing order of discriminator importance, and the discriminator was refitted using the remaining features. Results are shown for HIV in Figure 3G and for breast cancer and diabetes in Supplementary Figures S4G and S5G, respectively.

In HIV, the initially high AUC values produced by GMM, CVAE, and column-wise resampling generally declined as increasingly important features were removed. GMM and CVAE nevertheless remained substantially above chance after a large proportion of the features had been excluded, indicating that their distinguishability was distributed across the feature set rather than concentrated in only the highest-ranked biomarkers. Bootstrap, SMOTE and GMM-SMOTE maintained a comparatively stable intermediate AUC across much of the trajectory, actually displaying an increase in separability (increase in AUC) as more important features were removed, until feature number 52. Thus, Bootstrap, SMOTE and GMM-SMOTE began closer to chance and showed non-monotonic trajectories, with AUC initially increasing as features were removed before declining when few features remained.

For the breast cancer dataset (Figure S4G) showed similar method-specific ablation patterns. Column-wise resampling remained nearly completely distinguishable until most of the 16 features had been removed, whereas GMM declined rapidly but remained above chance throughout much of the trajectory. CVAE showed a more gradual reduction from its intermediate initial AUC, while bootstrap remained comparatively stable. SMOTE and GMM–SMOTE again became more distinguishable after removal of their highest-ranked features, indicating that the contribution of individual features to discrimination was not uniformly additive.

For the diabetes dataset (Figure S5G), GMM and CVAE retained high discriminator AUC after several of the eight features had been removed, before declining as the available feature set became small. Column-wise resampling approached chance more rapidly, while bootstrap remained relatively stable until the final ablation levels. SMOTE and GMM–SMOTE initially declined toward chance but increased again when only a small number of features remained, producing pronounced non-monotonic trajectories.

Across datasets, generators with high initial discriminator AUC generally remained distinguishable after removal of multiple top-ranked features, supporting the presence of discrepancies distributed across the feature set. The non-monotonic behavior of the interpolation-based methods further showed that individual features could either expose or obscure real–synthetic differences depending on the remaining feature context. Reverse-ablation patterns were therefore shaped jointly by the source dataset and generation method, rather than by a common subset of universally discriminating features.

### Generators produced distinct patterns of preserved, lost, and synthetic-only conditional dependencies

Graphical Lasso was used to compare sparse conditional-dependence structure between real and synthetic data. For each feature pair, the corresponding matrix cell was colored blue when the edge was present in both networks (preserved), red when it appeared only in the real network (lost), orange when it appeared only in the synthetic network (synthetic-only), and light gray when it was absent from both. Because the estimated networks were undirected, classifications were symmetric about the matrix diagonal. Edge-comparison matrices are shown for HIV in Figure 3H–M and for breast cancer and diabetes in Supplementary Figures S4H–M and S5H–M, respectively; edge-recovery proportion, synthetic-only edge proportion, and Frobenius distances are provided in Supplementary Table 3. For each data set feature ordering was derived once from the real-data partial-correlation structure and then held fixed across all graphical lasso analyses for that data set (see Supplementary Table S7). Therefore, differences in color between panels reflect changes in estimated edge status rather than changes in feature arrangement.

The HIV dataset showed the most pronounced differences among generators (Figure 3H–M). A representative regularization path is shown in Figure S10. Bootstrap retained the clustered structure of the real network with fewer lost or synthetic-only edges than the other methods. This agreement was expected because bootstrap resampling preserves complete observations and therefore retains their original feature combinations. Nevertheless, sampling with replacement duplicates some observations and excludes others, changing their empirical weights in the assembled dataset. When Graphical Lasso is re-estimated, these finite-sample changes can cause conditional-dependence edges to appear or disappear. The discrepancies observed under bootstrap (Figure 3H, yellow and red matrix elements) therefore provide a reference for resampling-induced structural variability, rather than artifacts attributable to a learned generative model. In contrast, column-wise resampling converted much of the real network into real-only edges (Figure 3I), consistent with widespread loss of feature interdependencies. Column-wise resampling was chosen as a control sampling method for this reason. GMM preserved substantial portions of the real network while both losing and introducing individual relationships (Figure 3J). This mixed pattern is consistent with modeling the joint distribution through component-specific means and covariance matrices: broad covariance structure can be retained, while mixture approximation and component smoothing alter the precision matrix sufficiently for individual conditional dependencies to appear or disappear. SMOTE also retained major regions of the real network (Figure 3K), as interpolation between neighboring observations preserves aspects of their local multivariate structure. However, concentrating observations along interpolated trajectories changes their relative density and covariance, potentially producing synthetic-only partial-correlation edges. GMM–SMOTE restricts interpolation using the fitted mixture structure and may therefore better preserve relationships within modeled regions (Figure 3L), but it remains susceptible to interpolation-induced dependencies and inaccuracies in the estimated mixture components. Overall, based on the Graphical Lasso analysis on the HIV dataset, GMM performed the best in terms of introducing the least number of synthetic-only relationships, preserving the most real relationships, and minimizing the loss of real relationships. An exploratory t-SNE overlay of the most discriminator-important features did not reveal consistent spatial organization across methods, but highlighted method-specific preserved, lost, and synthetic-only conditional dependencies involving these features (Supplementary Figure S9).

Breast cancer showed a similar but less pronounced separation among methods (Figure S4H– M). Bootstrap again displayed extensive preservation, whereas column-wise resampling produced the greatest concentration of lost relationships. GMM, SMOTE, and GMM–SMOTE retained substantial portions of the real network but exhibited localized combinations of lost and synthetic-only edges. CVAE also preserved much of the real structure while introducing synthetic-only relationships across several regions of the matrix (Figure S4M).

The diabetes dataset, which boasts the highest number of individuals but fewest features, showed more extensive edge preservation across all generators (Figure S5H–M). Bootstrap preserved most real-data relationships while introducing only a small number of synthetic-only edges. GMM and SMOTE likewise retained most of the real network, although each lost a small number of relationships and introduced additional edges. GMM–SMOTE and CVAE preserved the observed real-network structure particularly closely but still produced several synthetic-only dependencies. Column-wise resampling retained more real-data edges than it did in HIV or breast cancer, but remained distinct from bootstrap through its combination of lost and synthetic-only relationships. Thus, generation methods differed not only in the overall extent of conditional-dependence discrepancy but also in its direction: some primarily eliminated relationships present in the real data, whereas others preserved those relationships while introducing additional structure detectable only in the synthetic data. These single feature-level patterns varied across datasets and revealed structural changes that could not be identified through marginal agreement, PCA geometry, or real–synthetic distinguishability alone.

### Within-class permutation produced dataset-dependent distinguishability and edge loss

Progressive within-class permutation was used to disrupt cross-feature alignment while preserving the class-conditional marginal distribution of every feature. Across all three datasets, increasing permutation generally increased discriminator AUC and reduced recovery of real-data Graphical LASSO edges, although the trajectories were dataset-dependent and not strictly monotonic (Table 3 and Fig. S11).

**Table 3:** Within-class permutation sensitivity. Values are mean (SD) over 10 repeats. Permutation was performed independently within each outcome class and feature, preserving class-conditional marginal distributions while disrupting cross-feature dependence. AUC measures how well the discriminator separates original from permuted records, reported in a direction-invariant form; edge recovery is the proportion of real-data Graphical LASSO edges retained after permutation; Frobenius distance is the Frobenius norm of the difference between the original and permuted precision matrices.

| Permutation (%) | HIV |  |  | Breast cancer |  |  | Diabetes |  |  |
| --- | --- | --- | --- | --- | --- | --- | --- | --- | --- |
|  | AUC | Edge rec. | Frob. dist. | AUC | Edge rec. | Frob. dist. | AUC | Edge rec. | Frob. dist. |
| 0 | 0.500 (0.000) | 1.000 (0.000) | 0.000 (0.000) | 0.500 (0.000) | 1.000 (0.000) | 0.000 (0.000) | 0.500 (0.000) | 1.000 (0.000) | 0.000 (0.000) |
| 10 | 0.576 (0.041) | 0.667 (0.048) | 1.633 (0.177) | 0.674 (0.015) | 0.886 (0.030) | 0.852 (0.084) | 0.522 (0.011) | 0.967 (0.032) | 0.478 (0.071) |
| 20 | 0.522 (0.017) | 0.356 (0.061) | 2.521 (0.087) | 0.852 (0.022) | 0.751 (0.045) | 1.436 (0.095) | 0.586 (0.015) | 0.957 (0.027) | 0.815 (0.043) |
| 30 | 0.585 (0.061) | 0.167 (0.039) | 2.883 (0.072) | 0.938 (0.013) | 0.610 (0.023) | 1.814 (0.037) | 0.619 (0.021) | 0.876 (0.056) | 1.008 (0.080) |
| 40 | 0.607 (0.062) | 0.097 (0.038) | 2.956 (0.080) | 0.983 (0.006) | 0.529 (0.023) | 2.029 (0.036) | 0.688 (0.017) | 0.857 (0.039) | 1.236 (0.055) |
| 50 | 0.701 (0.033) | 0.037 (0.017) | 3.078 (0.051) | 0.993 (0.002) | 0.455 (0.028) | 2.132 (0.028) | 0.698 (0.034) | 0.829 (0.060) | 1.365 (0.067) |
| 60 | 0.768 (0.060) | 0.018 (0.016) | 3.125 (0.031) | 0.996 (0.002) | 0.409 (0.025) | 2.243 (0.015) | 0.718 (0.017) | 0.810 (0.103) | 1.429 (0.032) |
| 70 | 0.775 (0.040) | 0.008 (0.008) | 3.154 (0.033) | 0.999 (0.001) | 0.364 (0.021) | 2.303 (0.009) | 0.739 (0.024) | 0.800 (0.107) | 1.524 (0.061) |
| 80 | 0.791 (0.051) | 0.002 (0.004) | 3.159 (0.007) | 0.999 (0.000) | 0.329 (0.017) | 2.347 (0.017) | 0.757 (0.025) | 0.719 (0.104) | 1.589 (0.066) |
| 90 | 0.801 (0.056) | 0.002 (0.003) | 3.160 (0.006) | 1.000 (0.000) | 0.307 (0.015) | 2.375 (0.013) | 0.778 (0.028) | 0.757 (0.091) | 1.611 (0.050) |
| 100 | 0.738 (0.084) | 0.000 (0.000) | 3.162 (0.001) | 1.000 (0.000) | 0.306 (0.010) | 2.375 (0.014) | 0.764 (0.027) | 0.767 (0.091) | 1.649 (0.023) |

In HIV, edge recovery declined rapidly from 0.667 at 10% permutation to 0.037 at 50% and 0.000 at complete permutation. Discriminator AUC increased more gradually, reaching 0.701 at 50%, peaking at 0.801 at 90%, and declining to 0.738 at complete permutation. Breast cancer showed the strongest increase in distinguishability: AUC rose from 0.674 at 10% permutation to 0.993 at 50% and 1.000 at complete permutation, while edge recovery declined from 0.886 to 0.306. Changes were less pronounced in diabetes, where complete permutation produced an AUC of 0.764 while retaining 76.7% of real-data edges. Frobenius distance also increased with permutation across all three datasets, indicating that the permuted precision matrices moved progressively farther from the original Graphical LASSO structure. Thus, disrupting within-class feature alignment was sufficient to produce detectable real–synthetic differences without changing marginal distributions, while the differing AUC and edge-recovery trajectories confirmed that these measures captured complementary consequences of structural disruption.

## Discussion

We evaluated synthetic-data fidelity as a multidimensional construct (see Figure 1 for a schematic of our approach), asking which statistical, structural, and task-relevant properties were retained rather than reducing performance to a single quality score. Across three two-group biomedical datasets spanning contrasting sample-size and dimensionality regimes (Table 1), marginal agreement, predictive utility, real–synthetic distinguishability, low-dimensional geometry, and conditional-dependence preservation frequently yielded discordant assessments. Synthetic data could reproduce individual feature distributions or retain class-predictive utility while remaining distinguishable from real observations and exhibiting lost or synthetic-only dependencies. PCA and network discrepancies were most pronounced in the small-(n), high-(p) HIV dataset, generally intermediate in breast cancer, and subtler in the large-(n), low-(p) diabetes dataset; nevertheless, several generators, such as GMM, remained highly distinguishable in every setting (Table 2). It is well known and understood generative difficulty also depends on the effective dimensionality and dependence structure of the source distribution, including its within- and between-class dependencies (*70, 71*). We find fidelity to depend jointly on the generator and source-data characteristics, reinforcing the need to evaluate synthetic data from a multidimensional perspective against the properties required for its intended use.

Conventional fidelity measures provide complementary perspectives because each interrogates a distinct property of the real–synthetic relationship (*37*). Marginal tests assess features individually, PCA summarizes variation along a small number of linear directions, predictive utility asks whether group membership remains learnable, and real–synthetic discrimination detects systematic differences in the joint feature distribution. These objectives can diverge under class-conditional synthesis because a generator may retain between-group differences sufficient for classification while distorting the combinations of features occurring within each group. Column-wise resampling provided the clearest demonstration: it preserved class-conditional marginal distributions by construction and retained substantial predictive utility, yet breaking observation-level alignment increased real–synthetic distinguishability, compressed the data along the leading real-data principal-component directions, and eliminated many conditional dependencies (e.g. Figures 3I and 3B).

Feature-level analyses extended this conclusion beyond the deliberately structure-breaking control. Across all dataset–method combinations, only two feature comparisons, both for CVAE in HIV, had unadjusted Welch ( *p* ≤ 0.05), although several corresponding synthetic datasets remained readily distinguishable from the real data (Supplementary Tables S1-S4). Mean feature-level KLD and KS statistics likewise showed no consistent correspondence with discriminator AUC (Figure 2A–F). In HIV, GMM produced lower KLD and a lower mean KS statistic than CVAE of 0.192 versus 0.282, yet both achieved an AUC of 1.00; SMOTE and GMM–SMOTE had mean KS statistics of 0.111 and 0.133, respectively, but substantially lower AUC of ∼0.6. In breast cancer, column-wise resampling combined a mean KS statistic of 0.035 with an AUC of 1.00, whereas SMOTE and GMM–SMOTE had slightly larger mean KS statistics 0.050 and 0.049 but remained close to chance discrimination. Even in diabetes, mean KS statistics of 0.030 and 0.027 for bootstrap and column-wise resampling coexisted with AUC values of 0.72 and 0.79. Marginal agreement can therefore establish the plausibility of individual features but cannot determine whether those values occur in multivariate configurations supported by the source data.

For all three datasets GMM-generated synthetic data demonstrated near-complete real–synthetic separation (Figure 2A). Similarly, the CVAE-generated data for the HIV and diabetes data sets showed near-complete real–synthetic separation, however, the mean AUC for the breast cancer dataset dropped to 0.8. These differences in distinguishability were accompanied by method-specific changes in PCA geometry, which for GMM, indicate expanded variance in the projected component space, and for CVAE-generated data more compact ellipses, indicating attenuated variation and contraction toward the centre of the real-data distribution (Figs. 3, S5, S4 panels A– F). Despite this, structural comparisons showed that GMM preserved more real-feature relationships and produced fewer synthetic-only relationships (e.g. Figure 3J and M). Conversely, SMOTE and GMM–SMOTE combined high predictive utility with near-chance discriminator AUC in HIV and breast cancer, although their multivariate geometry and conditional-dependence networks were not fully preserved. Similar discrepancies between target-specific utility and global structural fidelity have been observed in broader tabular-generator benchmarks (*37*).

Whereas real–synthetic discriminator AUC establishes whether sample origin is learnable, it does not identify which structural differences support that discrimination. Graphical Lasso provides this relationship-level resolution by estimating a sparse precision matrix, in which nonzero off-diagonal elements indicate conditional associations between features after accounting for the remaining measured variables under a Gaussian graphical-model approximation (*52*). Comparing the resulting real and synthetic networks allowed individual relationships to be classified as preserved, real-only or lost, and synthetic-only (Figure 3H–M). The sampling controls provided intuitive reference points for interpreting these classifications: bootstrap retained much of the real network but altered some estimated edges because finite resampling changes observation multiplicities and, consequently, the empirical covariance structure, whereas column-wise resampling removed much of the network by intentionally disrupting cross-feature alignment. The learned generators exhibited method-specific combinations of lost and synthetic-only relationships, including cases in which marginal agreement, predictive utility, PCA geometry, or discriminator AUC appeared comparatively favorable. Unlike comparisons requiring a ground-truth causal graph, this observational network audit requires neither known causal structure nor assignment of edge direction (*37*). Its edges nevertheless represent regularized conditional associations, not causal or necessarily biological interactions. Graphical Lasso therefore does not provide a standalone quality score; it identifies where generation alters the dependence structure that subsequent scientific or predictive analyses may exploit.

Across the PCA and Graphical LASSO analyses, structural discrepancies were generally greatest for HIV, intermediate for breast cancer, and smallest for diabetes, broadly tracking the progression from a low-(n), high-(p) dataset to larger datasets with fewer features. This ordering was not universal across fidelity criteria or generators, indicating that performance depended on the balance between the information available within each class and the complexity of the fitted model, rather than on total sample size (n) and feature count (p) alone. Because synthesis was performed separately by outcome class, the relevant covariance-rank bound is (rank(Σ*_c_*) ≤ min( *p*, *n_c_* − 1)). For HIV, the class containing 23 participants and 63 features therefore had a maximum empirical covariance rank of 22, whereas both classes in the breast-cancer and diabetes datasets contained substantially more observations than features (Table 1). Neither high nor low rank is uniformly favorable. Higher effective rank indicates that variation is distributed across more substantively occupied directions and increases covariance-estimation complexity (*72*), whereas sample-imposed rank deficiency when ( *p* ≥ *n_c_*) makes the class-specific covariance estimate necessarily singular and leaves variation outside the observed subspace unsupported without additional assumptions or regularization (*73*). This distinction is particularly important for the full-covariance GMM, which estimates *p*( *p* + 1)/2 unique covariance elements per component—2,016 for HIV, compared with 465 for breast cancer and 36 for diabetes—before accounting for component means and weights (*41, 44*). Although covariance regularization ensures numerical invertibility, it cannot supply information about directions that are weakly supported by the observed data. This rank–complexity imbalance may help explain the pronounced expansion and reorientation of the HIV GMM projection. Although AIC- or BIC-based component selection improved marginal agreement or predictive utility in some datasets, discriminator AUC remained near (1.0) across all datasets and specifications, indicating that component number affected particular dimensions of fidelity but did not account for the persistent real–synthetic distinguishability of GMM-generated data (Supplementary Table S5; Supplementary Figure S3). The fixed 16-dimensional CVAE latent representation likewise imposed substantially greater dimensional reduction for HIV (*p* = 63) than for breast cancer (*p* = 30), and none for diabetes (*p* = 8), consistent with the greater attenuation of projected HIV variation (*50,51*). Nevertheless, GMM and CVAE remained highly distinguishable even in diabetes, where covariance estimation was comparatively well supported. Sample size, dimensionality, and rank therefore constrain generator performance but do not determine it; fidelity also depends on class separation, distributional shape, feature dependence, and the assumptions and complexity of the generating architecture (*70, 71*).

The sensitivity analyses further showed that real–synthetic discrepancies were structured, distributed, and dependent on feature context (Fig. S1). Adding independent, feature-scaled Gaussian noise generally increased discriminator AUC rather than reducing it, while GMM and CVAE remained nearly completely distinguishable across the evaluated noise levels. Insufficient dispersion or excessive concentration therefore could not explain the observed separation: random perturbation can broaden feature-level variation, but it cannot reconstruct coordinated relationships and likely produces implausible multivariate combinations. Progressive within-class permutation isolated this effect by preserving class-conditional marginal distributions while increasingly disrupting feature alignment across observations (Table 3). Greater permutation was generally accompanied by higher discriminator AUC and lower Graphical LASSO edge recovery, although the magnitude and trajectory differed across datasets. At complete permutation, HIV retained none of its real-data edges but reached an AUC of only (0.738), whereas breast cancer reached an AUC of (1.000) while retaining (30.6%) of its edges, and diabetes reached an AUC of (0.764) while retaining (76.7%). Distinguishability and sparse edge recovery therefore captured related but non-equivalent consequences of structural disruption. Reverse ablation further showed that generators with high initial AUC generally remained distinguishable after multiple top-ranked features were removed, indicating that discrepancies were distributed across the feature set rather than attributable to a single biomarker or small subset (Figure 3G; Supplementary Figures S4G and S5G). Non-monotonic ablation trajectories, particularly for interpolation-based methods, further indicated that feature contributions were context-dependent and could expose or mask discrepancies depending on the remaining feature set. Collectively, these analyses show that improving isolated marginals or adding nonspecific variance is insufficient; fidelity depends on preserving coordinated multivariate structure across features.

Structural fidelity is particularly consequential when shared synthetic longitudinal biomarker data are used to calibrate within-host compartmental ODE models or train ML and AI models. Simulation-based calibration studies have demonstrated that observation timing, measurement noise, measured biological compartments, and model complexity can materially alter practical identifiability and parameter recovery in within-host and epidemic ODE models (*16, 74–77*). These models infer biological mechanisms and timescales from coordinated variation across biomarkers, time points, and physiological compartments (*78–86*) and are increasingly being used to generate virtual cohorts (*87, 88*); relationships lost, attenuated, or synthetically-created by the generation method can therefore alter the information available both to distinguish competing mechanisms and to estimate the rates and timescales governing their dynamics. ML and AI models similarly learn predictive structure from relationships among features, outcomes, and time points. In a tuberculosis biomarker application, models trained on synthetic datasets of differing sizes exhibited markedly different performance when evaluated on an independent real-world cohort, and increasing synthetic sample size did not consistently improve generalization (*89*). Synthetic-only dependencies may consequently introduce spurious relationships, distort feature attribution, or impair generalization to real patients even when aggregate performance appears favorable. Thus, even when marginal distributions and individual trajectories appear plausible, altered covariance or conditional-dependence structure can reshape both the parameter-estimation landscape of mechanistic models and the decision functions learned by data-driven models. This risk is especially important for investigators who did not design the generation procedure and may reasonably assume that shared, or publicly-available, privacy-preserving synthetic data retain the relationships required for valid inference. We therefore propose that shared synthetic biomedical datasets, particularly those containing longitudinal or multicomponent biomarker measurements be accompanied by a multivariate fidelity assessment as a minimum reporting standard for downstream use, documenting whether relevant within-time, cross-time, cross-compartment, and class-conditional relationships have been preserved, lost, or synthetic-only. Graphical Lasso networks provide a structural audit of dependencies that mechanistic calibration or data-driven learning may implicitly exploit. Without such a feature-level assessment, neither mechanistic conclusions nor learned predictive behavior can be confidently separated from distortions introduced during synthetic-data generation.

Synthetic-data fidelity should not be reduced to a universal quality score, but evaluated as a fit-for-purpose account of the statistical, structural, and task-relevant properties retained from the source data. We therefore propose that shared synthetic biomedical datasets be accompanied by a multidimensional fidelity profile documenting marginal agreement, multivariate geometry, distinguishability, predictive utility, and feature dependencies preserved, lost, or synthetic-only. Making these properties explicit transforms trust from an assumption into evidence and is essential if synthetic data are to provide a reliable foundation for biomedical discovery, mechanistic inference, AI-enabled research, and regulatory science.

## Data & Code availability

Code supporting the analyses presented in this manuscript is publicly available on GitHub (*90*).

## Data Availability

All data produced in the present study are available upon reasonable request to the authors.

https://github.com/TonyTran03/synthetic_data_paper_code

## Acknowledgments

C.S.K. Acknowledges Bridge Funding Support for this project from the University of Guelph.

## Competing interests

We declare no competing interests.

## CRediT author statements

Conceptualization: C.S.K., M.S.G. Data Curation: T.T. Visualization: T.T. Methodology: C.S.K., M.S.G., T.T. Investigation: All authors. Validation: T.T. Formal Analysis: T.T. Supervision: C.S.K., M.S.G. Writing – Original Draft: C.S.K., T.T. Writing - Review & Editing: All authors. Funding Acquisition: C.S.K.

## S1 Supplementary Classification Analyses

### S1.1 Noise-sensitivity analysis

**Figure S1:**
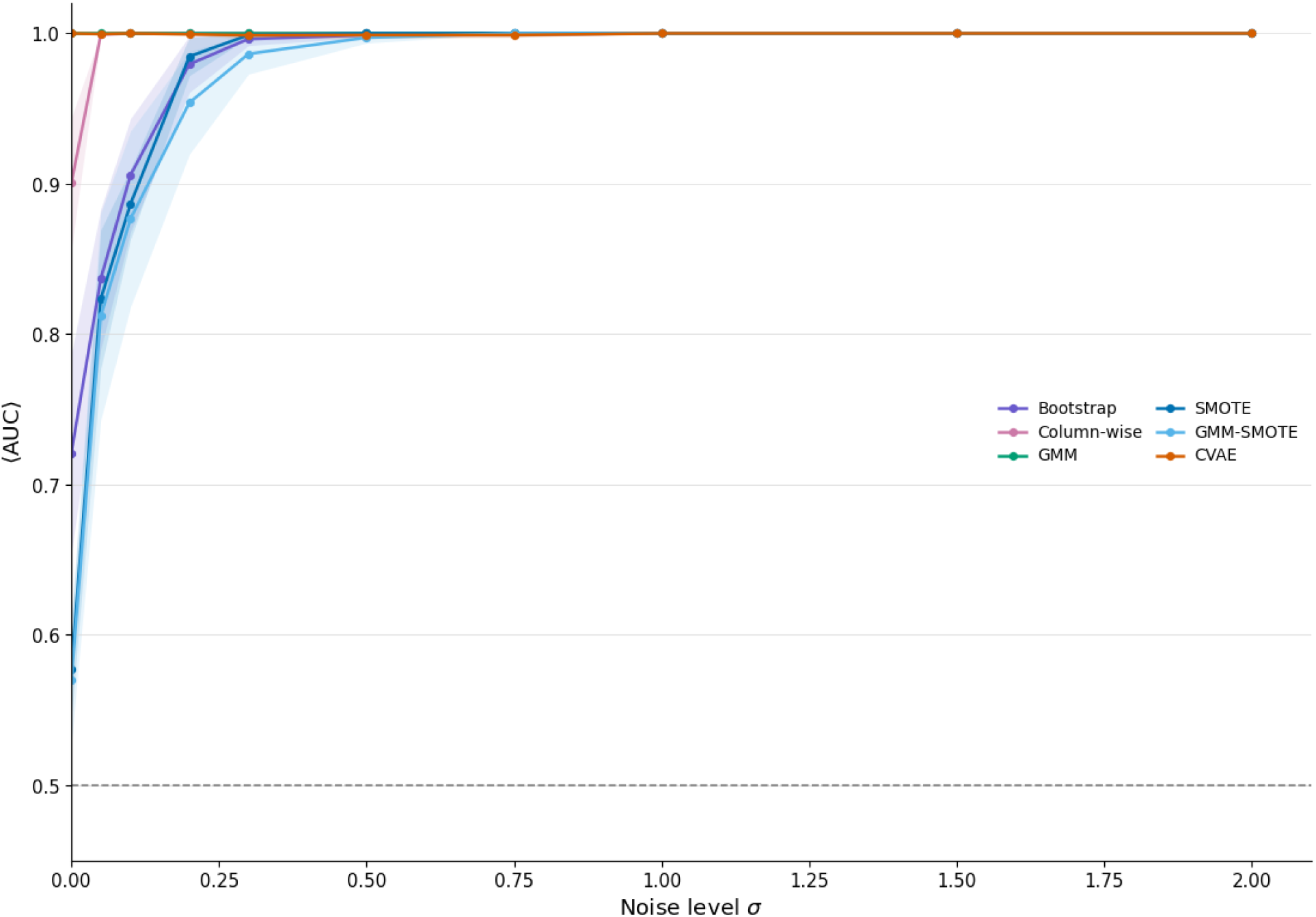
Real–synthetic discriminator AUC after adding independent Gaussian noise to each synthetic feature, scaled by the corresponding real-feature standard deviation. Results are shown across noise magnitudes *σ* = 0 to 2, with five repetitions per level. Increasing random variation generally increased rather than reduced distinguishability.

## S2 Feature-level marginal distributions

**Figure S2:**
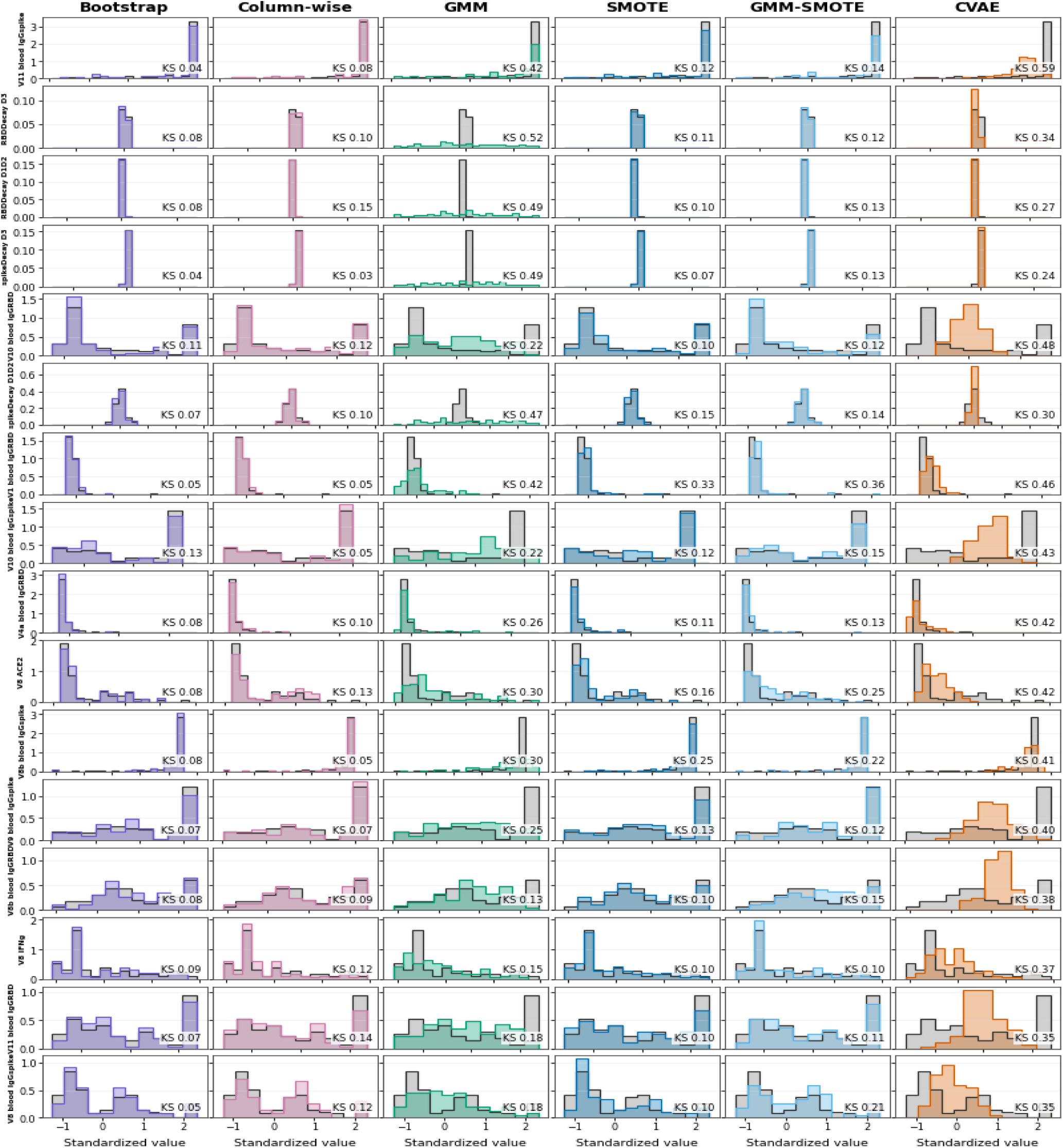
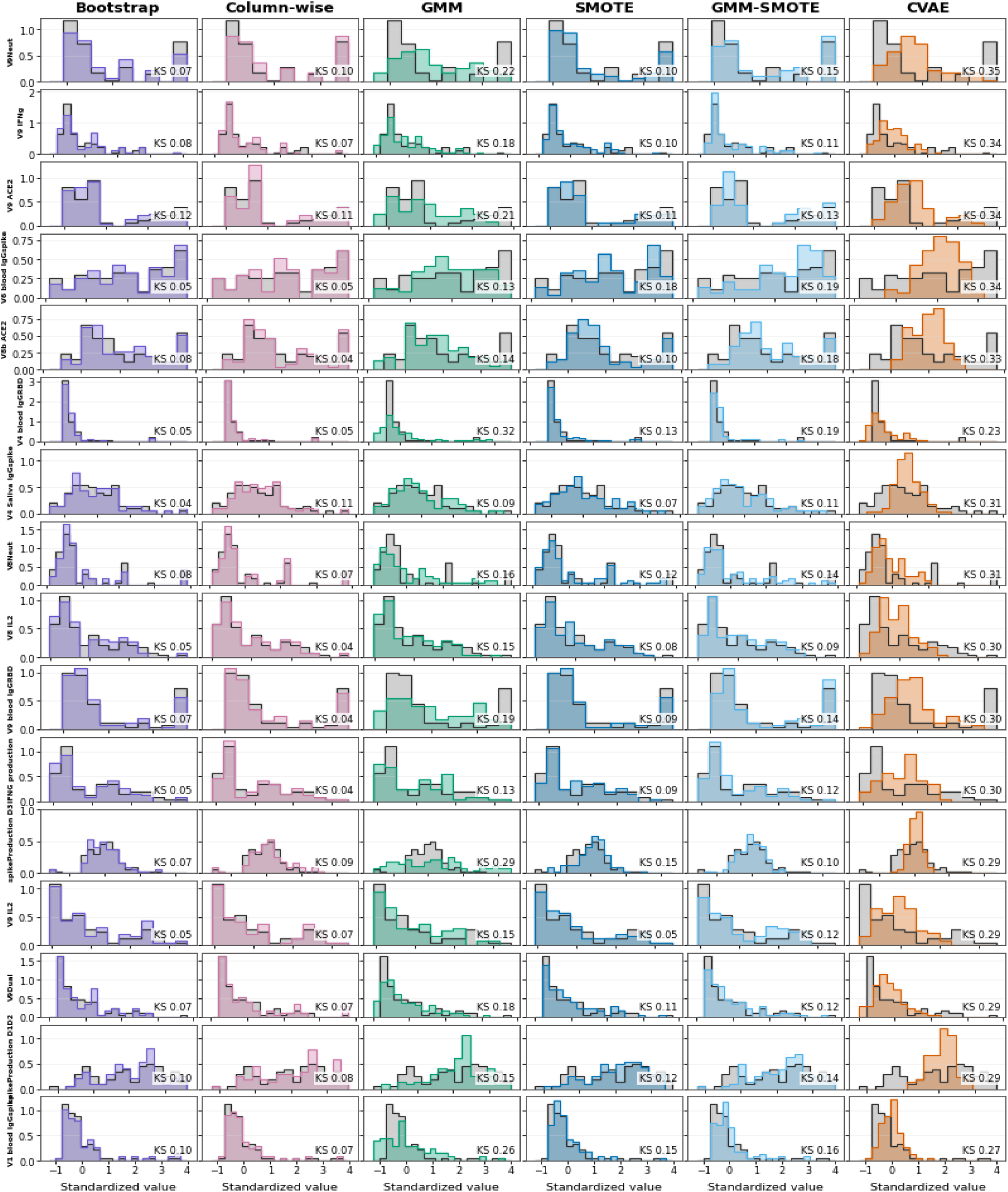
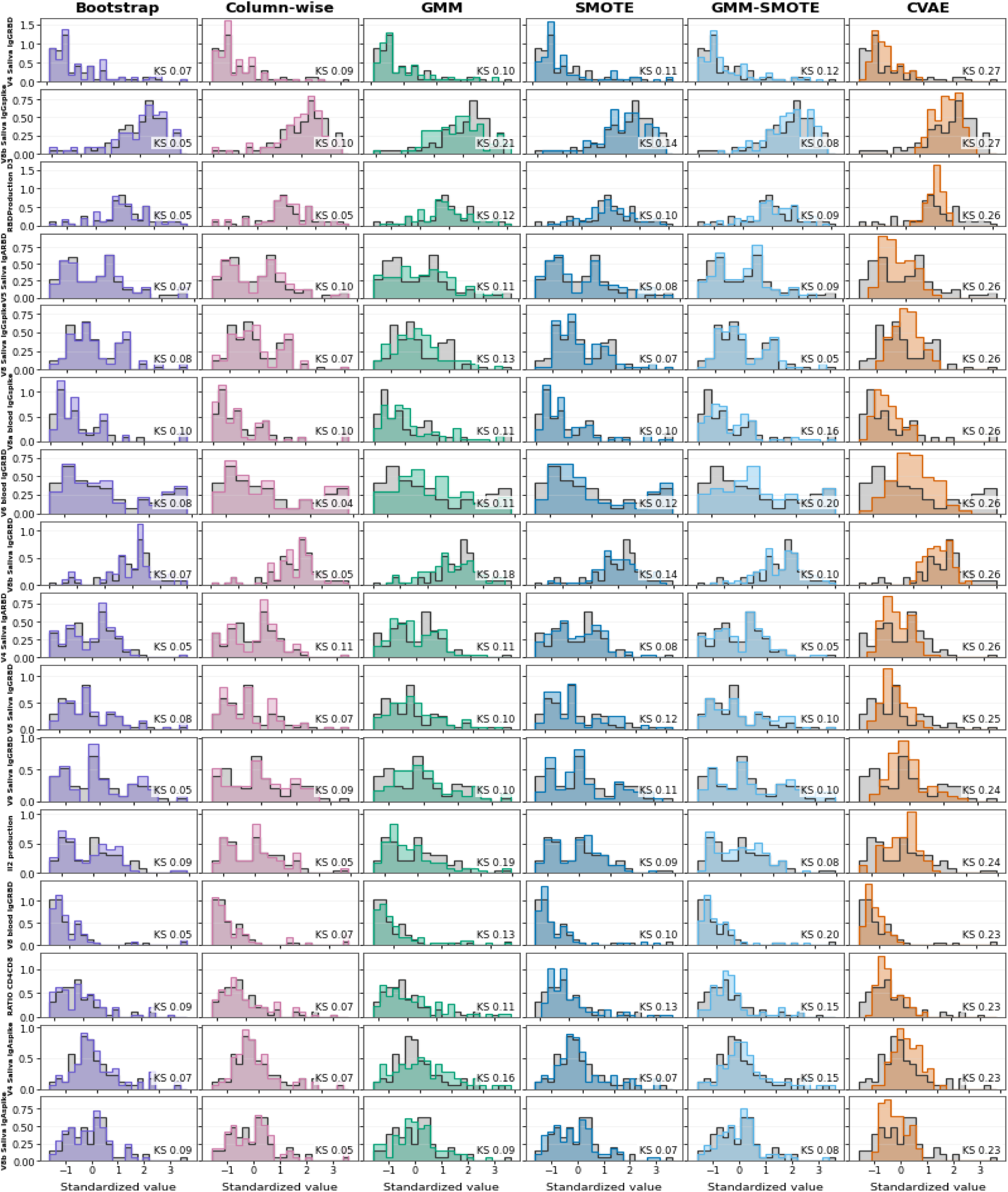
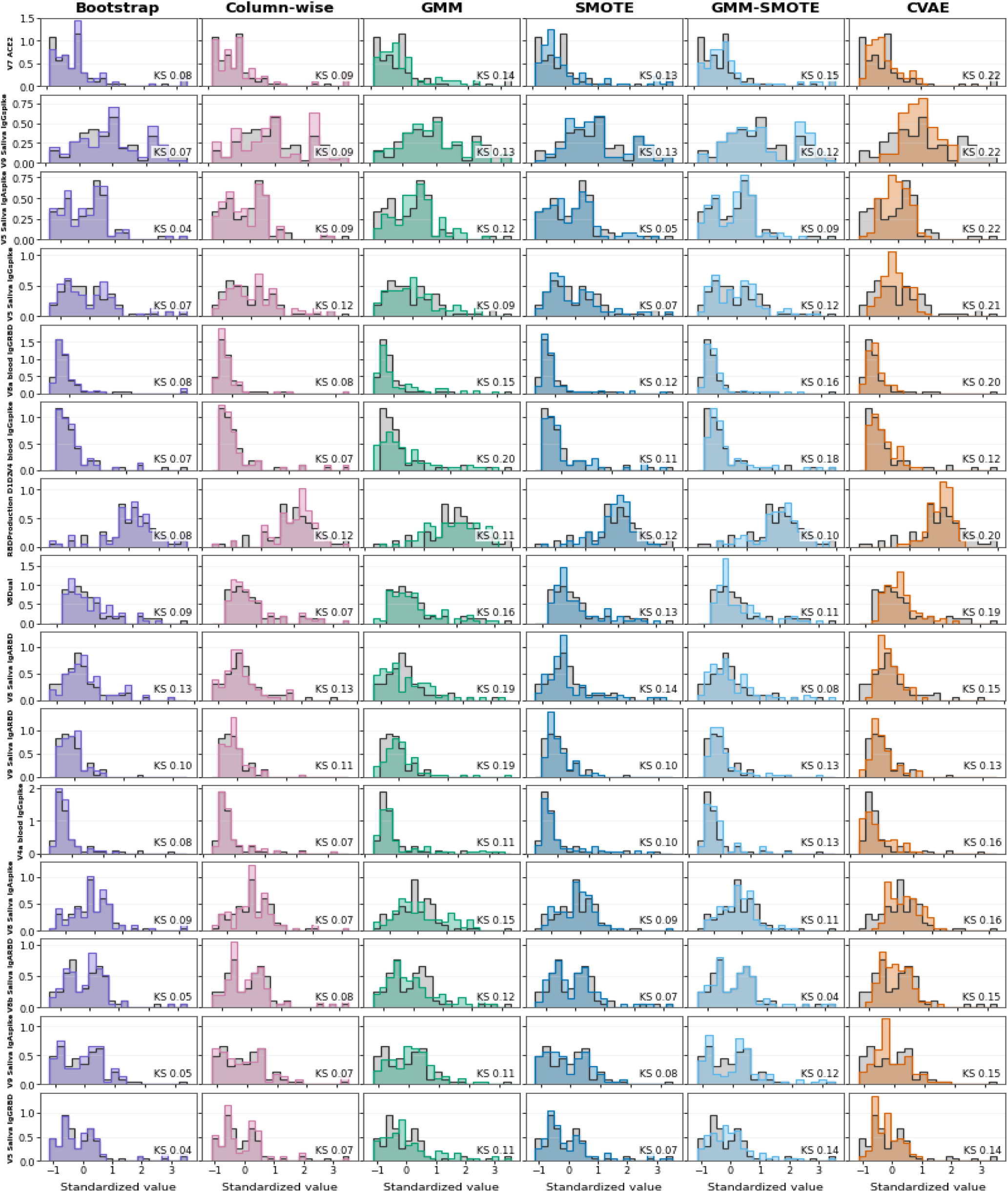
Feature-level marginal distributions for the HIV dataset across synthetic-data generation methods. Grey histograms show the real observations, and coloured histograms show the corresponding synthetic observations.

### S2.1 Feature-level marginal comparisons

**Table S1:** Summary of feature-level marginal comparisons for all datasets and all six synthesis methods. Welch p-values are unadjusted; KS denotes the two-sample Kolmogorov–Smirnov statistic.

| Dataset | Method | Features | $p \leq 0.05$ | Median $p$ | Mean KS |
| --- | --- | --- | --- | --- | --- |
| HIV | Bootstrap | 63 | 0 | 0.789 | 0.072 |
| HIV | Column-wise | 63 | 0 | 0.597 | 0.081 |
| HIV | GMM | 63 | 0 | 0.631 | 0.192 |
| HIV | SMOTE | 63 | 0 | 0.732 | 0.111 |
| HIV | GMM-SMOTE | 63 | 0 | 0.595 | 0.133 |
| HIV | CVAE | 63 | 2 | 0.427 | 0.282 |
| Breast cancer | Bootstrap | 30 | 0 | 0.792 | 0.033 |
| Breast cancer | Column-wise | 30 | 0 | 0.661 | 0.035 |
| Breast cancer | GMM | 30 | 0 | 0.721 | 0.108 |
| Breast cancer | SMOTE | 30 | 0 | 0.613 | 0.050 |
| Breast cancer | GMM-SMOTE | 30 | 0 | 0.702 | 0.049 |
| Breast cancer | CVAE | 30 | 0 | 0.499 | 0.074 |
| Diabetes | Bootstrap | 8 | 0 | 0.593 | 0.030 |
| Diabetes | Column-wise | 8 | 0 | 0.760 | 0.027 |
| Diabetes | GMM | 8 | 0 | 0.723 | 0.145 |
| Diabetes | SMOTE | 8 | 0 | 0.732 | 0.079 |
| Diabetes | GMM-SMOTE | 8 | 0 | 0.552 | 0.071 |
| Diabetes | CVAE | 8 | 0 | 0.356 | 0.177 |

**Table S2:** Complete feature-level marginal comparisons for HIV across all six synthesis methods. Difference is synthetic minus real; Welch p-values are unadjusted.

| Method | No. | Feature | n real | n synthetic | Real mean | Synthetic mean | Difference | Welch t | p | KS |
| --- | --- | --- | --- | --- | --- | --- | --- | --- | --- | --- |
| Bootstrap | 1 | spikeProduction_D1D2 | 91 | 91 | 3.141 | 3.145 | +0.003633 | -0.194 | 0.847 | 0.099 |
| Bootstrap | 2 | spikeDecay_D1D2 | 91 | 91 | 0.02447 | 0.02443 | -3.53e-05 | +0.250 | 0.803 | 0.066 |
| Bootstrap | 3 | spikeProduction_D3 | 91 | 91 | 0.6235 | 0.6236 | +8.548e-05 | -0.143 | 0.886 | 0.066 |
| Bootstrap | 4 | spikeDecay_D3 | 91 | 91 | 0.008704 | 0.008696 | -8.008e-06 | +0.233 | 0.816 | 0.044 |
| Bootstrap | 5 | RBDProduction_D1D2 | 91 | 91 | 2.405 | 2.412 | +0.007697 | -0.508 | 0.612 | 0.077 |
| Bootstrap | 6 | RBDDecay_D1D2 | 91 | 91 | 0.02156 | 0.02156 | +6.062e-06 | -0.175 | 0.861 | 0.077 |
| Bootstrap | 7 | RBDProduction_D3 | 91 | 91 | 0.817 | 0.8173 | +0.0002699 | -0.077 | 0.939 | 0.055 |
| Bootstrap | 8 | RBDDecay_D3 | 91 | 91 | 0.01138 | 0.01137 | -7.919e-06 | +0.243 | 0.808 | 0.077 |
| Bootstrap | 9 | V1_blood_IgGspike | 91 | 91 | 6.896 | 8.03 | +1.134 | -0.959 | 0.339 | 0.099 |
| Bootstrap | 10 | V4_blood_IgGspike | 91 | 91 | 203 | 211.9 | +8.911 | -0.203 | 0.839 | 0.066 |
| Bootstrap | 11 | V4a_blood_IgGspike | 91 | 91 | 125.3 | 134.3 | +8.918 | -0.261 | 0.795 | 0.077 |
| Bootstrap | 12 | V6_blood_IgGspike | 91 | 91 | 924.7 | 953 | +28.38 | -0.408 | 0.684 | 0.055 |
| Bootstrap | 13 | V8_blood_IgGspike | 91 | 91 | 483.4 | 488.7 | +5.268 | -0.082 | 0.934 | 0.055 |
| Bootstrap | 14 | V8a_blood_IgGspike | 91 | 91 | 339.4 | 332.8 | -6.625 | +0.135 | 0.893 | 0.099 |
| Bootstrap | 15 | V8b_blood_IgGspike | 91 | 91 | 1350 | 1387 | +37.07 | -0.877 | 0.382 | 0.077 |
| Bootstrap | 16 | V9_blood_IgGspike | 91 | 91 | 1015 | 1003 | -11.96 | +0.171 | 0.865 | 0.066 |
| Bootstrap | 17 | V10_blood_IgGspike | 91 | 91 | 962.7 | 873.6 | -89.14 | +1.114 | 0.267 | 0.132 |
| Bootstrap | 18 | V11_blood_IgGspike | 91 | 91 | 1315 | 1323 | +8.055 | -0.173 | 0.863 | 0.044 |
| Bootstrap | 19 | V1_blood_IgGRBD | 91 | 91 | 5.138 | 6.142 | +1.005 | -0.686 | 0.494 | 0.055 |
| Bootstrap | 20 | V4_blood_IgGRBD | 91 | 91 | 226 | 297.1 | +71.03 | -0.684 | 0.495 | 0.055 |
| Bootstrap | 21 | V4a_blood_IgGRBD | 91 | 91 | 159.5 | 217.6 | +58.16 | -0.595 | 0.553 | 0.077 |
| Bootstrap | 22 | V6_blood_IgGRBD | 91 | 91 | 1758 | 1837 | +79.65 | -0.379 | 0.705 | 0.077 |
| Bootstrap | 23 | V8_blood_IgGRBD | 91 | 91 | 562.1 | 594.4 | +32.28 | -0.260 | 0.795 | 0.055 |
| Bootstrap | 24 | V8a_blood_IgGRBD | 91 | 91 | 494.2 | 507.8 | +13.61 | -0.109 | 0.913 | 0.077 |
| Bootstrap | 25 | V8b_blood_IgGRBD | 91 | 91 | 2590 | 2700 | +110.2 | -0.587 | 0.558 | 0.077 |
| Bootstrap | 26 | V9_blood_IgGRBD | 91 | 91 | 1610 | 1459 | -151.8 | +0.654 | 0.514 | 0.066 |
| Bootstrap | 27 | V10_blood_IgGRBD | 91 | 91 | 1740 | 1574 | -166.3 | +0.648 | 0.518 | 0.110 |
| Bootstrap | 28 | V11_blood_IgGRBD | 91 | 91 | 2436 | 2342 | -94.02 | +0.423 | 0.673 | 0.066 |
| Bootstrap | 29 | V4_Saliva_IgGspike | 91 | 91 | 44.52 | 45.48 | +0.9611 | -0.219 | 0.827 | 0.044 |
| Bootstrap | 30 | V4_Saliva_IgGRBD | 91 | 91 | 34.03 | 34.73 | +0.6922 | -0.132 | 0.895 | 0.066 |
| Bootstrap | 31 | V5_Saliva_IgGspike | 91 | 91 | 145.8 | 151.4 | +5.645 | -0.346 | 0.73 | 0.066 |
| Bootstrap | 32 | V5_Saliva_IgGRBD | 91 | 91 | 153.5 | 157.8 | +4.285 | -0.210 | 0.834 | 0.044 |
| Bootstrap | 33 | V8_Saliva_IgGspike | 91 | 91 | 125.6 | 137.4 | +11.78 | -0.873 | 0.384 | 0.077 |
| Bootstrap | 34 | V8_Saliva_IgGRBD | 91 | 91 | 122.5 | 138.3 | +15.78 | -1.023 | 0.307 | 0.077 |
| Bootstrap | 35 | V8b_Saliva_IgGspike | 91 | 91 | 338.2 | 333.4 | -4.849 | +0.301 | 0.764 | 0.055 |
| Bootstrap | 36 | V8b_Saliva_IgGRBD | 91 | 91 | 398.6 | 393.1 | -5.437 | +0.261 | 0.794 | 0.066 |
| Bootstrap | 37 | V9_Saliva_IgGspike | 91 | 91 | 223.1 | 224.3 | +1.289 | -0.073 | 0.942 | 0.066 |
| Bootstrap | 38 | V9_Saliva_IgGRBD | 91 | 91 | 255.3 | 258.4 | +3.095 | -0.123 | 0.903 | 0.055 |

| Method | No. | Feature | n real | n synthetic | Real mean | Synthetic mean | Difference | Welch t | p | KS |
| --- | --- | --- | --- | --- | --- | --- | --- | --- | --- | --- |
| Bootstrap | 39 | V4_Saliva_IgAspike | 91 | 91 | 47.11 | 48.47 | +1.362 | -0.311 | 0.756 | 0.066 |
| Bootstrap | 40 | V4_Saliva_IgARBD | 91 | 91 | 43.1 | 43.99 | +0.8882 | -0.208 | 0.835 | 0.055 |
| Bootstrap | 41 | V5_Saliva_IgAspike | 91 | 91 | 84.74 | 88.62 | +3.879 | -0.408 | 0.684 | 0.044 |
| Bootstrap | 42 | V5_Saliva_IgARBD | 91 | 91 | 83.34 | 89.27 | +5.925 | -0.647 | 0.518 | 0.066 |
| Bootstrap | 43 | V8_Saliva_IgAspike | 91 | 91 | 51.38 | 54.19 | +2.807 | -0.495 | 0.621 | 0.088 |
| Bootstrap | 44 | V8_Saliva_IgARBD | 91 | 91 | 57.97 | 65.2 | +7.229 | -0.945 | 0.346 | 0.132 |
| Bootstrap | 45 | V8b_Saliva_IgAspike | 91 | 91 | 46.94 | 43.68 | -3.258 | +0.808 | 0.42 | 0.088 |
| Bootstrap | 46 | V8b_Saliva_IgARBD | 91 | 91 | 52.46 | 53.57 | +1.111 | -0.190 | 0.85 | 0.055 |
| Bootstrap | 47 | V9_Saliva_IgAspike | 91 | 91 | 27.17 | 26.5 | -0.6674 | +0.203 | 0.839 | 0.055 |
| Bootstrap | 48 | V9_Saliva_IgARBD | 91 | 91 | 26.6 | 26.71 | +0.1101 | -0.017 | 0.986 | 0.099 |
| Bootstrap | 49 | V8_IFNg | 91 | 91 | 31.17 | 32.46 | +1.294 | -0.268 | 0.789 | 0.088 |
| Bootstrap | 50 | V9_IFNg | 91 | 91 | 78.9 | 75.54 | -3.36 | +0.242 | 0.809 | 0.077 |
| Bootstrap | 51 | V8_IL2 | 91 | 91 | 130.2 | 128.7 | -1.486 | +0.079 | 0.937 | 0.055 |
| Bootstrap | 52 | V9_IL2 | 91 | 91 | 174.5 | 178.3 | +3.778 | -0.154 | 0.878 | 0.055 |
| Bootstrap | 53 | V9Dual | 91 | 91 | 19.91 | 19.29 | -0.611 | +0.164 | 0.87 | 0.066 |
| Bootstrap | 54 | V8Dual | 91 | 91 | 8.973 | 8.924 | -0.0489 | +0.029 | 0.977 | 0.088 |
| Bootstrap | 55 | IFNG_production | 91 | 91 | 0.09225 | 0.08842 | -0.003826 | +0.301 | 0.763 | 0.055 |
| Bootstrap | 56 | Il2_production | 91 | 91 | 0.2338 | 0.2262 | -0.00765 | +0.382 | 0.703 | 0.088 |
| Bootstrap | 57 | RATIO_CD4CD8 | 91 | 91 | 1.265 | 1.333 | +0.06758 | -0.446 | 0.656 | 0.088 |
| Bootstrap | 58 | V8Neut | 91 | 91 | 265.1 | 279.6 | +14.47 | -0.287 | 0.774 | 0.077 |
| Bootstrap | 59 | V9Neut | 91 | 91 | 935.7 | 893.9 | -41.79 | +0.297 | 0.767 | 0.066 |
| Bootstrap | 60 | V7_ACE2 | 91 | 91 | 334.4 | 331.4 | -2.976 | +0.059 | 0.953 | 0.077 |
| Bootstrap | 61 | V8_ACE2 | 91 | 91 | 136.2 | 125.7 | -10.44 | +0.545 | 0.587 | 0.077 |
| Bootstrap | 62 | V8b_ACE2 | 91 | 91 | 694.1 | 718.3 | +24.2 | -0.356 | 0.723 | 0.077 |
| Bootstrap | 63 | V9_ACE2 | 91 | 91 | 523.2 | 444.6 | -78.67 | +1.141 | 0.256 | 0.121 |
| Column-wise | 1 | spikeProduction_D1D2 | 91 | 91 | 3.141 | 3.152 | +0.01145 | -0.602 | 0.548 | 0.077 |
| Column-wise | 2 | spikeDecay_D1D2 | 91 | 91 | 0.02447 | 0.02441 | -6e-05 | +0.396 | 0.692 | 0.099 |
| Column-wise | 3 | spikeProduction_D3 | 91 | 91 | 0.6235 | 0.624 | +0.0005107 | -0.801 | 0.424 | 0.088 |
| Column-wise | 4 | spikeDecay_D3 | 91 | 91 | 0.008704 | 0.008685 | -1.838e-05 | +0.529 | 0.597 | 0.033 |
| Column-wise | 5 | RBDProduction_D1D2 | 91 | 91 | 2.405 | 2.413 | +0.00806 | -0.570 | 0.569 | 0.121 |
| Column-wise | 6 | RBDDecay_D1D2 | 91 | 91 | 0.02156 | 0.02152 | -4.037e-05 | +1.107 | 0.27 | 0.154 |
| Column-wise | 7 | RBDProduction_D3 | 91 | 91 | 0.817 | 0.8148 | -0.00216 | +0.622 | 0.535 | 0.055 |
| Column-wise | 8 | RBDDecay_D3 | 91 | 91 | 0.01138 | 0.01142 | +4.257e-05 | -1.302 | 0.195 | 0.099 |
| Column-wise | 9 | V1_blood_IgGspike | 91 | 91 | 6.896 | 8.064 | +1.168 | -0.913 | 0.363 | 0.066 |
| Column-wise | 10 | V4_blood_IgGspike | 91 | 91 | 203 | 228.7 | +25.69 | -0.536 | 0.592 | 0.066 |
| Column-wise | 11 | V4a_blood_IgGspike | 91 | 91 | 125.3 | 126.5 | +1.173 | -0.042 | 0.967 | 0.066 |
| Column-wise | 12 | V6_blood_IgGspike | 91 | 91 | 924.7 | 909.2 | -15.47 | +0.216 | 0.829 | 0.055 |
| Column-wise | 13 | V8_blood_IgGspike | 91 | 91 | 483.4 | 558.2 | +74.72 | -1.161 | 0.247 | 0.121 |

| Method | No. | Feature | n real | n synthetic | Real mean | Synthetic mean | Difference | Welch t | p | KS |
| --- | --- | --- | --- | --- | --- | --- | --- | --- | --- | --- |
| Column-wise | 14 | V8a_blood_IgGspike | 91 | 91 | 339.4 | 307.6 | -31.74 | +0.625 | 0.533 | 0.099 |
| Column-wise | 15 | V8b_blood_IgGspike | 91 | 91 | 1350 | 1363 | +13.73 | -0.310 | 0.757 | 0.055 |
| Column-wise | 16 | V9_blood_IgGspike | 91 | 91 | 1015 | 1050 | +34.83 | -0.483 | 0.63 | 0.066 |
| Column-wise | 17 | V10_blood_IgGspike | 91 | 91 | 962.7 | 992.2 | +29.42 | -0.365 | 0.715 | 0.055 |
| Column-wise | 18 | V11_blood_IgGspike | 91 | 91 | 1315 | 1339 | +24.48 | -0.518 | 0.605 | 0.077 |
| Column-wise | 19 | V1_blood_IgGRBD | 91 | 91 | 5.138 | 5.433 | +0.295 | -0.229 | 0.819 | 0.055 |
| Column-wise | 20 | V4_blood_IgGRBD | 91 | 91 | 226 | 165.7 | -60.31 | +0.914 | 0.362 | 0.055 |
| Column-wise | 21 | V4a_blood_IgGRBD | 91 | 91 | 159.5 | 215.4 | +55.94 | -0.646 | 0.519 | 0.099 |
| Column-wise | 22 | V6_blood_IgGRBD | 91 | 91 | 1758 | 1833 | +75.21 | -0.352 | 0.725 | 0.044 |
| Column-wise | 23 | V8_blood_IgGRBD | 91 | 91 | 562.1 | 719.2 | +157.1 | -1.098 | 0.274 | 0.066 |
| Column-wise | 24 | V8a_blood_IgGRBD | 91 | 91 | 494.2 | 522.5 | +28.34 | -0.210 | 0.834 | 0.077 |
| Column-wise | 25 | V8b_blood_IgGRBD | 91 | 91 | 2590 | 2684 | +94.86 | -0.486 | 0.628 | 0.088 |
| Column-wise | 26 | V9_blood_IgGRBD | 91 | 91 | 1610 | 1559 | -51.03 | +0.214 | 0.831 | 0.044 |
| Column-wise | 27 | V10_blood_IgGRBD | 91 | 91 | 1740 | 1933 | +192.7 | -0.757 | 0.45 | 0.121 |
| Column-wise | 28 | V11_blood_IgGRBD | 91 | 91 | 2436 | 2183 | -252.6 | +1.152 | 0.251 | 0.143 |
| Column-wise | 29 | V4_Saliva_IgGspike | 91 | 91 | 44.52 | 46.88 | +2.357 | -0.583 | 0.561 | 0.110 |
| Column-wise | 30 | V4_Saliva_IgGRBD | 91 | 91 | 34.03 | 30.26 | -3.777 | +0.711 | 0.478 | 0.088 |
| Column-wise | 31 | V5_Saliva_IgGspike | 91 | 91 | 145.8 | 170.2 | +24.43 | -1.474 | 0.142 | 0.121 |
| Column-wise | 32 | V5_Saliva_IgGRBD | 91 | 91 | 153.5 | 145 | -8.512 | +0.440 | 0.66 | 0.066 |
| Column-wise | 33 | V8_Saliva_IgGspike | 91 | 91 | 125.6 | 125.5 | -0.09937 | +0.007 | 0.994 | 0.066 |
| Column-wise | 34 | V8_Saliva_IgGRBD | 91 | 91 | 122.5 | 121.5 | -0.9894 | +0.066 | 0.948 | 0.066 |
| Column-wise | 35 | V8b_Saliva_IgGspike | 91 | 91 | 338.2 | 329.3 | -8.955 | +0.558 | 0.577 | 0.099 |
| Column-wise | 36 | V8b_Saliva_IgGRBD | 91 | 91 | 398.6 | 389.6 | -8.927 | +0.435 | 0.664 | 0.055 |
| Column-wise | 37 | V9_Saliva_IgGspike | 91 | 91 | 223.1 | 225.3 | +2.277 | -0.123 | 0.902 | 0.088 |
| Column-wise | 38 | V9_Saliva_IgGRBD | 91 | 91 | 255.3 | 270.6 | +15.31 | -0.602 | 0.548 | 0.088 |
| Column-wise | 39 | V4_Saliva_IgAspike | 91 | 91 | 47.11 | 43.63 | -3.474 | +0.827 | 0.409 | 0.066 |
| Column-wise | 40 | V4_Saliva_IgARBD | 91 | 91 | 43.1 | 48.49 | +5.394 | -1.198 | 0.233 | 0.110 |
| Column-wise | 41 | V5_Saliva_IgAspike | 91 | 91 | 84.74 | 81.42 | -3.313 | +0.373 | 0.709 | 0.088 |
| Column-wise | 42 | V5_Saliva_IgARBD | 91 | 91 | 83.34 | 91.78 | +8.444 | -0.859 | 0.391 | 0.099 |
| Column-wise | 43 | V8_Saliva_IgAspike | 91 | 91 | 51.38 | 50.15 | -1.229 | +0.227 | 0.821 | 0.066 |
| Column-wise | 44 | V8_Saliva_IgARBD | 91 | 91 | 57.97 | 49.49 | -8.473 | +1.141 | 0.255 | 0.132 |
| Column-wise | 45 | V8b_Saliva_IgAspike | 91 | 91 | 46.94 | 47.68 | +0.7407 | -0.166 | 0.869 | 0.055 |
| Column-wise | 46 | V8b_Saliva_IgARBD | 91 | 91 | 52.46 | 51.26 | -1.201 | +0.214 | 0.831 | 0.077 |
| Column-wise | 47 | V9_Saliva_IgAspike | 91 | 91 | 27.17 | 27.87 | +0.703 | -0.193 | 0.847 | 0.066 |
| Column-wise | 48 | V9_Saliva_IgARBD | 91 | 91 | 26.6 | 31.42 | +4.812 | -0.643 | 0.521 | 0.110 |
| Column-wise | 49 | V8_IFNg | 91 | 91 | 31.17 | 25.48 | -5.686 | +1.312 | 0.191 | 0.121 |
| Column-wise | 50 | V9_IFNg | 91 | 91 | 78.9 | 67.88 | -11.02 | +0.823 | 0.411 | 0.066 |
| Column-wise | 51 | V8_IL2 | 91 | 91 | 130.2 | 131 | +0.8381 | -0.044 | 0.965 | 0.044 |

| Method | No. | Feature | n real | n synthetic | Real mean | Synthetic mean | Difference | Welch t | p | KS |
| --- | --- | --- | --- | --- | --- | --- | --- | --- | --- | --- |
| Column-wise | 52 | V9_IL2 | 91 | 91 | 174.5 | 178.9 | +4.391 | -0.178 | 0.859 | 0.066 |
| Column-wise | 53 | V9Dual | 91 | 91 | 19.91 | 22.71 | +2.806 | -0.680 | 0.497 | 0.066 |
| Column-wise | 54 | V8Dual | 91 | 91 | 8.973 | 8.441 | -0.5321 | +0.323 | 0.747 | 0.066 |
| Column-wise | 55 | IFNG_production | 91 | 91 | 0.09225 | 0.09494 | +0.002687 | -0.206 | 0.837 | 0.044 |
| Column-wise | 56 | Il2_production | 91 | 91 | 0.2338 | 0.2337 | -0.0001503 | +0.008 | 0.994 | 0.055 |
| Column-wise | 57 | RATIO_CD4CD8 | 91 | 91 | 1.265 | 1.282 | +0.01668 | -0.119 | 0.905 | 0.066 |
| Column-wise | 58 | V8Neut | 91 | 91 | 265.1 | 299.7 | +34.56 | -0.672 | 0.502 | 0.066 |
| Column-wise | 59 | V9Neut | 91 | 91 | 935.7 | 1043 | +107.8 | -0.725 | 0.469 | 0.099 |
| Column-wise | 60 | V7_ACE2 | 91 | 91 | 334.4 | 358.8 | +24.36 | -0.460 | 0.646 | 0.088 |
| Column-wise | 61 | V8_ACE2 | 91 | 91 | 136.2 | 155.5 | +19.37 | -0.978 | 0.329 | 0.132 |
| Column-wise | 62 | V8b_ACE2 | 91 | 91 | 694.1 | 718.4 | +24.3 | -0.350 | 0.727 | 0.044 |
| Column-wise | 63 | V9_ACE2 | 91 | 91 | 523.2 | 550.6 | +27.36 | -0.366 | 0.715 | 0.110 |
| GMM | 1 | spikeProduction_D1D2 | 91 | 91 | 3.141 | 3.157 | +0.01615 | -0.911 | 0.364 | 0.154 |
| GMM | 2 | spikeDecay_D1D2 | 91 | 91 | 0.02447 | 0.02609 | +0.001617 | -1.686 | 0.0952 | 0.473 |
| GMM | 3 | spikeProduction_D3 | 91 | 91 | 0.6235 | 0.625 | +0.001483 | -1.237 | 0.219 | 0.286 |
| GMM | 4 | spikeDecay_D3 | 91 | 91 | 0.008704 | 0.008376 | -0.0003274 | +0.283 | 0.778 | 0.495 |
| GMM | 5 | RBDProduction_D1D2 | 91 | 91 | 2.405 | 2.407 | +0.002113 | -0.144 | 0.885 | 0.110 |
| GMM | 6 | RBDDecay_D1D2 | 91 | 91 | 0.02156 | 0.02198 | +0.0004186 | -0.382 | 0.703 | 0.495 |
| GMM | 7 | RBDProduction_D3 | 91 | 91 | 0.817 | 0.8146 | -0.002427 | +0.685 | 0.494 | 0.121 |
| GMM | 8 | RBDDecay_D3 | 91 | 91 | 0.01138 | 0.01148 | +0.0001079 | -0.091 | 0.928 | 0.516 |
| GMM | 9 | V1_blood_IgGspike | 91 | 91 | 6.896 | 6.527 | -0.3691 | +0.325 | 0.745 | 0.264 |
| GMM | 10 | V4_blood_IgGspike | 91 | 91 | 203 | 220.6 | +17.57 | -0.405 | 0.686 | 0.198 |
| GMM | 11 | V4a_blood_IgGspike | 91 | 91 | 125.3 | 136.9 | +11.6 | -0.409 | 0.683 | 0.110 |
| GMM | 12 | V6_blood_IgGspike | 91 | 91 | 924.7 | 908.5 | -16.17 | +0.214 | 0.831 | 0.132 |
| GMM | 13 | V8_blood_IgGspike | 91 | 91 | 483.4 | 508.1 | +24.62 | -0.363 | 0.717 | 0.176 |
| GMM | 14 | V8a_blood_IgGspike | 91 | 91 | 339.4 | 361.7 | +22.34 | -0.423 | 0.673 | 0.110 |
| GMM | 15 | V8b_blood_IgGspike | 91 | 91 | 1350 | 1316 | -33.45 | +0.718 | 0.474 | 0.297 |
| GMM | 16 | V9_blood_IgGspike | 91 | 91 | 1015 | 922.8 | -92.56 | +1.239 | 0.217 | 0.253 |
| GMM | 17 | V10_blood_IgGspike | 91 | 91 | 962.7 | 991.7 | +28.96 | -0.358 | 0.721 | 0.220 |
| GMM | 18 | V11_blood_IgGspike | 91 | 91 | 1315 | 1263 | -51.36 | +1.004 | 0.317 | 0.418 |
| GMM | 19 | V1_blood_IgGRBD | 91 | 91 | 5.138 | 4.572 | -0.5659 | +0.498 | 0.619 | 0.418 |
| GMM | 20 | V4_blood_IgGRBD | 91 | 91 | 226 | 251.9 | +25.85 | -0.314 | 0.754 | 0.319 |
| GMM | 21 | V4a_blood_IgGRBD | 91 | 91 | 159.5 | 178.4 | +18.95 | -0.270 | 0.788 | 0.264 |
| GMM | 22 | V6_blood_IgGRBD | 91 | 91 | 1758 | 1718 | -39.84 | +0.185 | 0.853 | 0.110 |
| GMM | 23 | V8_blood_IgGRBD | 91 | 91 | 562.1 | 650.8 | +88.67 | -0.691 | 0.49 | 0.132 |
| GMM | 24 | V8a_blood_IgGRBD | 91 | 91 | 494.2 | 563.1 | +68.94 | -0.545 | 0.587 | 0.154 |
| GMM | 25 | V8b_blood_IgGRBD | 91 | 91 | 2590 | 2343 | -246.2 | +1.206 | 0.23 | 0.132 |
| GMM | 26 | V9_blood_IgGRBD | 91 | 91 | 1610 | 1600 | -10.22 | +0.040 | 0.968 | 0.187 |

| Method | No. | Feature | n real | n synthetic | Real mean | Synthetic mean | Difference | Welch t | p | KS |
| --- | --- | --- | --- | --- | --- | --- | --- | --- | --- | --- |
| GMM | 27 | V10_blood_IgGRBD | 91 | 91 | 1740 | 1862 | +121.2 | -0.480 | 0.631 | 0.220 |
| GMM | 28 | V11_blood_IgGRBD | 91 | 91 | 2436 | 2243 | -192.3 | +0.850 | 0.397 | 0.176 |
| GMM | 29 | V4_Saliva_IgGspike | 91 | 91 | 44.52 | 44.66 | +0.1399 | -0.033 | 0.973 | 0.088 |
| GMM | 30 | V4_Saliva_IgGRBD | 91 | 91 | 34.03 | 35.31 | +1.275 | -0.225 | 0.822 | 0.099 |
| GMM | 31 | V5_Saliva_IgGspike | 91 | 91 | 145.8 | 145.6 | -0.1341 | +0.009 | 0.993 | 0.088 |
| GMM | 32 | V5_Saliva_IgGRBD | 91 | 91 | 153.5 | 159.1 | +5.553 | -0.276 | 0.783 | 0.110 |
| GMM | 33 | V8_Saliva_IgGspike | 91 | 91 | 125.6 | 112.2 | -13.4 | +0.994 | 0.322 | 0.132 |
| GMM | 34 | V8_Saliva_IgGRBD | 91 | 91 | 122.5 | 119.6 | -2.901 | +0.202 | 0.841 | 0.099 |
| GMM | 35 | V8b_Saliva_IgGspike | 91 | 91 | 338.2 | 319 | -19.18 | +1.305 | 0.194 | 0.209 |
| GMM | 36 | V8b_Saliva_IgGRBD | 91 | 91 | 398.6 | 376.8 | -21.76 | +1.107 | 0.27 | 0.176 |
| GMM | 37 | V9_Saliva_IgGspike | 91 | 91 | 223.1 | 207.9 | -15.2 | +0.904 | 0.367 | 0.132 |
| GMM | 38 | V9_Saliva_IgGRBD | 91 | 91 | 255.3 | 244.6 | -10.7 | +0.437 | 0.662 | 0.099 |
| GMM | 39 | V4_Saliva_IgAspike | 91 | 91 | 47.11 | 49.43 | +2.323 | -0.513 | 0.609 | 0.165 |
| GMM | 40 | V4_Saliva_IgARBD | 91 | 91 | 43.1 | 40.75 | -2.353 | +0.576 | 0.565 | 0.110 |
| GMM | 41 | V5_Saliva_IgAspike | 91 | 91 | 84.74 | 83.36 | -1.376 | +0.161 | 0.872 | 0.121 |
| GMM | 42 | V5_Saliva_IgARBD | 91 | 91 | 83.34 | 81.27 | -2.075 | +0.230 | 0.818 | 0.110 |
| GMM | 43 | V8_Saliva_IgAspike | 91 | 91 | 51.38 | 48.25 | -3.135 | +0.576 | 0.565 | 0.154 |
| GMM | 44 | V8_Saliva_IgARBD | 91 | 91 | 57.97 | 49.73 | -8.236 | +1.008 | 0.315 | 0.187 |
| GMM | 45 | V8b_Saliva_IgAspike | 91 | 91 | 46.94 | 47.16 | +0.2235 | -0.055 | 0.956 | 0.088 |
| GMM | 46 | V8b_Saliva_IgARBD | 91 | 91 | 52.46 | 47.6 | -4.854 | +0.858 | 0.392 | 0.121 |
| GMM | 47 | V9_Saliva_IgAspike | 91 | 91 | 27.17 | 27.48 | +0.3055 | -0.093 | 0.926 | 0.110 |
| GMM | 48 | V9_Saliva_IgARBD | 91 | 91 | 26.6 | 27.17 | +0.5615 | -0.088 | 0.93 | 0.187 |
| GMM | 49 | V8_IFNg | 91 | 91 | 31.17 | 24.75 | -6.422 | +1.487 | 0.139 | 0.154 |
| GMM | 50 | V9_IFNg | 91 | 91 | 78.9 | 59.04 | -19.86 | +1.550 | 0.123 | 0.176 |
| GMM | 51 | V8_IL2 | 91 | 91 | 130.2 | 109.5 | -20.7 | +1.201 | 0.231 | 0.154 |
| GMM | 52 | V9_IL2 | 91 | 91 | 174.5 | 146.1 | -28.41 | +1.217 | 0.225 | 0.154 |
| GMM | 53 | V9Dual | 91 | 91 | 19.91 | 14.71 | -5.2 | +1.557 | 0.121 | 0.176 |
| GMM | 54 | V8Dual | 91 | 91 | 8.973 | 6.836 | -2.137 | +1.437 | 0.153 | 0.165 |
| GMM | 55 | IFNG_production | 91 | 91 | 0.09225 | 0.08016 | -0.01209 | +0.995 | 0.321 | 0.132 |
| GMM | 56 | Il2_production | 91 | 91 | 0.2338 | 0.2184 | -0.01546 | +0.822 | 0.412 | 0.187 |
| GMM | 57 | RATIO_CD4CD8 | 91 | 91 | 1.265 | 1.244 | -0.021 | +0.141 | 0.888 | 0.110 |
| GMM | 58 | V8Neut | 91 | 91 | 265.1 | 259.3 | -5.773 | +0.121 | 0.904 | 0.165 |
| GMM | 59 | V9Neut | 91 | 91 | 935.7 | 891.4 | -44.34 | +0.294 | 0.769 | 0.220 |
| GMM | 60 | V7_ACE2 | 91 | 91 | 334.4 | 341.6 | +7.195 | -0.132 | 0.895 | 0.143 |
| GMM | 61 | V8_ACE2 | 91 | 91 | 136.2 | 153.6 | +17.41 | -0.744 | 0.458 | 0.297 |
| GMM | 62 | V8b_ACE2 | 91 | 91 | 694.1 | 633.2 | -60.93 | +0.791 | 0.43 | 0.143 |
| GMM | 63 | V9_ACE2 | 91 | 91 | 523.2 | 499.5 | -23.69 | +0.306 | 0.76 | 0.209 |
| SMOTE | 1 | spikeProduction_D1D2 | 91 | 91 | 3.141 | 3.157 | +0.01568 | -0.912 | 0.363 | 0.121 |

| Method | No. | Feature | n real | n synthetic | Real mean | Synthetic mean | Difference | Welch t | p | KS |
| --- | --- | --- | --- | --- | --- | --- | --- | --- | --- | --- |
| SMOTE | 2 | spikeDecay_D1D2 | 91 | 91 | 0.02447 | 0.02434 | -0.0001325 | +1.031 | 0.304 | 0.154 |
| SMOTE | 3 | spikeProduction_D3 | 91 | 91 | 0.6235 | 0.624 | +0.0004879 | -0.830 | 0.408 | 0.154 |
| SMOTE | 4 | spikeDecay_D3 | 91 | 91 | 0.008704 | 0.008694 | -9.774e-06 | +0.283 | 0.777 | 0.066 |
| SMOTE | 5 | RBDProduction_D1D2 | 91 | 91 | 2.405 | 2.402 | -0.002518 | +0.190 | 0.849 | 0.121 |
| SMOTE | 6 | RBDDecay_D1D2 | 91 | 91 | 0.02156 | 0.02156 | -3.905e-07 | +0.012 | 0.99 | 0.099 |
| SMOTE | 7 | RBDProduction_D3 | 91 | 91 | 0.817 | 0.8166 | -0.0003909 | +0.123 | 0.903 | 0.099 |
| SMOTE | 8 | RBDDecay_D3 | 91 | 91 | 0.01138 | 0.0114 | +2.491e-05 | -0.807 | 0.421 | 0.110 |
| SMOTE | 9 | V1_blood_IgGspike | 91 | 91 | 6.896 | 7.321 | +0.4253 | -0.396 | 0.692 | 0.154 |
| SMOTE | 10 | V4_blood_IgGspike | 91 | 91 | 203 | 224.3 | +21.3 | -0.515 | 0.607 | 0.110 |
| SMOTE | 11 | V4a_blood_IgGspike | 91 | 91 | 125.3 | 136.8 | +11.42 | -0.449 | 0.654 | 0.099 |
| SMOTE | 12 | V6_blood_IgGspike | 91 | 91 | 924.7 | 927 | +2.338 | -0.035 | 0.972 | 0.176 |
| SMOTE | 13 | V8_blood_IgGspike | 91 | 91 | 483.4 | 514.8 | +31.35 | -0.483 | 0.629 | 0.099 |
| SMOTE | 14 | V8a_blood_IgGspike | 91 | 91 | 339.4 | 343.1 | +3.723 | -0.076 | 0.94 | 0.099 |
| SMOTE | 15 | V8b_blood_IgGspike | 91 | 91 | 1350 | 1343 | -6.596 | +0.157 | 0.875 | 0.253 |
| SMOTE | 16 | V9_blood_IgGspike | 91 | 91 | 1015 | 974.6 | -40.68 | +0.582 | 0.561 | 0.132 |
| SMOTE | 17 | V10_blood_IgGspike | 91 | 91 | 962.7 | 970.9 | +8.137 | -0.104 | 0.917 | 0.121 |
| SMOTE | 18 | V11_blood_IgGspike | 91 | 91 | 1315 | 1280 | -34.46 | +0.711 | 0.478 | 0.121 |
| SMOTE | 19 | V1_blood_IgGRBD | 91 | 91 | 5.138 | 5.476 | +0.3384 | -0.342 | 0.733 | 0.330 |
| SMOTE | 20 | V4_blood_IgGRBD | 91 | 91 | 226 | 221.3 | -4.737 | +0.069 | 0.945 | 0.132 |
| SMOTE | 21 | V4a_blood_IgGRBD | 91 | 91 | 159.5 | 148.5 | -11.03 | +0.196 | 0.845 | 0.110 |
| SMOTE | 22 | V6_blood_IgGRBD | 91 | 91 | 1758 | 1806 | +48.55 | -0.231 | 0.817 | 0.121 |
| SMOTE | 23 | V8_blood_IgGRBD | 91 | 91 | 562.1 | 579.6 | +17.49 | -0.148 | 0.882 | 0.099 |
| SMOTE | 24 | V8a_blood_IgGRBD | 91 | 91 | 494.2 | 487.3 | -6.877 | +0.060 | 0.952 | 0.121 |
| SMOTE | 25 | V8b_blood_IgGRBD | 91 | 91 | 2590 | 2457 | -132.5 | +0.705 | 0.482 | 0.099 |
| SMOTE | 26 | V9_blood_IgGRBD | 91 | 91 | 1610 | 1487 | -123.2 | +0.523 | 0.601 | 0.088 |
| SMOTE | 27 | V10_blood_IgGRBD | 91 | 91 | 1740 | 1843 | +102.3 | -0.402 | 0.688 | 0.099 |
| SMOTE | 28 | V11_blood_IgGRBD | 91 | 91 | 2436 | 2332 | -103.4 | +0.461 | 0.646 | 0.099 |
| SMOTE | 29 | V4_Saliva_IgGspike | 91 | 91 | 44.52 | 43.48 | -1.037 | +0.256 | 0.798 | 0.066 |
| SMOTE | 30 | V4_Saliva_IgGRBD | 91 | 91 | 34.03 | 36.52 | +2.483 | -0.447 | 0.656 | 0.110 |
| SMOTE | 31 | V5_Saliva_IgGspike | 91 | 91 | 145.8 | 149 | +3.228 | -0.205 | 0.837 | 0.066 |
| SMOTE | 32 | V5_Saliva_IgGRBD | 91 | 91 | 153.5 | 160.6 | +7.072 | -0.343 | 0.732 | 0.066 |
| SMOTE | 33 | V8_Saliva_IgGspike | 91 | 91 | 125.6 | 124.7 | -0.9246 | +0.072 | 0.942 | 0.066 |
| SMOTE | 34 | V8_Saliva_IgGRBD | 91 | 91 | 122.5 | 122.3 | -0.1801 | +0.013 | 0.99 | 0.121 |
| SMOTE | 35 | V8b_Saliva_IgGspike | 91 | 91 | 338.2 | 329.3 | -8.953 | +0.628 | 0.531 | 0.143 |
| SMOTE | 36 | V8b_Saliva_IgGRBD | 91 | 91 | 398.6 | 399.5 | +0.9156 | -0.051 | 0.96 | 0.143 |
| SMOTE | 37 | V9_Saliva_IgGspike | 91 | 91 | 223.1 | 217.6 | -5.445 | +0.334 | 0.739 | 0.132 |
| SMOTE | 38 | V9_Saliva_IgGRBD | 91 | 91 | 255.3 | 246.4 | -8.91 | +0.368 | 0.713 | 0.110 |
| SMOTE | 39 | V4_Saliva_IgAspike | 91 | 91 | 47.11 | 46.12 | -0.9928 | +0.228 | 0.82 | 0.066 |

| Method | No. | Feature | n real | n synthetic | Real mean | Synthetic mean | Difference | Welch t | p | KS |
| --- | --- | --- | --- | --- | --- | --- | --- | --- | --- | --- |
| SMOTE | 40 | V4_Saliva_IgARBD | 91 | 91 | 43.1 | 42.34 | -0.7625 | +0.195 | 0.846 | 0.077 |
| SMOTE | 41 | V5_Saliva_IgAspike | 91 | 91 | 84.74 | 85.52 | +0.7832 | -0.089 | 0.929 | 0.055 |
| SMOTE | 42 | V5_Saliva_IgARBD | 91 | 91 | 83.34 | 82.18 | -1.161 | +0.130 | 0.897 | 0.077 |
| SMOTE | 43 | V8_Saliva_IgAspike | 91 | 91 | 51.38 | 51.82 | +0.4347 | -0.082 | 0.935 | 0.088 |
| SMOTE | 44 | V8_Saliva_IgARBD | 91 | 91 | 57.97 | 55.89 | -2.08 | +0.281 | 0.779 | 0.143 |
| SMOTE | 45 | V8b_Saliva_IgAspike | 91 | 91 | 46.94 | 44.65 | -2.283 | +0.557 | 0.578 | 0.066 |
| SMOTE | 46 | V8b_Saliva_IgARBD | 91 | 91 | 52.46 | 49.72 | -2.741 | +0.517 | 0.606 | 0.066 |
| SMOTE | 47 | V9_Saliva_IgAspike | 91 | 91 | 27.17 | 26.01 | -1.16 | +0.370 | 0.712 | 0.077 |
| SMOTE | 48 | V9_Saliva_IgARBD | 91 | 91 | 26.6 | 25.09 | -1.515 | +0.266 | 0.791 | 0.099 |
| SMOTE | 49 | V8_IFNg | 91 | 91 | 31.17 | 27.46 | -3.71 | +0.875 | 0.383 | 0.099 |
| SMOTE | 50 | V9_IFNg | 91 | 91 | 78.9 | 75.02 | -3.876 | +0.297 | 0.767 | 0.099 |
| SMOTE | 51 | V8_IL2 | 91 | 91 | 130.2 | 123.5 | -6.653 | +0.400 | 0.69 | 0.077 |
| SMOTE | 52 | V9_IL2 | 91 | 91 | 174.5 | 164.4 | -10.11 | +0.437 | 0.663 | 0.055 |
| SMOTE | 53 | V9Dual | 91 | 91 | 19.91 | 18.35 | -1.551 | +0.470 | 0.639 | 0.110 |
| SMOTE | 54 | V8Dual | 91 | 91 | 8.973 | 7.895 | -1.077 | +0.764 | 0.446 | 0.132 |
| SMOTE | 55 | IFNG_production | 91 | 91 | 0.09225 | 0.08774 | -0.004506 | +0.389 | 0.698 | 0.088 |
| SMOTE | 56 | II2_production | 91 | 91 | 0.2338 | 0.2314 | -0.002378 | +0.134 | 0.893 | 0.088 |
| SMOTE | 57 | RATIO_CD4CD8 | 91 | 91 | 1.265 | 1.288 | +0.02272 | -0.167 | 0.867 | 0.132 |
| SMOTE | 58 | V8Neut | 91 | 91 | 265.1 | 288.8 | +23.73 | -0.499 | 0.619 | 0.121 |
| SMOTE | 59 | V9Neut | 91 | 91 | 935.7 | 854.2 | -81.47 | +0.565 | 0.573 | 0.099 |
| SMOTE | 60 | V7_ACE2 | 91 | 91 | 334.4 | 358.7 | +24.25 | -0.444 | 0.657 | 0.132 |
| SMOTE | 61 | V8_ACE2 | 91 | 91 | 136.2 | 143.3 | +7.119 | -0.315 | 0.753 | 0.165 |
| SMOTE | 62 | V8b_ACE2 | 91 | 91 | 694.1 | 662.5 | -31.66 | +0.474 | 0.636 | 0.099 |
| SMOTE | 63 | V9_ACE2 | 91 | 91 | 523.2 | 481.4 | -41.84 | +0.573 | 0.568 | 0.110 |
| GMM-SMOTE | 1 | spikeProduction_D1D2 | 91 | 91 | 3.141 | 3.154 | +0.013 | -0.735 | 0.463 | 0.143 |
| GMM-SMOTE | 2 | spikeDecay_D1D2 | 91 | 91 | 0.02447 | 0.02437 | -0.0001021 | +0.746 | 0.457 | 0.143 |
| GMM-SMOTE | 3 | spikeProduction_D3 | 91 | 91 | 0.6235 | 0.6232 | -0.0003151 | +0.588 | 0.557 | 0.099 |
| GMM-SMOTE | 4 | spikeDecay_D3 | 91 | 91 | 0.008704 | 0.008728 | +2.424e-05 | -0.782 | 0.435 | 0.132 |
| GMM-SMOTE | 5 | RBDProduction_D1D2 | 91 | 91 | 2.405 | 2.412 | +0.007393 | -0.559 | 0.577 | 0.099 |
| GMM-SMOTE | 6 | RBDDecay_D1D2 | 91 | 91 | 0.02156 | 0.02156 | +1.385e-06 | -0.046 | 0.963 | 0.132 |
| GMM-SMOTE | 7 | RBDProduction_D3 | 91 | 91 | 0.817 | 0.8178 | +0.000777 | -0.249 | 0.803 | 0.088 |
| GMM-SMOTE | 8 | RBDDecay_D3 | 91 | 91 | 0.01138 | 0.01137 | -2.864e-06 | +0.097 | 0.923 | 0.121 |
| GMM-SMOTE | 9 | V1_blood_IgGspike | 91 | 91 | 6.896 | 7.585 | +0.689 | -0.650 | 0.516 | 0.165 |
| GMM-SMOTE | 10 | V4_blood_IgGspike | 91 | 91 | 203 | 224 | +20.99 | -0.522 | 0.602 | 0.176 |
| GMM-SMOTE | 11 | V4a_blood_IgGspike | 91 | 91 | 125.3 | 130.9 | +5.514 | -0.207 | 0.836 | 0.132 |
| GMM-SMOTE | 12 | V6_blood_IgGspike | 91 | 91 | 924.7 | 1035 | +110 | -1.661 | 0.0986 | 0.187 |
| GMM-SMOTE | 13 | V8_blood_IgGspike | 91 | 91 | 483.4 | 560.1 | +76.68 | -1.257 | 0.21 | 0.209 |
| GMM-SMOTE | 14 | V8a_blood_IgGspike | 91 | 91 | 339.4 | 368.7 | +29.34 | -0.615 | 0.539 | 0.165 |

| Method | No. | Feature | n real | n synthetic | Real mean | Synthetic mean | Difference | Welch t | p | KS |
| --- | --- | --- | --- | --- | --- | --- | --- | --- | --- | --- |
| GMM-SMOTE | 15 | V8b_blood_IgGspike | 91 | 91 | 1350 | 1394 | +44.66 | -1.118 | 0.265 | 0.220 |
| GMM-SMOTE | 16 | V9_blood_IgGspike | 91 | 91 | 1015 | 1096 | +80.19 | -1.192 | 0.235 | 0.121 |
| GMM-SMOTE | 17 | V10_blood_IgGspike | 91 | 91 | 962.7 | 930.4 | -32.31 | +0.428 | 0.669 | 0.154 |
| GMM-SMOTE | 18 | V11_blood_IgGspike | 91 | 91 | 1315 | 1328 | +13.44 | -0.302 | 0.763 | 0.143 |
| GMM-SMOTE | 19 | V1_blood_IgGRBD | 91 | 91 | 5.138 | 6.197 | +1.06 | -0.854 | 0.394 | 0.363 |
| GMM-SMOTE | 20 | V4_blood_IgGRBD | 91 | 91 | 226 | 248.4 | +22.32 | -0.289 | 0.773 | 0.187 |
| GMM-SMOTE | 21 | V4a_blood_IgGRBD | 91 | 91 | 159.5 | 166.9 | +7.469 | -0.112 | 0.911 | 0.132 |
| GMM-SMOTE | 22 | V6_blood_IgGRBD | 91 | 91 | 1758 | 2010 | +252.8 | -1.264 | 0.208 | 0.198 |
| GMM-SMOTE | 23 | V8_blood_IgGRBD | 91 | 91 | 562.1 | 598.4 | +36.26 | -0.336 | 0.737 | 0.198 |
| GMM-SMOTE | 24 | V8a_blood_IgGRBD | 91 | 91 | 494.2 | 473.6 | -20.59 | +0.193 | 0.848 | 0.165 |
| GMM-SMOTE | 25 | V8b_blood_IgGRBD | 91 | 91 | 2590 | 2742 | +152.3 | -0.843 | 0.4 | 0.154 |
| GMM-SMOTE | 26 | V9_blood_IgGRBD | 91 | 91 | 1610 | 1922 | +311.4 | -1.273 | 0.205 | 0.143 |
| GMM-SMOTE | 27 | V10_blood_IgGRBD | 91 | 91 | 1740 | 1507 | -233.1 | +0.961 | 0.338 | 0.121 |
| GMM-SMOTE | 28 | V11_blood_IgGRBD | 91 | 91 | 2436 | 2434 | -1.529 | +0.007 | 0.994 | 0.110 |
| GMM-SMOTE | 29 | V4_Saliva_IgGspike | 91 | 91 | 44.52 | 41.29 | -3.234 | +0.806 | 0.421 | 0.110 |
| GMM-SMOTE | 30 | V4_Saliva_IgGRBD | 91 | 91 | 34.03 | 34.74 | +0.7069 | -0.139 | 0.889 | 0.121 |
| GMM-SMOTE | 31 | V5_Saliva_IgGspike | 91 | 91 | 145.8 | 152.1 | +6.373 | -0.422 | 0.673 | 0.121 |
| GMM-SMOTE | 32 | V5_Saliva_IgGRBD | 91 | 91 | 153.5 | 160.9 | +7.319 | -0.379 | 0.705 | 0.143 |
| GMM-SMOTE | 33 | V8_Saliva_IgGspike | 91 | 91 | 125.6 | 127.4 | +1.722 | -0.132 | 0.895 | 0.055 |
| GMM-SMOTE | 34 | V8_Saliva_IgGRBD | 91 | 91 | 122.5 | 134.1 | +11.62 | -0.821 | 0.413 | 0.099 |
| GMM-SMOTE | 35 | V8b_Saliva_IgGspike | 91 | 91 | 338.2 | 341.1 | +2.891 | -0.204 | 0.839 | 0.077 |
| GMM-SMOTE | 36 | V8b_Saliva_IgGRBD | 91 | 91 | 398.6 | 396.2 | -2.421 | +0.130 | 0.897 | 0.099 |
| GMM-SMOTE | 37 | V9_Saliva_IgGspike | 91 | 91 | 223.1 | 241.2 | +18.18 | -1.047 | 0.296 | 0.121 |
| GMM-SMOTE | 38 | V9_Saliva_IgGRBD | 91 | 91 | 255.3 | 291 | +35.77 | -1.364 | 0.174 | 0.099 |
| GMM-SMOTE | 39 | V4_Saliva_IgAspike | 91 | 91 | 47.11 | 50.22 | +3.117 | -0.739 | 0.461 | 0.154 |
| GMM-SMOTE | 40 | V4_Saliva_IgARBD | 91 | 91 | 43.1 | 43.29 | +0.1914 | -0.048 | 0.962 | 0.055 |
| GMM-SMOTE | 41 | V5_Saliva_IgAspike | 91 | 91 | 84.74 | 82.25 | -2.488 | +0.317 | 0.752 | 0.088 |
| GMM-SMOTE | 42 | V5_Saliva_IgARBD | 91 | 91 | 83.34 | 82.08 | -1.257 | +0.152 | 0.879 | 0.088 |
| GMM-SMOTE | 43 | V8_Saliva_IgAspike | 91 | 91 | 51.38 | 47.51 | -3.876 | +0.857 | 0.393 | 0.110 |
| GMM-SMOTE | 44 | V8_Saliva_IgARBD | 91 | 91 | 57.97 | 59.83 | +1.862 | -0.235 | 0.815 | 0.077 |
| GMM-SMOTE | 45 | V8b_Saliva_IgAspike | 91 | 91 | 46.94 | 47.86 | +0.9268 | -0.227 | 0.821 | 0.077 |
| GMM-SMOTE | 46 | V8b_Saliva_IgARBD | 91 | 91 | 52.46 | 52.53 | +0.06822 | -0.012 | 0.99 | 0.044 |
| GMM-SMOTE | 47 | V9_Saliva_IgAspike | 91 | 91 | 27.17 | 32.11 | +4.942 | -1.341 | 0.182 | 0.121 |
| GMM-SMOTE | 48 | V9_Saliva_IgARBD | 91 | 91 | 26.6 | 35 | +8.394 | -1.209 | 0.228 | 0.132 |
| GMM-SMOTE | 49 | V8_IFNg | 91 | 91 | 31.17 | 29.64 | -1.528 | +0.336 | 0.737 | 0.099 |
| GMM-SMOTE | 50 | V9_IFNg | 91 | 91 | 78.9 | 74.86 | -4.041 | +0.296 | 0.767 | 0.110 |
| GMM-SMOTE | 51 | V8_IL2 | 91 | 91 | 130.2 | 134.1 | +3.96 | -0.226 | 0.821 | 0.088 |
| GMM-SMOTE | 52 | V9_IL2 | 91 | 91 | 174.5 | 188.3 | +13.82 | -0.587 | 0.558 | 0.121 |

| Method | No. | Feature | n real | n synthetic | Real mean | Synthetic mean | Difference | Welch t | p | KS |
| --- | --- | --- | --- | --- | --- | --- | --- | --- | --- | --- |
| GMM-SMOTE | 53 | V9Dual | 91 | 91 | 19.91 | 19.6 | -0.3052 | +0.086 | 0.931 | 0.121 |
| GMM-SMOTE | 54 | V8Dual | 91 | 91 | 8.973 | 8.198 | -0.7745 | +0.506 | 0.613 | 0.110 |
| GMM-SMOTE | 55 | IFNG_production | 91 | 91 | 0.09225 | 0.07755 | -0.0147 | +1.285 | 0.201 | 0.121 |
| GMM-SMOTE | 56 | IL2_production | 91 | 91 | 0.2338 | 0.2298 | -0.003975 | +0.235 | 0.814 | 0.077 |
| GMM-SMOTE | 57 | RATIO_CD4CD8 | 91 | 91 | 1.265 | 1.329 | +0.06339 | -0.477 | 0.634 | 0.154 |
| GMM-SMOTE | 58 | V8Neut | 91 | 91 | 265.1 | 292.9 | +27.82 | -0.584 | 0.56 | 0.143 |
| GMM-SMOTE | 59 | V9Neut | 91 | 91 | 935.7 | 1134 | +198.6 | -1.342 | 0.181 | 0.154 |
| GMM-SMOTE | 60 | V7_ACE2 | 91 | 91 | 334.4 | 387.8 | +53.38 | -1.002 | 0.318 | 0.154 |
| GMM-SMOTE | 61 | V8_ACE2 | 91 | 91 | 136.2 | 146.5 | +10.32 | -0.533 | 0.595 | 0.253 |
| GMM-SMOTE | 62 | V8b_ACE2 | 91 | 91 | 694.1 | 749 | +54.86 | -0.839 | 0.403 | 0.176 |
| GMM-SMOTE | 63 | V9_ACE2 | 91 | 91 | 523.2 | 618.4 | +95.13 | -1.238 | 0.217 | 0.132 |
| CVAE | 1 | spikeProduction_D1D2 | 91 | 91 | 3.141 | 3.154 | +0.01285 | -0.892 | 0.374 | 0.286 |
| CVAE | 2 | spikeDecay_D1D2 | 91 | 91 | 0.02447 | 0.02439 | -8.166e-05 | +0.777 | 0.439 | 0.297 |
| CVAE | 3 | spikeProduction_D3 | 91 | 91 | 0.6235 | 0.6238 | +0.0002667 | -0.583 | 0.561 | 0.286 |
| CVAE | 4 | spikeDecay_D3 | 91 | 91 | 0.008704 | 0.008713 | +9.169e-06 | -0.340 | 0.734 | 0.242 |
| CVAE | 5 | RBDProduction_D1D2 | 91 | 91 | 2.405 | 2.41 | +0.005742 | -0.502 | 0.617 | 0.198 |
| CVAE | 6 | RBDDecay_D1D2 | 91 | 91 | 0.02156 | 0.02156 | +3.404e-06 | -0.128 | 0.898 | 0.275 |
| CVAE | 7 | RBDProduction_D3 | 91 | 91 | 0.817 | 0.8176 | +0.0006512 | -0.258 | 0.797 | 0.264 |
| CVAE | 8 | RBDDecay_D3 | 91 | 91 | 0.01138 | 0.01135 | -2.792e-05 | +1.117 | 0.266 | 0.341 |
| CVAE | 9 | V1_blood_IgGspike | 91 | 91 | 6.896 | 6.866 | -0.03043 | +0.035 | 0.972 | 0.275 |
| CVAE | 10 | V4_blood_IgGspike | 91 | 91 | 203 | 167.7 | -35.29 | +1.071 | 0.286 | 0.121 |
| CVAE | 11 | V4a_blood_IgGspike | 91 | 91 | 125.3 | 94.83 | -30.52 | +1.379 | 0.17 | 0.165 |
| CVAE | 12 | V6_blood_IgGspike | 91 | 91 | 924.7 | 956.4 | +31.72 | -0.568 | 0.571 | 0.341 |
| CVAE | 13 | V8_blood_IgGspike | 91 | 91 | 483.4 | 443.8 | -39.65 | +0.797 | 0.427 | 0.352 |
| CVAE | 14 | V8a_blood_IgGspike | 91 | 91 | 339.4 | 291.8 | -47.59 | +1.231 | 0.221 | 0.264 |
| CVAE | 15 | V8b_blood_IgGspike | 91 | 91 | 1350 | 1386 | +36.18 | -1.069 | 0.287 | 0.407 |
| CVAE | 16 | V9_blood_IgGspike | 91 | 91 | 1015 | 999.5 | -15.8 | +0.282 | 0.779 | 0.396 |
| CVAE | 17 | V10_blood_IgGspike | 91 | 91 | 962.7 | 982.2 | +19.46 | -0.331 | 0.741 | 0.429 |
| CVAE | 18 | V11_blood_IgGspike | 91 | 91 | 1315 | 1335 | +20.62 | -0.562 | 0.575 | 0.593 |
| CVAE | 19 | V1_blood_IgGRBD | 91 | 91 | 5.138 | 5.953 | +0.8154 | -0.938 | 0.35 | 0.462 |
| CVAE | 20 | V4_blood_IgGRBD | 91 | 91 | 226 | 184.8 | -41.22 | +0.650 | 0.517 | 0.231 |
| CVAE | 21 | V4a_blood_IgGRBD | 91 | 91 | 159.5 | 96.33 | -63.14 | +1.143 | 0.255 | 0.418 |
| CVAE | 22 | V6_blood_IgGRBD | 91 | 91 | 1758 | 1670 | -87.84 | +0.531 | 0.596 | 0.264 |
| CVAE | 23 | V8_blood_IgGRBD | 91 | 91 | 562.1 | 427.9 | -134.3 | +1.491 | 0.139 | 0.231 |
| CVAE | 24 | V8a_blood_IgGRBD | 91 | 91 | 494.2 | 354.6 | -139.6 | +1.531 | 0.129 | 0.198 |
| CVAE | 25 | V8b_blood_IgGRBD | 91 | 91 | 2590 | 2774 | +184.8 | -1.280 | 0.203 | 0.385 |
| CVAE | 26 | V9_blood_IgGRBD | 91 | 91 | 1610 | 1440 | -170.1 | +0.877 | 0.382 | 0.297 |
| CVAE | 27 | V10_blood_IgGRBD | 91 | 91 | 1740 | 1712 | -27.89 | +0.149 | 0.882 | 0.484 |

| Method | No. | Feature | n real | n synthetic | Real mean | Synthetic mean | Difference | Welch t | p | KS |
| --- | --- | --- | --- | --- | --- | --- | --- | --- | --- | --- |
| CVAE | 28 | V11_blood_IgGRBD | 91 | 91 | 2436 | 2418 | -17.35 | +0.101 | 0.92 | 0.352 |
| CVAE | 29 | V4_Saliva_IgGspike | 91 | 91 | 44.52 | 45.8 | +1.276 | -0.387 | 0.699 | 0.308 |
| CVAE | 30 | V4_Saliva_IgGRBD | 91 | 91 | 34.03 | 28.57 | -5.464 | +1.320 | 0.189 | 0.275 |
| CVAE | 31 | V5_Saliva_IgGspike | 91 | 91 | 145.8 | 129.9 | -15.85 | +1.307 | 0.194 | 0.209 |
| CVAE | 32 | V5_Saliva_IgGRBD | 91 | 91 | 153.5 | 125.9 | -27.59 | +1.763 | 0.0804 | 0.143 |
| CVAE | 33 | V8_Saliva_IgGspike | 91 | 91 | 125.6 | 127.2 | +1.557 | -0.150 | 0.881 | 0.264 |
| CVAE | 34 | V8_Saliva_IgGRBD | 91 | 91 | 122.5 | 108.3 | -14.17 | +1.292 | 0.199 | 0.253 |
| CVAE | 35 | V8b_Saliva_IgGspike | 91 | 91 | 338.2 | 330.1 | -8.115 | +0.676 | 0.5 | 0.275 |
| CVAE | 36 | V8b_Saliva_IgGRBD | 91 | 91 | 398.6 | 382 | -16.53 | +1.087 | 0.279 | 0.264 |
| CVAE | 37 | V9_Saliva_IgGspike | 91 | 91 | 223.1 | 214.1 | -8.967 | +0.659 | 0.511 | 0.220 |
| CVAE | 38 | V9_Saliva_IgGRBD | 91 | 91 | 255.3 | 243.5 | -11.76 | +0.580 | 0.563 | 0.242 |
| CVAE | 39 | V4_Saliva_IgAspike | 91 | 91 | 47.11 | 47.58 | +0.4758 | -0.136 | 0.892 | 0.231 |
| CVAE | 40 | V4_Saliva_IgARBD | 91 | 91 | 43.1 | 37.79 | -5.312 | +1.646 | 0.102 | 0.264 |
| CVAE | 41 | V5_Saliva_IgAspike | 91 | 91 | 84.74 | 79.3 | -5.437 | +0.781 | 0.436 | 0.220 |
| CVAE | 42 | V5_Saliva_IgARBD | 91 | 91 | 83.34 | 69.02 | -14.32 | +2.045 | 0.043 | 0.264 |
| CVAE | 43 | V8_Saliva_IgAspike | 91 | 91 | 51.38 | 52.48 | +1.093 | -0.251 | 0.802 | 0.165 |
| CVAE | 44 | V8_Saliva_IgARBD | 91 | 91 | 57.97 | 47.44 | -10.53 | +1.767 | 0.0799 | 0.154 |
| CVAE | 45 | V8b_Saliva_IgAspike | 91 | 91 | 46.94 | 39.61 | -7.323 | +2.205 | 0.0293 | 0.231 |
| CVAE | 46 | V8b_Saliva_IgARBD | 91 | 91 | 52.46 | 47.24 | -5.219 | +1.153 | 0.251 | 0.154 |
| CVAE | 47 | V9_Saliva_IgAspike | 91 | 91 | 27.17 | 24.92 | -2.249 | +0.799 | 0.425 | 0.154 |
| CVAE | 48 | V9_Saliva_IgARBD | 91 | 91 | 26.6 | 21.04 | -5.56 | +1.083 | 0.281 | 0.132 |
| CVAE | 49 | V8_IFNg | 91 | 91 | 31.17 | 27.98 | -3.185 | +0.851 | 0.396 | 0.374 |
| CVAE | 50 | V9_IFNg | 91 | 91 | 78.9 | 73.58 | -5.322 | +0.474 | 0.636 | 0.341 |
| CVAE | 51 | V8_IL2 | 91 | 91 | 130.2 | 119.5 | -10.68 | +0.734 | 0.464 | 0.297 |
| CVAE | 52 | V9_IL2 | 91 | 91 | 174.5 | 165.6 | -8.865 | +0.461 | 0.645 | 0.286 |
| CVAE | 53 | V9Dual | 91 | 91 | 19.91 | 15.76 | -4.145 | +1.414 | 0.16 | 0.286 |
| CVAE | 54 | V8Dual | 91 | 91 | 8.973 | 7.665 | -1.308 | +1.015 | 0.312 | 0.187 |
| CVAE | 55 | IFNG_production | 91 | 91 | 0.09225 | 0.09138 | -0.0008717 | +0.085 | 0.933 | 0.297 |
| CVAE | 56 | Il2_production | 91 | 91 | 0.2338 | 0.2298 | -0.003967 | +0.265 | 0.791 | 0.242 |
| CVAE | 57 | RATIO_CD4CD8 | 91 | 91 | 1.265 | 1.132 | -0.1332 | +1.206 | 0.23 | 0.231 |
| CVAE | 58 | V8Neut | 91 | 91 | 265.1 | 241.9 | -23.19 | +0.607 | 0.545 | 0.308 |
| CVAE | 59 | V9Neut | 91 | 91 | 935.7 | 824.6 | -111.2 | +0.928 | 0.355 | 0.352 |
| CVAE | 60 | V7_ACE2 | 91 | 91 | 334.4 | 287.8 | -46.57 | +1.126 | 0.262 | 0.220 |
| CVAE | 61 | V8_ACE2 | 91 | 91 | 136.2 | 114.6 | -21.57 | +1.317 | 0.19 | 0.418 |
| CVAE | 62 | V8b_ACE2 | 91 | 91 | 694.1 | 682.2 | -11.91 | +0.222 | 0.824 | 0.330 |
| CVAE | 63 | V9_ACE2 | 91 | 91 | 523.2 | 511.6 | -11.66 | +0.189 | 0.85 | 0.341 |

**Table S3:** Complete feature-level marginal comparisons for Breast Cancer across all six synthesis methods. Difference is synthetic minus real; Welch p-values are unadjusted.

| Method | No. | Feature | n real | n synthetic | Real mean | Synthetic mean | Difference | Welch t | p | KS |
| --- | --- | --- | --- | --- | --- | --- | --- | --- | --- | --- |
| Bootstrap | 1 | mean radius | 569 | 569 | 14.13 | 14.09 | -0.03277 | +0.155 | 0.877 | 0.030 |
| Bootstrap | 2 | mean texture | 569 | 569 | 19.29 | 19.43 | +0.1417 | -0.548 | 0.584 | 0.030 |
| Bootstrap | 3 | mean perimeter | 569 | 569 | 91.97 | 91.71 | -0.2582 | +0.178 | 0.859 | 0.028 |
| Bootstrap | 4 | mean area | 569 | 569 | 654.9 | 652.8 | -2.121 | +0.101 | 0.92 | 0.032 |
| Bootstrap | 5 | mean smoothness | 569 | 569 | 0.09636 | 0.09588 | -0.0004755 | +0.580 | 0.562 | 0.025 |
| Bootstrap | 6 | mean compactness | 569 | 569 | 0.1043 | 0.1035 | -0.0008148 | +0.266 | 0.791 | 0.032 |
| Bootstrap | 7 | mean concavity | 569 | 569 | 0.0888 | 0.08675 | -0.002046 | +0.443 | 0.658 | 0.016 |
| Bootstrap | 8 | mean concave points | 569 | 569 | 0.04892 | 0.04786 | -0.001055 | +0.460 | 0.645 | 0.025 |
| Bootstrap | 9 | mean symmetry | 569 | 569 | 0.1812 | 0.1815 | +0.0003381 | -0.207 | 0.836 | 0.035 |
| Bootstrap | 10 | mean fractal dimension | 569 | 569 | 0.0628 | 0.06282 | +2.32e-05 | -0.057 | 0.955 | 0.025 |
| Bootstrap | 11 | radius error | 569 | 569 | 0.4052 | 0.3808 | -0.02435 | +1.597 | 0.111 | 0.062 |
| Bootstrap | 12 | texture error | 569 | 569 | 1.217 | 1.21 | -0.006474 | +0.204 | 0.839 | 0.028 |
| Bootstrap | 13 | perimeter error | 569 | 569 | 2.866 | 2.715 | -0.1513 | +1.374 | 0.17 | 0.046 |
| Bootstrap | 14 | area error | 569 | 569 | 40.34 | 37.09 | -3.248 | +1.362 | 0.173 | 0.049 |
| Bootstrap | 15 | smoothness error | 569 | 569 | 0.007041 | 0.00703 | -1.053e-05 | +0.059 | 0.953 | 0.018 |
| Bootstrap | 16 | compactness error | 569 | 569 | 0.02548 | 0.02506 | -0.0004193 | +0.406 | 0.685 | 0.021 |
| Bootstrap | 17 | concavity error | 569 | 569 | 0.03189 | 0.0316 | -0.0002922 | +0.170 | 0.865 | 0.023 |
| Bootstrap | 18 | concave points error | 569 | 569 | 0.0118 | 0.01161 | -0.0001828 | +0.510 | 0.61 | 0.023 |
| Bootstrap | 19 | symmetry error | 569 | 569 | 0.02054 | 0.0208 | +0.0002624 | -0.517 | 0.605 | 0.026 |
| Bootstrap | 20 | fractal dimension error | 569 | 569 | 0.003795 | 0.003712 | -8.241e-05 | +0.562 | 0.574 | 0.033 |
| Bootstrap | 21 | worst radius | 569 | 569 | 16.27 | 16.18 | -0.0903 | +0.314 | 0.754 | 0.049 |
| Bootstrap | 22 | worst texture | 569 | 569 | 25.68 | 25.95 | +0.273 | -0.737 | 0.462 | 0.035 |
| Bootstrap | 23 | worst perimeter | 569 | 569 | 107.3 | 106.7 | -0.5233 | +0.263 | 0.793 | 0.033 |
| Bootstrap | 24 | worst area | 569 | 569 | 880.6 | 872.9 | -7.725 | +0.228 | 0.819 | 0.044 |
| Bootstrap | 25 | worst smoothness | 569 | 569 | 0.1324 | 0.1326 | +0.0002048 | -0.151 | 0.88 | 0.019 |
| Bootstrap | 26 | worst compactness | 569 | 569 | 0.2543 | 0.2548 | +0.000493 | -0.054 | 0.957 | 0.042 |
| Bootstrap | 27 | worst concavity | 569 | 569 | 0.2722 | 0.2729 | +0.0006872 | -0.056 | 0.955 | 0.028 |
| Bootstrap | 28 | worst concave points | 569 | 569 | 0.1146 | 0.1147 | +8.552e-05 | -0.022 | 0.983 | 0.021 |
| Bootstrap | 29 | worst symmetry | 569 | 569 | 0.2901 | 0.2934 | +0.003364 | -0.923 | 0.356 | 0.040 |
| Bootstrap | 30 | worst fractal dimension | 569 | 569 | 0.08395 | 0.084 | +5.422e-05 | -0.053 | 0.958 | 0.063 |
| Column-wise | 1 | mean radius | 569 | 569 | 14.13 | 14.12 | -0.008337 | +0.041 | 0.967 | 0.030 |
| Column-wise | 2 | mean texture | 569 | 569 | 19.29 | 19.04 | -0.2537 | +0.991 | 0.322 | 0.037 |
| Column-wise | 3 | mean perimeter | 569 | 569 | 91.97 | 91.42 | -0.5464 | +0.380 | 0.704 | 0.025 |
| Column-wise | 4 | mean area | 569 | 569 | 654.9 | 650 | -4.88 | +0.236 | 0.814 | 0.028 |
| Column-wise | 5 | mean smoothness | 569 | 569 | 0.09636 | 0.09574 | -0.0006251 | +0.738 | 0.461 | 0.047 |
| Column-wise | 6 | mean compactness | 569 | 569 | 0.1043 | 0.1033 | -0.001045 | +0.334 | 0.739 | 0.032 |
| Column-wise | 7 | mean concavity | 569 | 569 | 0.0888 | 0.08559 | -0.003206 | +0.686 | 0.493 | 0.033 |
| Column-wise | 8 | mean concave points | 569 | 569 | 0.04892 | 0.04922 | +0.0003033 | -0.130 | 0.897 | 0.035 |

| Method | No. | Feature | n real | n synthetic | Real mean | Synthetic mean | Difference | Welch t | p | KS |
| --- | --- | --- | --- | --- | --- | --- | --- | --- | --- | --- |
| Column-wise | 9 | mean symmetry | 569 | 569 | 0.1812 | 0.1805 | -0.0007065 | +0.435 | 0.664 | 0.039 |
| Column-wise | 10 | mean fractal dimension | 569 | 569 | 0.0628 | 0.06294 | +0.000144 | -0.343 | 0.732 | 0.033 |
| Column-wise | 11 | radius error | 569 | 569 | 0.4052 | 0.3969 | -0.008304 | +0.498 | 0.619 | 0.032 |
| Column-wise | 12 | texture error | 569 | 569 | 1.217 | 1.23 | +0.01311 | -0.399 | 0.69 | 0.030 |
| Column-wise | 13 | perimeter error | 569 | 569 | 2.866 | 2.782 | -0.08367 | +0.717 | 0.473 | 0.037 |
| Column-wise | 14 | area error | 569 | 569 | 40.34 | 42.96 | +2.623 | -0.884 | 0.377 | 0.047 |
| Column-wise | 15 | smoothness error | 569 | 569 | 0.007041 | 0.00696 | -8.059e-05 | +0.441 | 0.659 | 0.035 |
| Column-wise | 16 | compactness error | 569 | 569 | 0.02548 | 0.02682 | +0.001343 | -1.272 | 0.204 | 0.058 |
| Column-wise | 17 | concavity error | 569 | 569 | 0.03189 | 0.03052 | -0.001377 | +0.825 | 0.41 | 0.033 |
| Column-wise | 18 | concave points error | 569 | 569 | 0.0118 | 0.01179 | -2.023e-06 | +0.005 | 0.996 | 0.039 |
| Column-wise | 19 | symmetry error | 569 | 569 | 0.02054 | 0.02025 | -0.0002876 | +0.598 | 0.55 | 0.032 |
| Column-wise | 20 | fractal dimension error | 569 | 569 | 0.003795 | 0.003731 | -6.433e-05 | +0.412 | 0.681 | 0.039 |
| Column-wise | 21 | worst radius | 569 | 569 | 16.27 | 16.12 | -0.1534 | +0.542 | 0.588 | 0.028 |
| Column-wise | 22 | worst texture | 569 | 569 | 25.68 | 26 | +0.3253 | -0.872 | 0.383 | 0.033 |
| Column-wise | 23 | worst perimeter | 569 | 569 | 107.3 | 106.5 | -0.7362 | +0.364 | 0.716 | 0.037 |
| Column-wise | 24 | worst area | 569 | 569 | 880.6 | 884.4 | +3.85 | -0.114 | 0.909 | 0.026 |
| Column-wise | 25 | worst smoothness | 569 | 569 | 0.1324 | 0.1334 | +0.001032 | -0.758 | 0.449 | 0.032 |
| Column-wise | 26 | worst compactness | 569 | 569 | 0.2543 | 0.2538 | -0.0005093 | +0.057 | 0.955 | 0.051 |
| Column-wise | 27 | worst concavity | 569 | 569 | 0.2722 | 0.2725 | +0.0002705 | -0.022 | 0.983 | 0.025 |
| Column-wise | 28 | worst concave points | 569 | 569 | 0.1146 | 0.1123 | -0.002311 | +0.603 | 0.547 | 0.033 |
| Column-wise | 29 | worst symmetry | 569 | 569 | 0.2901 | 0.2882 | -0.001918 | +0.524 | 0.6 | 0.035 |
| Column-wise | 30 | worst fractal dimension | 569 | 569 | 0.08395 | 0.08427 | +0.0003251 | -0.303 | 0.762 | 0.037 |
| GMM | 1 | mean radius | 569 | 569 | 14.13 | 14.02 | -0.1062 | +0.500 | 0.617 | 0.056 |
| GMM | 2 | mean texture | 569 | 569 | 19.29 | 19.24 | -0.04491 | +0.175 | 0.861 | 0.058 |
| GMM | 3 | mean perimeter | 569 | 569 | 91.97 | 91.27 | -0.6961 | +0.475 | 0.635 | 0.053 |
| GMM | 4 | mean area | 569 | 569 | 654.9 | 648.4 | -6.513 | +0.307 | 0.759 | 0.063 |
| GMM | 5 | mean smoothness | 569 | 569 | 0.09636 | 0.09549 | -0.0008737 | +0.921 | 0.357 | 0.100 |
| GMM | 6 | mean compactness | 569 | 569 | 0.1043 | 0.1034 | -0.0009312 | +0.297 | 0.767 | 0.056 |
| GMM | 7 | mean concavity | 569 | 569 | 0.0888 | 0.08822 | -0.0005746 | +0.123 | 0.902 | 0.111 |
| GMM | 8 | mean concave points | 569 | 569 | 0.04892 | 0.04737 | -0.00155 | +0.655 | 0.512 | 0.067 |
| GMM | 9 | mean symmetry | 569 | 569 | 0.1812 | 0.1808 | -0.0003164 | +0.185 | 0.853 | 0.056 |
| GMM | 10 | mean fractal dimension | 569 | 569 | 0.0628 | 0.06258 | -0.0002195 | +0.367 | 0.713 | 0.200 |
| GMM | 11 | radius error | 569 | 569 | 0.4052 | 0.3986 | -0.00656 | +0.395 | 0.693 | 0.076 |
| GMM | 12 | texture error | 569 | 569 | 1.217 | 1.182 | -0.03496 | +1.067 | 0.286 | 0.067 |
| GMM | 13 | perimeter error | 569 | 569 | 2.866 | 2.857 | -0.009159 | +0.076 | 0.939 | 0.083 |
| GMM | 14 | area error | 569 | 569 | 40.34 | 39.99 | -0.3437 | +0.126 | 0.9 | 0.105 |
| GMM | 15 | smoothness error | 569 | 569 | 0.007041 | 0.007702 | +0.0006613 | -1.367 | 0.172 | 0.346 |
| GMM | 16 | compactness error | 569 | 569 | 0.02548 | 0.02526 | -0.000223 | +0.197 | 0.844 | 0.137 |

| Method | No. | Feature | n real | n synthetic | Real mean | Synthetic mean | Difference | Welch t | p | KS |
| --- | --- | --- | --- | --- | --- | --- | --- | --- | --- | --- |
| GMM | 17 | concavity error | 569 | 569 | 0.03189 | 0.03253 | +0.0006316 | -0.348 | 0.728 | 0.120 |
| GMM | 18 | concave points error | 569 | 569 | 0.0118 | 0.01116 | -0.0006405 | +1.120 | 0.263 | 0.243 |
| GMM | 19 | symmetry error | 569 | 569 | 0.02054 | 0.02143 | +0.000885 | -1.404 | 0.161 | 0.192 |
| GMM | 20 | fractal dimension error | 569 | 569 | 0.003795 | 0.003378 | -0.0004172 | +0.910 | 0.363 | 0.420 |
| GMM | 21 | worst radius | 569 | 569 | 16.27 | 16.16 | -0.1066 | +0.366 | 0.714 | 0.054 |
| GMM | 22 | worst texture | 569 | 569 | 25.68 | 25.59 | -0.08444 | +0.232 | 0.816 | 0.047 |
| GMM | 23 | worst perimeter | 569 | 569 | 107.3 | 106.8 | -0.4887 | +0.241 | 0.81 | 0.058 |
| GMM | 24 | worst area | 569 | 569 | 880.6 | 873.1 | -7.49 | +0.218 | 0.827 | 0.060 |
| GMM | 25 | worst smoothness | 569 | 569 | 0.1324 | 0.1323 | -8.853e-05 | +0.065 | 0.948 | 0.044 |
| GMM | 26 | worst compactness | 569 | 569 | 0.2543 | 0.2549 | +0.0005983 | -0.065 | 0.948 | 0.054 |
| GMM | 27 | worst concavity | 569 | 569 | 0.2722 | 0.2767 | +0.004518 | -0.372 | 0.71 | 0.100 |
| GMM | 28 | worst concave points | 569 | 569 | 0.1146 | 0.1139 | -0.0007422 | +0.189 | 0.85 | 0.039 |
| GMM | 29 | worst symmetry | 569 | 569 | 0.2901 | 0.2926 | +0.002548 | -0.681 | 0.496 | 0.067 |
| GMM | 30 | worst fractal dimension | 569 | 569 | 0.08395 | 0.08352 | -0.0004303 | +0.382 | 0.703 | 0.095 |
| SMOTE | 1 | mean radius | 569 | 569 | 14.13 | 14.15 | +0.0206 | -0.099 | 0.921 | 0.021 |
| SMOTE | 2 | mean texture | 569 | 569 | 19.29 | 19.12 | -0.1659 | +0.687 | 0.492 | 0.037 |
| SMOTE | 3 | mean perimeter | 569 | 569 | 91.97 | 92.04 | +0.07009 | -0.049 | 0.961 | 0.019 |
| SMOTE | 4 | mean area | 569 | 569 | 654.9 | 656.2 | +1.329 | -0.065 | 0.949 | 0.025 |
| SMOTE | 5 | mean smoothness | 569 | 569 | 0.09636 | 0.0959 | -0.0004621 | +0.596 | 0.551 | 0.060 |
| SMOTE | 6 | mean compactness | 569 | 569 | 0.1043 | 0.1029 | -0.001467 | +0.498 | 0.618 | 0.060 |
| SMOTE | 7 | mean concavity | 569 | 569 | 0.0888 | 0.08637 | -0.002426 | +0.543 | 0.587 | 0.065 |
| SMOTE | 8 | mean concave points | 569 | 569 | 0.04892 | 0.04767 | -0.001244 | +0.562 | 0.574 | 0.046 |
| SMOTE | 9 | mean symmetry | 569 | 569 | 0.1812 | 0.1804 | -0.0007565 | +0.514 | 0.608 | 0.069 |
| SMOTE | 10 | mean fractal dimension | 569 | 569 | 0.0628 | 0.06253 | -0.000267 | +0.669 | 0.504 | 0.053 |
| SMOTE | 11 | radius error | 569 | 569 | 0.4052 | 0.3944 | -0.01073 | +0.692 | 0.489 | 0.054 |
| SMOTE | 12 | texture error | 569 | 569 | 1.217 | 1.184 | -0.03287 | +1.108 | 0.268 | 0.069 |
| SMOTE | 13 | perimeter error | 569 | 569 | 2.866 | 2.775 | -0.09131 | +0.818 | 0.414 | 0.053 |
| SMOTE | 14 | area error | 569 | 569 | 40.34 | 39.27 | -1.067 | +0.414 | 0.679 | 0.053 |
| SMOTE | 15 | smoothness error | 569 | 569 | 0.007041 | 0.006903 | -0.0001382 | +0.860 | 0.39 | 0.042 |
| SMOTE | 16 | compactness error | 569 | 569 | 0.02548 | 0.02484 | -0.0006429 | +0.664 | 0.507 | 0.084 |
| SMOTE | 17 | concavity error | 569 | 569 | 0.03189 | 0.03081 | -0.001083 | +0.675 | 0.5 | 0.079 |
| SMOTE | 18 | concave points error | 569 | 569 | 0.0118 | 0.01142 | -0.0003765 | +1.131 | 0.258 | 0.054 |
| SMOTE | 19 | symmetry error | 569 | 569 | 0.02054 | 0.02025 | -0.000288 | +0.633 | 0.527 | 0.053 |
| SMOTE | 20 | fractal dimension error | 569 | 569 | 0.003795 | 0.003711 | -8.406e-05 | +0.571 | 0.568 | 0.074 |
| SMOTE | 21 | worst radius | 569 | 569 | 16.27 | 16.33 | +0.06428 | -0.223 | 0.824 | 0.018 |
| SMOTE | 22 | worst texture | 569 | 569 | 25.68 | 25.59 | -0.08522 | +0.245 | 0.807 | 0.042 |
| SMOTE | 23 | worst perimeter | 569 | 569 | 107.3 | 107.6 | +0.3287 | -0.164 | 0.869 | 0.023 |
| SMOTE | 24 | worst area | 569 | 569 | 880.6 | 889.2 | +8.569 | -0.251 | 0.802 | 0.016 |

| Method | No. | Feature | n real | n synthetic | Real mean | Synthetic mean | Difference | Welch t | p | KS |
| --- | --- | --- | --- | --- | --- | --- | --- | --- | --- | --- |
| SMOTE | 25 | worst smoothness | 569 | 569 | 0.1324 | 0.1325 | +0.0001814 | -0.141 | 0.888 | 0.044 |
| SMOTE | 26 | worst compactness | 569 | 569 | 0.2543 | 0.2558 | +0.001485 | -0.166 | 0.868 | 0.054 |
| SMOTE | 27 | worst concavity | 569 | 569 | 0.2722 | 0.272 | -0.0002189 | +0.019 | 0.985 | 0.054 |
| SMOTE | 28 | worst concave points | 569 | 569 | 0.1146 | 0.1142 | -0.0003908 | +0.103 | 0.918 | 0.040 |
| SMOTE | 29 | worst symmetry | 569 | 569 | 0.2901 | 0.2913 | +0.001195 | -0.351 | 0.726 | 0.074 |
| SMOTE | 30 | worst fractal dimension | 569 | 569 | 0.08395 | 0.0839 | -4.614e-05 | +0.046 | 0.963 | 0.054 |
| GMM-SMOTE | 1 | mean radius | 569 | 569 | 14.13 | 14.22 | +0.08863 | -0.420 | 0.675 | 0.026 |
| GMM-SMOTE | 2 | mean texture | 569 | 569 | 19.29 | 19.27 | -0.0236 | +0.099 | 0.921 | 0.051 |
| GMM-SMOTE | 3 | mean perimeter | 569 | 569 | 91.97 | 92.47 | +0.5038 | -0.347 | 0.729 | 0.030 |
| GMM-SMOTE | 4 | mean area | 569 | 569 | 654.9 | 664.2 | +9.315 | -0.441 | 0.659 | 0.028 |
| GMM-SMOTE | 5 | mean smoothness | 569 | 569 | 0.09636 | 0.09562 | -0.0007398 | +0.959 | 0.338 | 0.070 |
| GMM-SMOTE | 6 | mean compactness | 569 | 569 | 0.1043 | 0.1021 | -0.002232 | +0.752 | 0.452 | 0.054 |
| GMM-SMOTE | 7 | mean concavity | 569 | 569 | 0.0888 | 0.08567 | -0.003132 | +0.703 | 0.482 | 0.047 |
| GMM-SMOTE | 8 | mean concave points | 569 | 569 | 0.04892 | 0.04825 | -0.0006715 | +0.301 | 0.764 | 0.053 |
| GMM-SMOTE | 9 | mean symmetry | 569 | 569 | 0.1812 | 0.1798 | -0.00135 | +0.888 | 0.375 | 0.056 |
| GMM-SMOTE | 10 | mean fractal dimension | 569 | 569 | 0.0628 | 0.06217 | -0.0006274 | +1.597 | 0.111 | 0.063 |
| GMM-SMOTE | 11 | radius error | 569 | 569 | 0.4052 | 0.403 | -0.002212 | +0.140 | 0.889 | 0.049 |
| GMM-SMOTE | 12 | texture error | 569 | 569 | 1.217 | 1.201 | -0.01542 | +0.516 | 0.606 | 0.069 |
| GMM-SMOTE | 13 | perimeter error | 569 | 569 | 2.866 | 2.832 | -0.03399 | +0.294 | 0.769 | 0.044 |
| GMM-SMOTE | 14 | area error | 569 | 569 | 40.34 | 40.1 | -0.2338 | +0.091 | 0.928 | 0.032 |
| GMM-SMOTE | 15 | smoothness error | 569 | 569 | 0.007041 | 0.006998 | -4.296e-05 | +0.257 | 0.798 | 0.039 |
| GMM-SMOTE | 16 | compactness error | 569 | 569 | 0.02548 | 0.02492 | -0.0005626 | +0.577 | 0.564 | 0.063 |
| GMM-SMOTE | 17 | concavity error | 569 | 569 | 0.03189 | 0.03058 | -0.001309 | +0.850 | 0.396 | 0.051 |
| GMM-SMOTE | 18 | concave points error | 569 | 569 | 0.0118 | 0.01181 | +1.589e-05 | -0.046 | 0.963 | 0.046 |
| GMM-SMOTE | 19 | symmetry error | 569 | 569 | 0.02054 | 0.02042 | -0.0001239 | +0.268 | 0.788 | 0.060 |
| GMM-SMOTE | 20 | fractal dimension error | 569 | 569 | 0.003795 | 0.003608 | -0.0001874 | +1.362 | 0.173 | 0.062 |
| GMM-SMOTE | 21 | worst radius | 569 | 569 | 16.27 | 16.34 | +0.07212 | -0.249 | 0.803 | 0.026 |
| GMM-SMOTE | 22 | worst texture | 569 | 569 | 25.68 | 25.66 | -0.01733 | +0.051 | 0.959 | 0.065 |
| GMM-SMOTE | 23 | worst perimeter | 569 | 569 | 107.3 | 107.6 | +0.3003 | -0.150 | 0.881 | 0.026 |
| GMM-SMOTE | 24 | worst area | 569 | 569 | 880.6 | 888.8 | +8.244 | -0.242 | 0.809 | 0.028 |
| GMM-SMOTE | 25 | worst smoothness | 569 | 569 | 0.1324 | 0.1315 | -0.00084 | +0.674 | 0.501 | 0.069 |
| GMM-SMOTE | 26 | worst compactness | 569 | 569 | 0.2543 | 0.2491 | -0.005138 | +0.575 | 0.566 | 0.054 |
| GMM-SMOTE | 27 | worst concavity | 569 | 569 | 0.2722 | 0.2633 | -0.008931 | +0.766 | 0.444 | 0.054 |
| GMM-SMOTE | 28 | worst concave points | 569 | 569 | 0.1146 | 0.1137 | -0.0009352 | +0.247 | 0.805 | 0.044 |
| GMM-SMOTE | 29 | worst symmetry | 569 | 569 | 0.2901 | 0.2892 | -0.0009066 | +0.261 | 0.794 | 0.046 |
| GMM-SMOTE | 30 | worst fractal dimension | 569 | 569 | 0.08395 | 0.08257 | -0.00138 | +1.368 | 0.172 | 0.074 |
| CVAE | 1 | mean radius | 569 | 569 | 14.13 | 13.92 | -0.2037 | +1.019 | 0.309 | 0.049 |
| CVAE | 2 | mean texture | 569 | 569 | 19.29 | 19.2 | -0.08935 | +0.374 | 0.709 | 0.058 |

| Method | No. | Feature | n real | n synthetic | Real mean | Synthetic mean | Difference | Welch t | p | KS |
| --- | --- | --- | --- | --- | --- | --- | --- | --- | --- | --- |
| CVAE | 3 | mean perimeter | 569 | 569 | 91.97 | 90.77 | -1.195 | +0.864 | 0.388 | 0.054 |
| CVAE | 4 | mean area | 569 | 569 | 654.9 | 635 | -19.92 | +1.012 | 0.312 | 0.044 |
| CVAE | 5 | mean smoothness | 569 | 569 | 0.09636 | 0.09666 | +0.000302 | -0.401 | 0.689 | 0.086 |
| CVAE | 6 | mean compactness | 569 | 569 | 0.1043 | 0.1048 | +0.0004246 | -0.147 | 0.884 | 0.086 |
| CVAE | 7 | mean concavity | 569 | 569 | 0.0888 | 0.08581 | -0.002993 | +0.676 | 0.499 | 0.047 |
| CVAE | 8 | mean concave points | 569 | 569 | 0.04892 | 0.0483 | -0.0006179 | +0.282 | 0.778 | 0.051 |
| CVAE | 9 | mean symmetry | 569 | 569 | 0.1812 | 0.1822 | +0.0009939 | -0.676 | 0.499 | 0.070 |
| CVAE | 10 | mean fractal dimension | 569 | 569 | 0.0628 | 0.06292 | +0.0001251 | -0.336 | 0.737 | 0.097 |
| CVAE | 11 | radius error | 569 | 569 | 0.4052 | 0.4027 | -0.002461 | +0.166 | 0.868 | 0.065 |
| CVAE | 12 | texture error | 569 | 569 | 1.217 | 1.254 | +0.03666 | -1.200 | 0.23 | 0.090 |
| CVAE | 13 | perimeter error | 569 | 569 | 2.866 | 2.871 | +0.004924 | -0.046 | 0.964 | 0.093 |
| CVAE | 14 | area error | 569 | 569 | 40.34 | 39.7 | -0.6338 | +0.265 | 0.791 | 0.056 |
| CVAE | 15 | smoothness error | 569 | 569 | 0.007041 | 0.007185 | +0.0001444 | -0.924 | 0.356 | 0.120 |
| CVAE | 16 | compactness error | 569 | 569 | 0.02548 | 0.02576 | +0.0002792 | -0.290 | 0.772 | 0.102 |
| CVAE | 17 | concavity error | 569 | 569 | 0.03189 | 0.03136 | -0.0005349 | +0.355 | 0.722 | 0.114 |
| CVAE | 18 | concave points error | 569 | 569 | 0.0118 | 0.01225 | +0.0004504 | -1.358 | 0.175 | 0.100 |
| CVAE | 19 | symmetry error | 569 | 569 | 0.02054 | 0.02115 | +0.0006053 | -1.387 | 0.166 | 0.132 |
| CVAE | 20 | fractal dimension error | 569 | 569 | 0.003795 | 0.003657 | -0.0001377 | +1.028 | 0.304 | 0.083 |
| CVAE | 21 | worst radius | 569 | 569 | 16.27 | 16.03 | -0.237 | +0.848 | 0.397 | 0.069 |
| CVAE | 22 | worst texture | 569 | 569 | 25.68 | 25.61 | -0.07006 | +0.202 | 0.84 | 0.058 |
| CVAE | 23 | worst perimeter | 569 | 569 | 107.3 | 105.6 | -1.703 | +0.878 | 0.38 | 0.062 |
| CVAE | 24 | worst area | 569 | 569 | 880.6 | 857.2 | -23.36 | +0.717 | 0.474 | 0.072 |
| CVAE | 25 | worst smoothness | 569 | 569 | 0.1324 | 0.1329 | +0.0005566 | -0.452 | 0.652 | 0.063 |
| CVAE | 26 | worst compactness | 569 | 569 | 0.2543 | 0.246 | -0.008308 | +0.972 | 0.331 | 0.042 |
| CVAE | 27 | worst concavity | 569 | 569 | 0.2722 | 0.2622 | -0.01 | +0.882 | 0.378 | 0.056 |
| CVAE | 28 | worst concave points | 569 | 569 | 0.1146 | 0.1148 | +0.0001736 | -0.047 | 0.963 | 0.051 |
| CVAE | 29 | worst symmetry | 569 | 569 | 0.2901 | 0.2917 | +0.001663 | -0.506 | 0.613 | 0.083 |
| CVAE | 30 | worst fractal dimension | 569 | 569 | 0.08395 | 0.08277 | -0.001172 | +1.229 | 0.219 | 0.067 |

**Table S4:** Complete feature-level marginal comparisons for Diabetes across all six synthesis methods. Difference is synthetic minus real; Welch p-values are unadjusted.

| Method | No. | Feature | n real | n synthetic | Real mean | Synthetic mean | Difference | Welch t | p | KS |
| --- | --- | --- | --- | --- | --- | --- | --- | --- | --- | --- |
| Bootstrap | 1 | preg | 768 | 768 | 3.845 | 3.717 | -0.1276 | +0.751 | 0.453 | 0.030 |
| Bootstrap | 2 | plas | 768 | 768 | 120.9 | 119.9 | -0.9896 | +0.605 | 0.545 | 0.020 |
| Bootstrap | 3 | pres | 768 | 768 | 69.11 | 69.49 | +0.3854 | -0.405 | 0.686 | 0.018 |
| Bootstrap | 4 | skin | 768 | 768 | 20.54 | 21.32 | +0.7839 | -0.956 | 0.339 | 0.039 |
| Bootstrap | 5 | insu | 768 | 768 | 79.8 | 81.19 | +1.393 | -0.242 | 0.809 | 0.022 |
| Bootstrap | 6 | mass | 768 | 768 | 31.99 | 32.54 | +0.548 | -1.349 | 0.178 | 0.059 |
| Bootstrap | 7 | pedi | 768 | 768 | 0.4719 | 0.4697 | -0.002129 | +0.125 | 0.901 | 0.021 |
| Bootstrap | 8 | age | 768 | 768 | 33.24 | 32.96 | -0.2812 | +0.466 | 0.641 | 0.035 |
| Column-wise | 1 | preg | 768 | 768 | 3.845 | 3.759 | -0.08594 | +0.501 | 0.617 | 0.022 |
| Column-wise | 2 | plas | 768 | 768 | 120.9 | 121.1 | +0.1992 | -0.123 | 0.902 | 0.025 |
| Column-wise | 3 | pres | 768 | 768 | 69.11 | 68.53 | -0.5742 | +0.575 | 0.566 | 0.031 |
| Column-wise | 4 | skin | 768 | 768 | 20.54 | 20.56 | +0.02604 | -0.032 | 0.975 | 0.030 |
| Column-wise | 5 | insu | 768 | 768 | 79.8 | 87.54 | +7.74 | -1.281 | 0.2 | 0.035 |
| Column-wise | 6 | mass | 768 | 768 | 31.99 | 31.8 | -0.1951 | +0.490 | 0.624 | 0.025 |
| Column-wise | 7 | pedi | 768 | 768 | 0.4719 | 0.4741 | +0.002174 | -0.130 | 0.896 | 0.033 |
| Column-wise | 8 | age | 768 | 768 | 33.24 | 33.2 | -0.03906 | +0.065 | 0.948 | 0.013 |
| GMM | 1 | preg | 768 | 768 | 3.845 | 3.874 | +0.02862 | -0.166 | 0.868 | 0.154 |
| GMM | 2 | plas | 768 | 768 | 120.9 | 121.2 | +0.3405 | -0.207 | 0.836 | 0.044 |
| GMM | 3 | pres | 768 | 768 | 69.11 | 68.34 | -0.7682 | +0.754 | 0.451 | 0.143 |
| GMM | 4 | skin | 768 | 768 | 20.54 | 21.19 | +0.6544 | -0.819 | 0.413 | 0.188 |
| GMM | 5 | insu | 768 | 768 | 79.8 | 79.32 | -0.4776 | +0.081 | 0.936 | 0.277 |
| GMM | 6 | mass | 768 | 768 | 31.99 | 31.84 | -0.1527 | +0.371 | 0.71 | 0.068 |
| GMM | 7 | pedi | 768 | 768 | 0.4719 | 0.4814 | +0.009475 | -0.564 | 0.573 | 0.132 |
| GMM | 8 | age | 768 | 768 | 33.24 | 33.45 | +0.2067 | -0.339 | 0.735 | 0.155 |
| SMOTE | 1 | preg | 768 | 768 | 3.845 | 3.846 | +0.0005044 | -0.003 | 0.997 | 0.146 |
| SMOTE | 2 | plas | 768 | 768 | 120.9 | 120.1 | -0.7447 | +0.471 | 0.638 | 0.033 |
| SMOTE | 3 | pres | 768 | 768 | 69.11 | 69.99 | +0.8823 | -0.971 | 0.332 | 0.086 |
| SMOTE | 4 | skin | 768 | 768 | 20.54 | 19.82 | -0.7209 | +0.913 | 0.361 | 0.046 |
| SMOTE | 5 | insu | 768 | 768 | 79.8 | 81.09 | +1.29 | -0.221 | 0.825 | 0.018 |
| SMOTE | 6 | mass | 768 | 768 | 31.99 | 31.47 | -0.5215 | +1.384 | 0.167 | 0.078 |
| SMOTE | 7 | pedi | 768 | 768 | 0.4719 | 0.4711 | -0.0007566 | +0.050 | 0.96 | 0.129 |
| SMOTE | 8 | age | 768 | 768 | 33.24 | 33.33 | +0.08629 | -0.149 | 0.882 | 0.094 |
| GMM-SMOTE | 1 | preg | 768 | 768 | 3.845 | 3.926 | +0.08097 | -0.505 | 0.614 | 0.155 |
| GMM-SMOTE | 2 | plas | 768 | 768 | 120.9 | 120.4 | -0.5321 | +0.333 | 0.739 | 0.023 |
| GMM-SMOTE | 3 | pres | 768 | 768 | 69.11 | 68.18 | -0.9304 | +0.940 | 0.348 | 0.074 |
| GMM-SMOTE | 4 | skin | 768 | 768 | 20.54 | 19.73 | -0.8063 | +1.013 | 0.311 | 0.051 |
| GMM-SMOTE | 5 | insu | 768 | 768 | 79.8 | 75.83 | -3.97 | +0.690 | 0.491 | 0.026 |
| GMM-SMOTE | 6 | mass | 768 | 768 | 31.99 | 31.85 | -0.1454 | +0.386 | 0.699 | 0.051 |

| Method | No. | Feature | n real | n synthetic | Real mean | Synthetic mean | Difference | Welch t | p | KS |
| --- | --- | --- | --- | --- | --- | --- | --- | --- | --- | --- |
| GMM-SMOTE | 7 | pedi | 768 | 768 | 0.4719 | 0.4684 | -0.00349 | +0.229 | 0.819 | 0.100 |
| GMM-SMOTE | 8 | age | 768 | 768 | 33.24 | 33.79 | +0.5497 | -0.953 | 0.341 | 0.086 |
| CVAE | 1 | preg | 768 | 768 | 3.845 | 3.561 | -0.2839 | +1.880 | 0.0604 | 0.215 |
| CVAE | 2 | plas | 768 | 768 | 120.9 | 122.5 | +1.57 | -1.039 | 0.299 | 0.081 |
| CVAE | 3 | pres | 768 | 768 | 69.11 | 70.45 | +1.349 | -1.611 | 0.107 | 0.091 |
| CVAE | 4 | skin | 768 | 768 | 20.54 | 20.86 | +0.3221 | -0.424 | 0.672 | 0.237 |
| CVAE | 5 | insu | 768 | 768 | 79.8 | 82.97 | +3.173 | -0.590 | 0.555 | 0.402 |
| CVAE | 6 | mass | 768 | 768 | 31.99 | 32.32 | +0.3248 | -0.857 | 0.392 | 0.048 |
| CVAE | 7 | pedi | 768 | 768 | 0.4719 | 0.4869 | +0.01499 | -0.995 | 0.32 | 0.191 |
| CVAE | 8 | age | 768 | 768 | 33.24 | 32.98 | -0.258 | +0.461 | 0.645 | 0.152 |

## S3 Supplementary GMM Model-Selection Analysis

**Table S5:** AIC- and BIC-based selection of component number for class-specific Gaussian mixture models. Candidate models containing (K=2,. . . ,5) components were fitted separately to each outcome class within each dataset. (n) denotes the class-specific sample size, and lower AIC or BIC values indicate stronger support under the corresponding criterion. AIC (K) and BIC (K) identify the component numbers minimizing each criterion, while “Agree” indicates whether both criteria selected the same value of (K). The criteria agreed only for HIV class 0, for which both selected (K=2).

| Dataset | Class | $n$ | AIC | | | | BIC | | | | AIC $K$ | BIC $K$ | Agree |
| --- | --- | --- | --- | --- | --- | --- | --- | --- | --- | --- | --- | --- | --- |
| | | | $K = 2$ | $K = 3$ | $K = 4$ | $K = 5$ | $K = 2$ | $K = 3$ | $K = 4$ | $K = 5$ | | | |
| Breast Cancer | 0 | 212 | 3278.90 | 3091.85 | 2290.20 | 1205.98 | 6605.28 | 8083.09 | 8946.31 | 9526.96 | 5 | 2 | No |
| Breast Cancer | 1 | 357 | 3460.49 | 2546.79 | 1521.41 | 1418.28 | 7303.32 | 8312.98 | 9210.96 | 11031.19 | 5 | 2 | No |
| Diabetes | 0 | 500 | 8626.79 | 7521.42 | 6962.43 | 6819.60 | 9001.89 | 8086.18 | 7716.84 | 7763.67 | 5 | 4 | No |
| Diabetes | 1 | 268 | 4243.42 | 4140.65 | 3996.91 | 3954.00 | 4563.02 | 4621.85 | 4639.70 | 4758.38 | 5 | 2 | No |
| HIV | 0 | 23 | 603.46 | 4711.78 | 7910.85 | 12246.29 | 5325.98 | 11796.13 | 17357.02 | 24054.29 | 2 | 2 | Yes |
| HIV | 1 | 68 | -2797.14 | -3649.28 | 309.62 | 498.00 | 6433.79 | 10198.23 | 18773.71 | 23578.66 | 3 | 2 | No |

**Figure S3:**
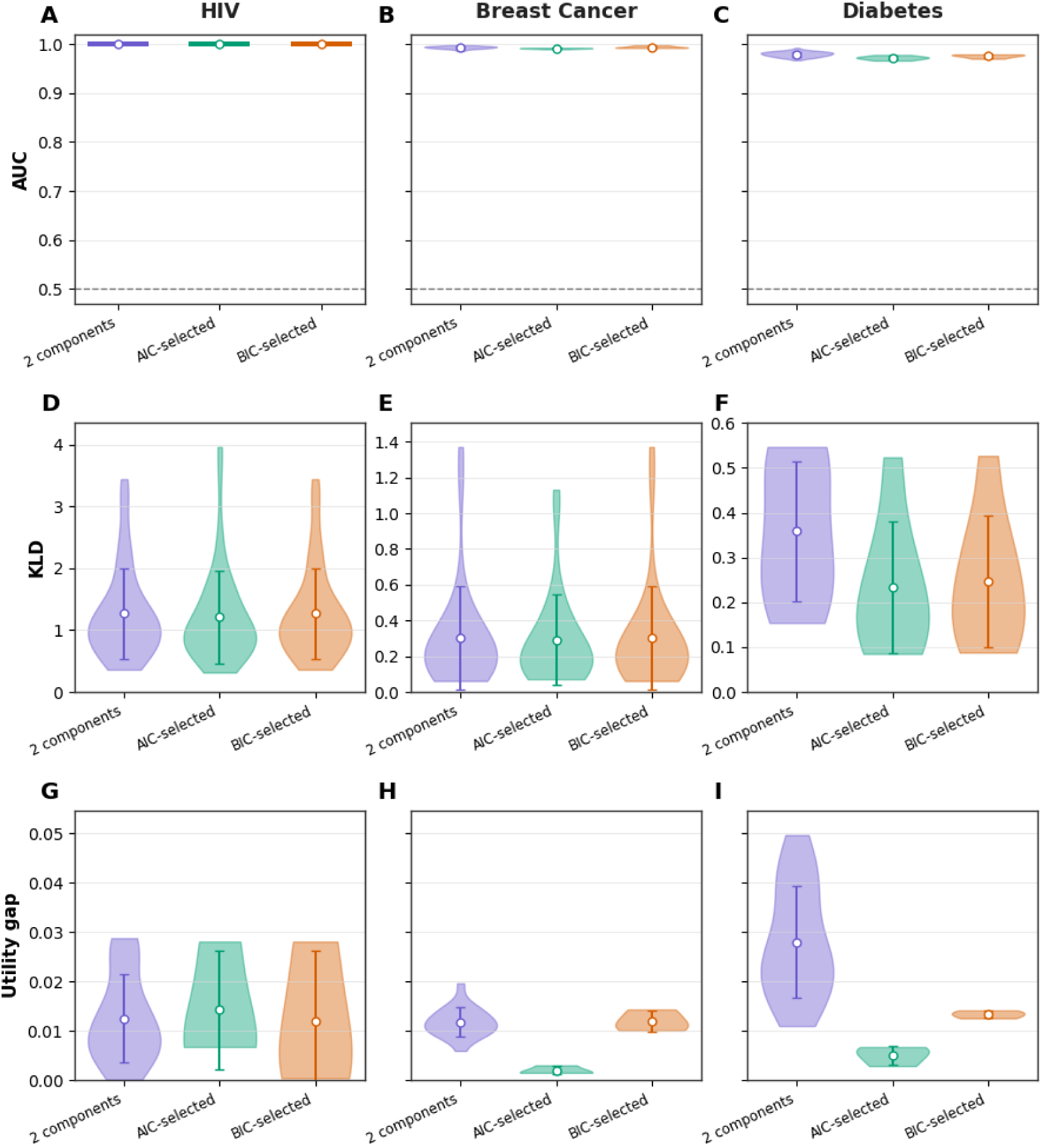
Effect of GMM component-number selection on synthetic-data fidelity. The original models used two components per outcome class, while the alternative models selected the component number separately by class using AIC or BIC. Panels A–C show discriminator AUC, panels D–F show mean feature-level KLD, and panels G–I show the absolute utility gap. Circles indicate means and error bars indicate ±1 standard deviation.

## S4 Multidimensional fidelity results for diabetes and breast cancer data sets

**Figure S4:**
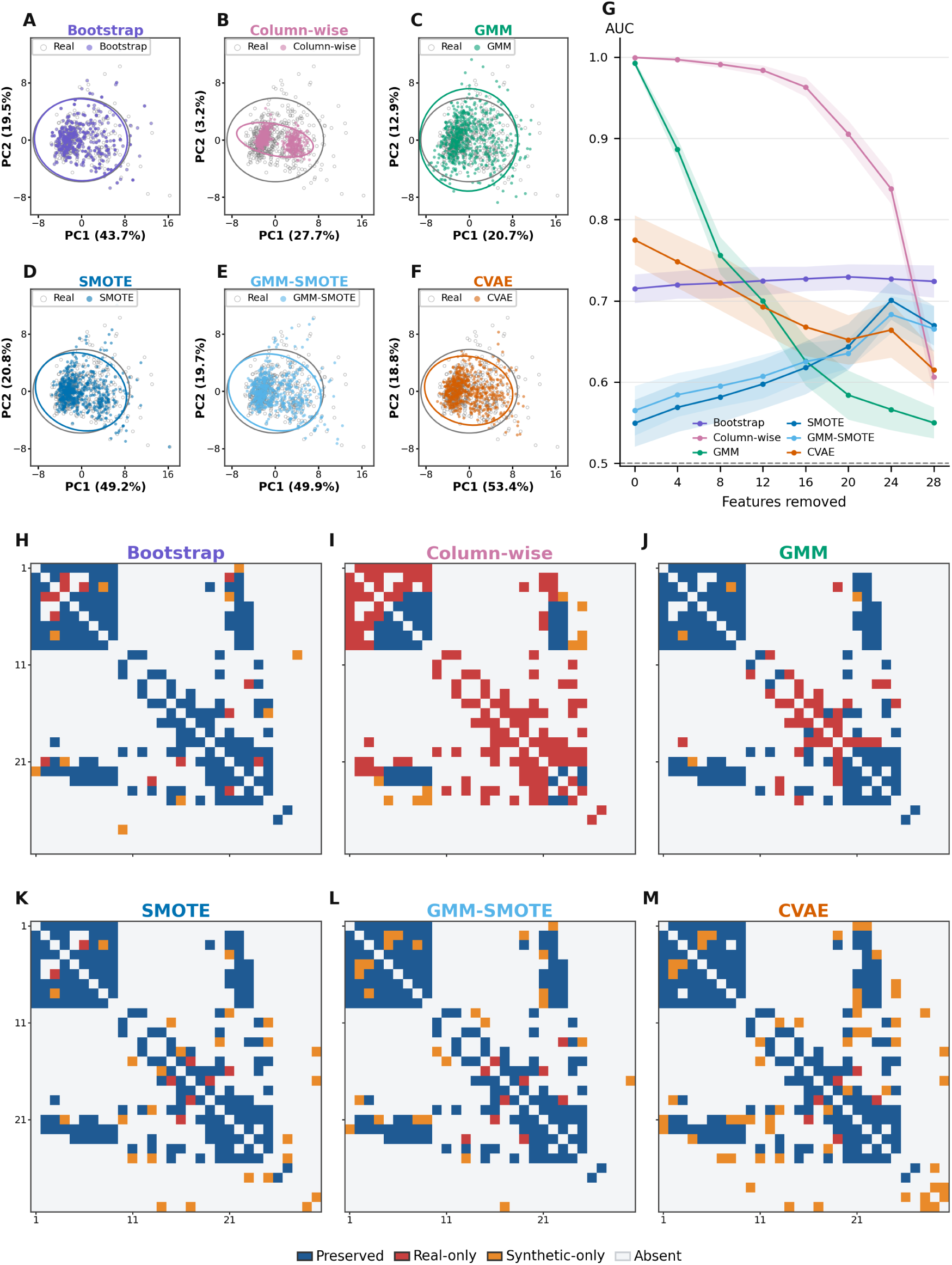
Multidimensional fidelity assessment for the breast cancer dataset. A–F) real and synthetic observations projected onto the first two principal components fitted to the standardized real data, following the same method order as panels H–M. G) real–synthetic discriminator AUC as features were progressively removed in decreasing order of importance; shaded regions indicate variation across repeated train–test splits, and the dashed line denotes chance-level discrimination at AUC (=0.5). H–M) Graphical LASSO conditional-dependence networks for bootstrap, column-wise resampling, GMM, SMOTE, GMM–SMOTE, and CVAE, respectively. Blue cells indicate dependencies preserved between the real and synthetic networks, red cells indicate real-only dependencies lost during generation, orange cells indicate synthetic-only dependencies, and light-gray cells indicate dependencies absent from both networks. The feature ordering was derived from the real-data network and held fixed across methods.

**Figure S5:**
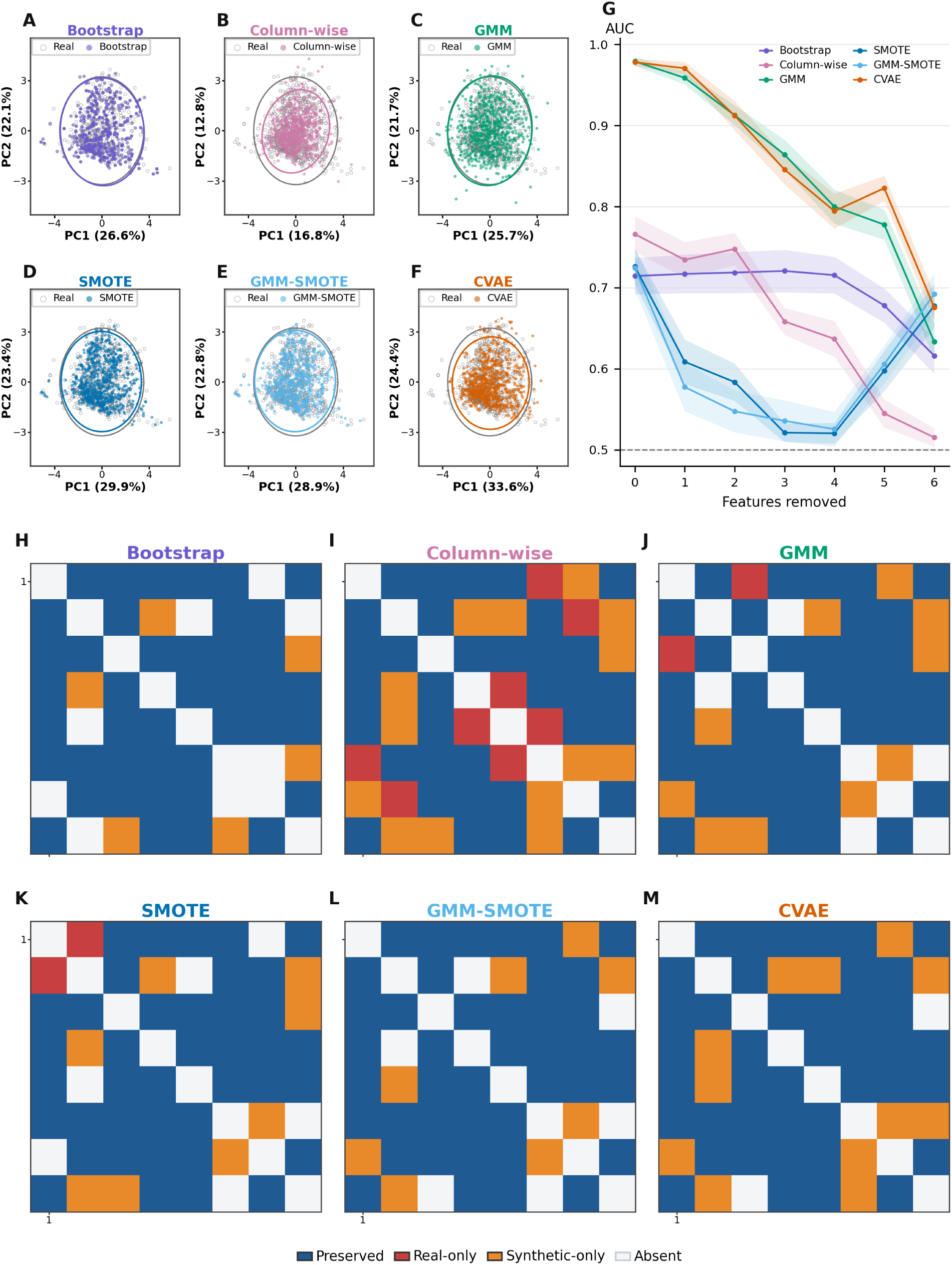
Multidimensional fidelity assessment for the diabetes dataset. A–F) real and synthetic observations projected onto the first two principal components fitted to the standardized real data, following the same method order as panels H–M. G) real–synthetic discriminator AUC as features were progressively removed in decreasing order of importance; shaded regions indicate variation across repeated train–test splits, and the dashed line denotes chance-level discrimination at AUC (=0.5). H–M) Graphical LASSO conditional-dependence networks for bootstrap, column-wise resampling, GMM, SMOTE, GMM–SMOTE, and CVAE, respectively. Blue cells indicate dependencies preserved between the real and synthetic networks, red cells indicate real-only dependencies lost during generation, orange cells indicate synthetic-only dependencies, and light-gray cells indicate dependencies absent from both networks. The feature ordering was derived from the real-data network and held fixed across methods.

## S5 Pairwise correlation comparisons

**Table S6:** Agreement between the Pearson correlation matrices of the synthetic and corresponding real datasets. Frobenius distance is the off-diagonal Frobenius norm, ‖*R_S_* − *R_R_* ‖ *_F_*. The dimension-normalized error, 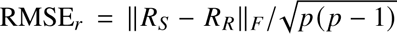, is the root-mean-square error across off-diagonal correlation coefficients and is comparable across datasets. Lower values indicate closer preservation of the real-data pairwise correlation structure. The lowest value within each dataset is shown in bold as a visual aid and does not indicate statistical significance.

| Method | HIV |  | Breast cancer |  | Diabetes |  |
| --- | --- | --- | --- | --- | --- | --- |
|  | Frob. dist. | RMSE<sub>r</sub> | Frob. dist. | RMSE<sub>r</sub> | Frob. dist. | RMSE<sub>r</sub> |
| Bootstrap | **5.234** | **0.0837** | **1.006** | **0.0341** | 0.276 | 0.0369 |
| Column-wise | 22.651 | 0.3624 | 8.148 | 0.2762 | 1.368 | 0.1828 |
| GMM | 6.653 | 0.1065 | 3.610 | 0.1224 | **0.275** | **0.0367** |
| SMOTE | 8.181 | 0.1309 | 1.925 | 0.0653 | 0.378 | 0.0505 |
| GMM-SMOTE | 6.646 | 0.1063 | 1.612 | 0.0546 | 0.315 | 0.0421 |
| CVAE | 12.636 | 0.2022 | 2.758 | 0.0935 | 0.756 | 0.1010 |

**Figure S6:**
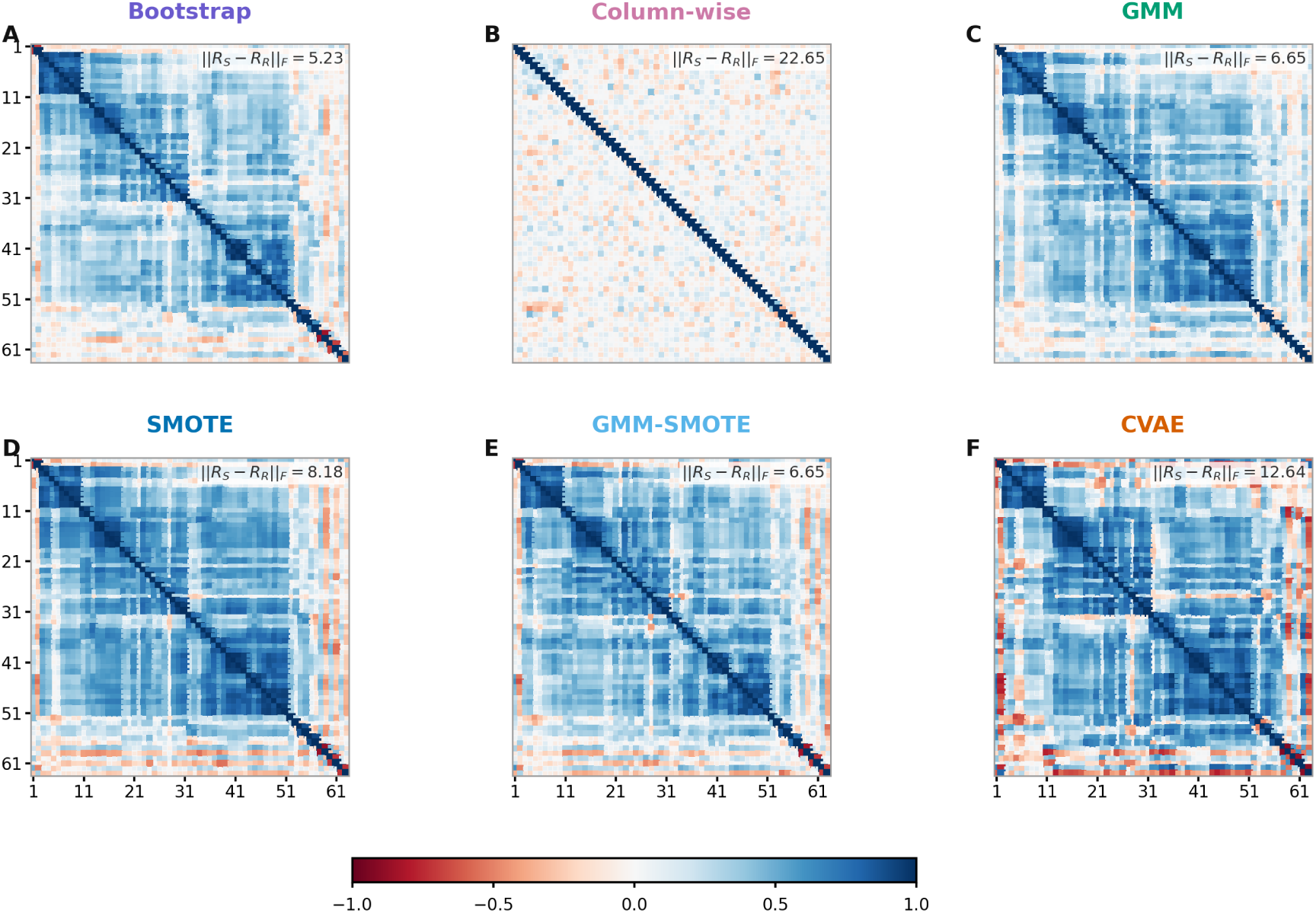
Pairwise Pearson correlation comparison for the HIV dataset. Panels A–F show the correlation matrices for bootstrap, column-wise resampling, GMM, SMOTE, GMM–SMOTE, and CVAE, respectively. Values shown in each panel report the Frobenius distance between the synthetic and real correlation matrices, ‖*R_S_* − *R_R_*‖ *_F_*, with smaller values indicating closer agreement.

**Figure S7:**
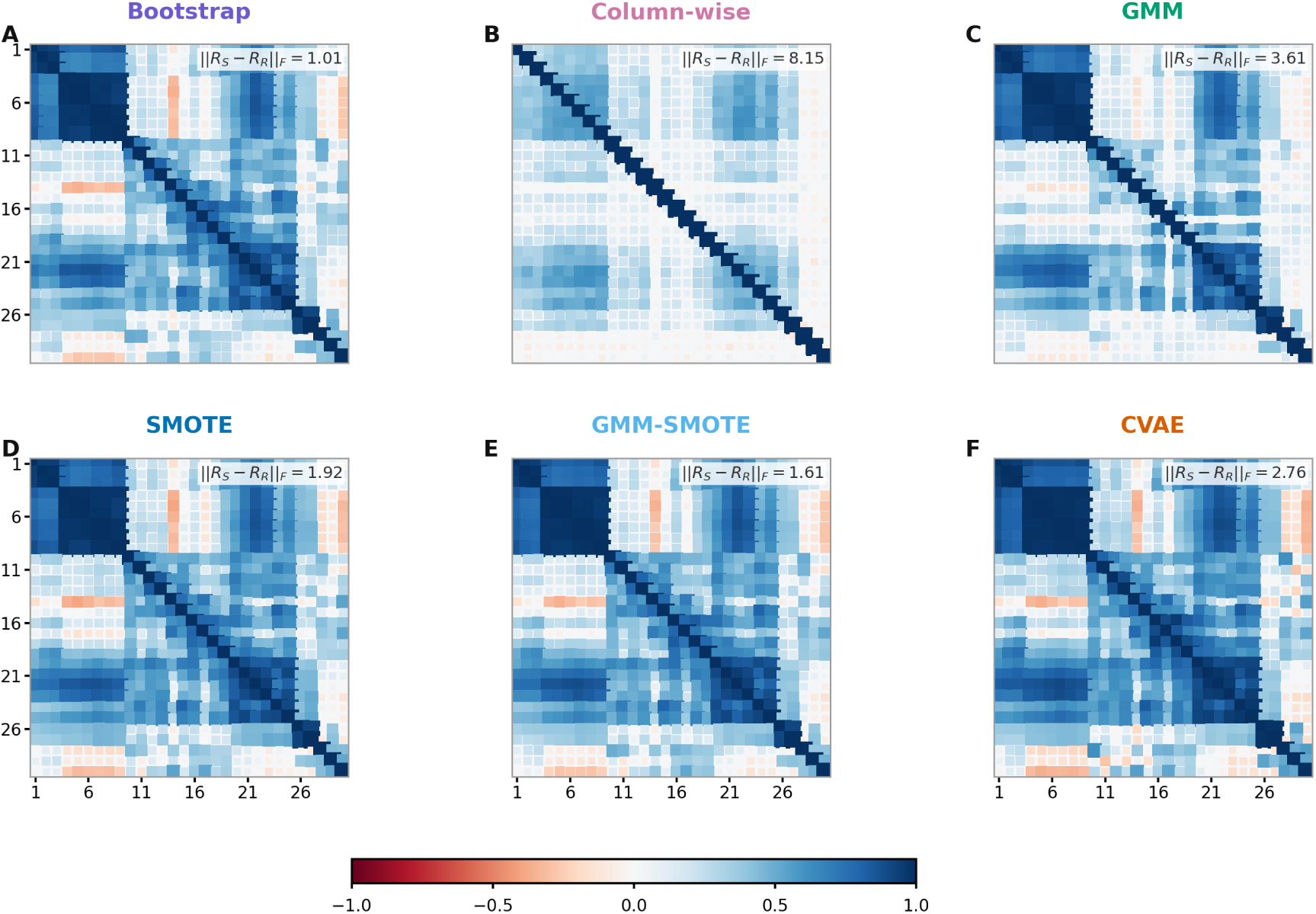
Pairwise Pearson correlation comparison for the breast cancer dataset. Panels A–F show the correlation matrices for bootstrap, column-wise resampling, GMM, SMOTE, GMM–SMOTE, and CVAE, respectively. Values shown in each panel report the Frobenius distance between the synthetic and real correlation matrices, ‖*R_S_*−*R_R_*‖*_F_*, with smaller values indicating closer agreement.

**Figure S8:**
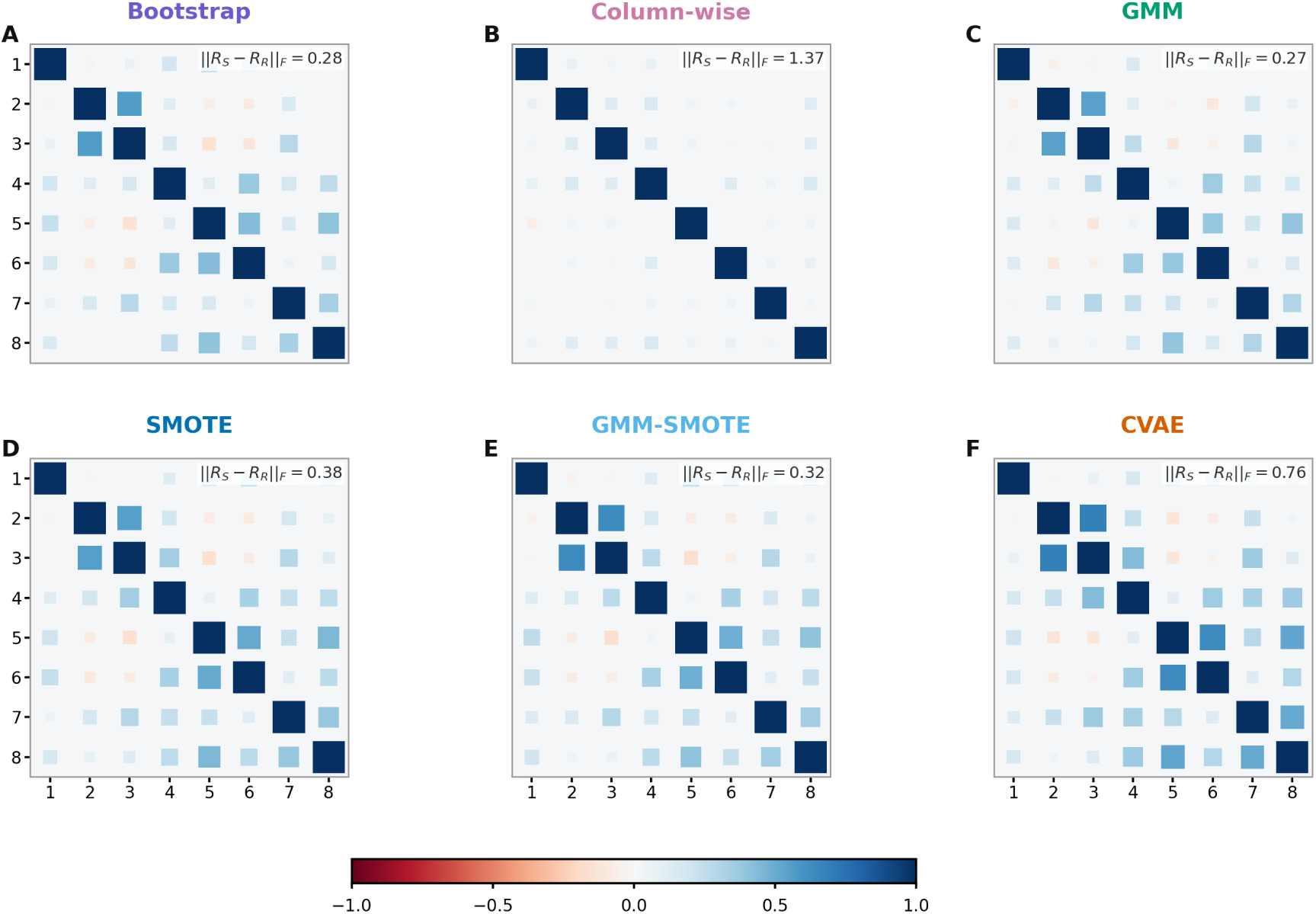
Pairwise Pearson correlation comparison for the diabetes dataset. Panels A–F show the correlation matrices for bootstrap, column-wise resampling, GMM, SMOTE, GMM–SMOTE, and CVAE, respectively. Values shown in each panel report the Frobenius distance between the synthetic and real correlation matrices, ‖*R_S_* − *R_R_*‖ *_F_*, with smaller values indicating closer agreement.

## S6 Supplementary Graphical Lasso Analyses & Within-class Permutation

**Figure S9:**
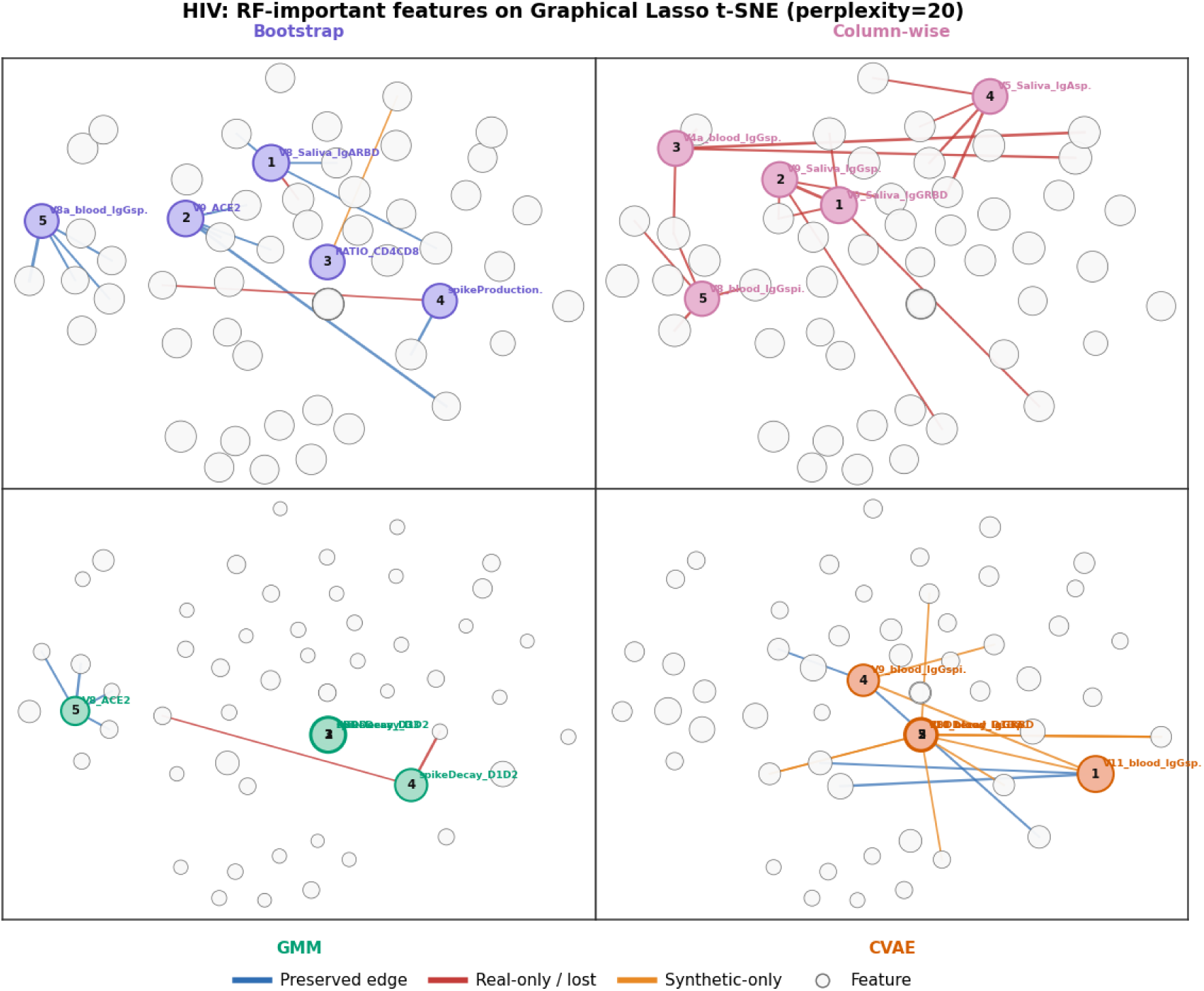
Exploratory overlay of discriminator-important HIV features on Graphical Lasso t-SNE layouts. Features are arranged using two-dimensional t-SNE representations of the Graphical Lasso networks for bootstrap, column-wise resampling, GMM, and CVAE (perplexity = 20). The five features contributing most strongly to the corresponding real–synthetic random-forest discriminator are highlighted and numbered by importance rank; all remaining features are shown as open gray circles. Edges are colored blue when preserved in the real and synthetic networks, red when present only in the real network (lost), and orange when present only in the synthetic network. The t-SNE layouts are exploratory and did not reveal a consistent spatial organization of discriminator-important features across methods; distances and apparent clusters should therefore not be interpreted quantitatively.

### S6.1 Regularization path example

**Figure S10:**
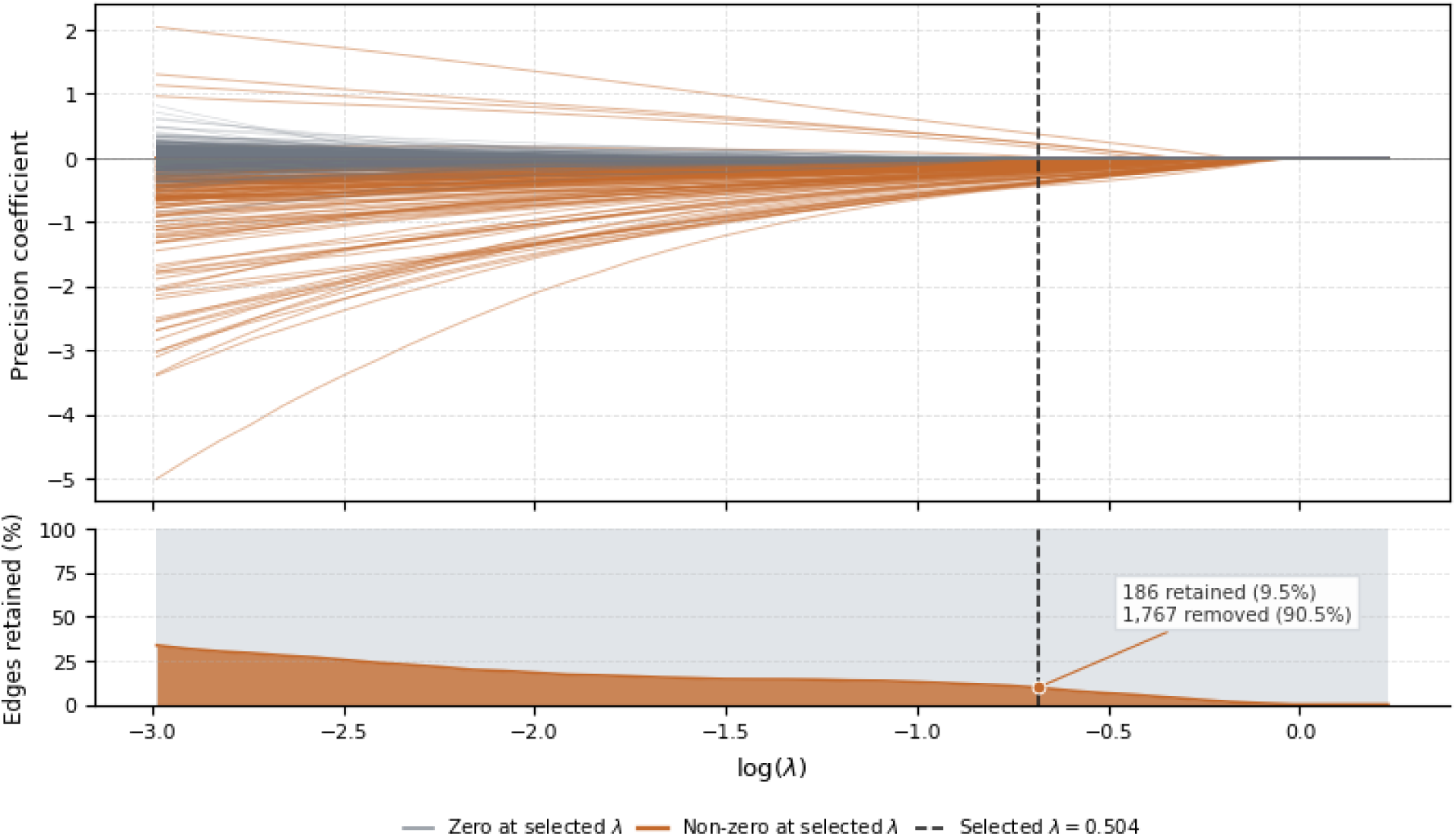
Representative Graphical Lasso regularization path for the HIV dataset. The upper panel shows the regularization paths of the 1,953 possible unique off-diagonal precision-matrix coefficients among the 63 features as a function of log(*λ*). The dashed vertical line marks the cross-validation-selected penalty, *λ* = 0.504; coefficients remaining nonzero at *λ* are shown in orange, whereas coefficients that had shrunk to zero are shown in gray. The lower panel summarizes the percentage of possible edges retained across the regularization path. At *λ*^∗^, 186 edges remained nonzero (9.5%) and 1,767 were set to zero (90.5%), illustrating the sparsity imposed in the HIV real-data network.

### S6.2 Feature ordering

**Table S7:**
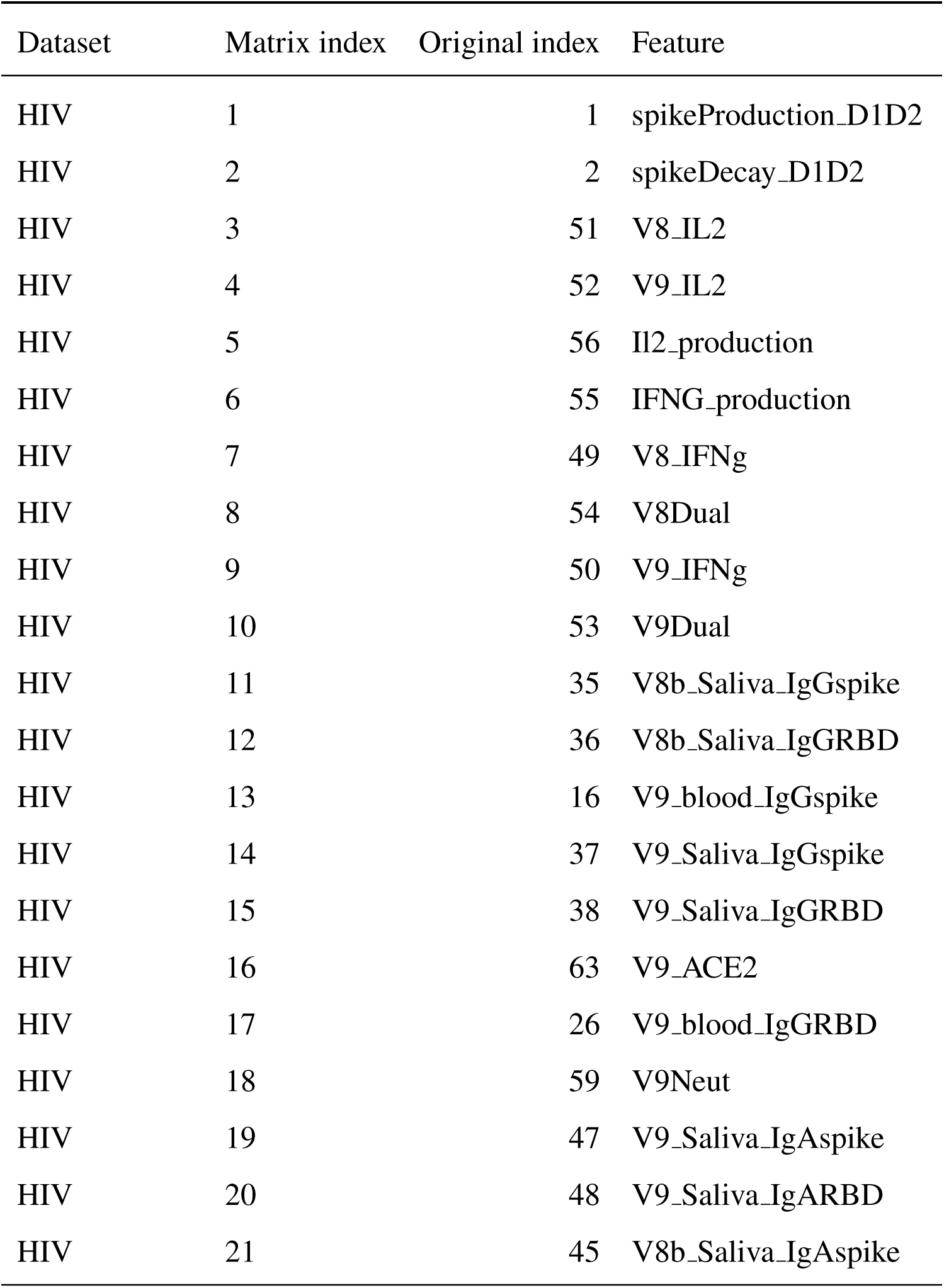
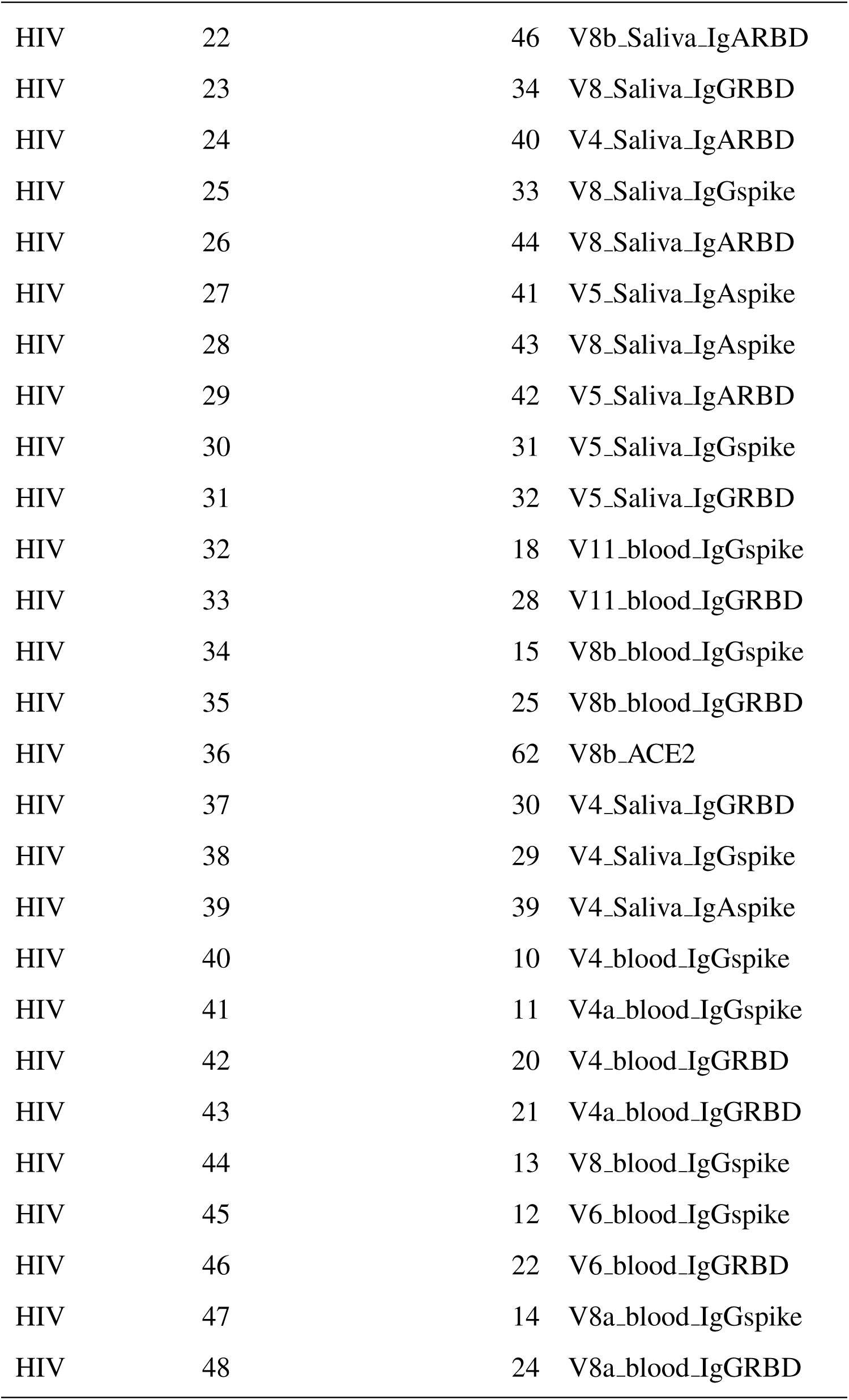
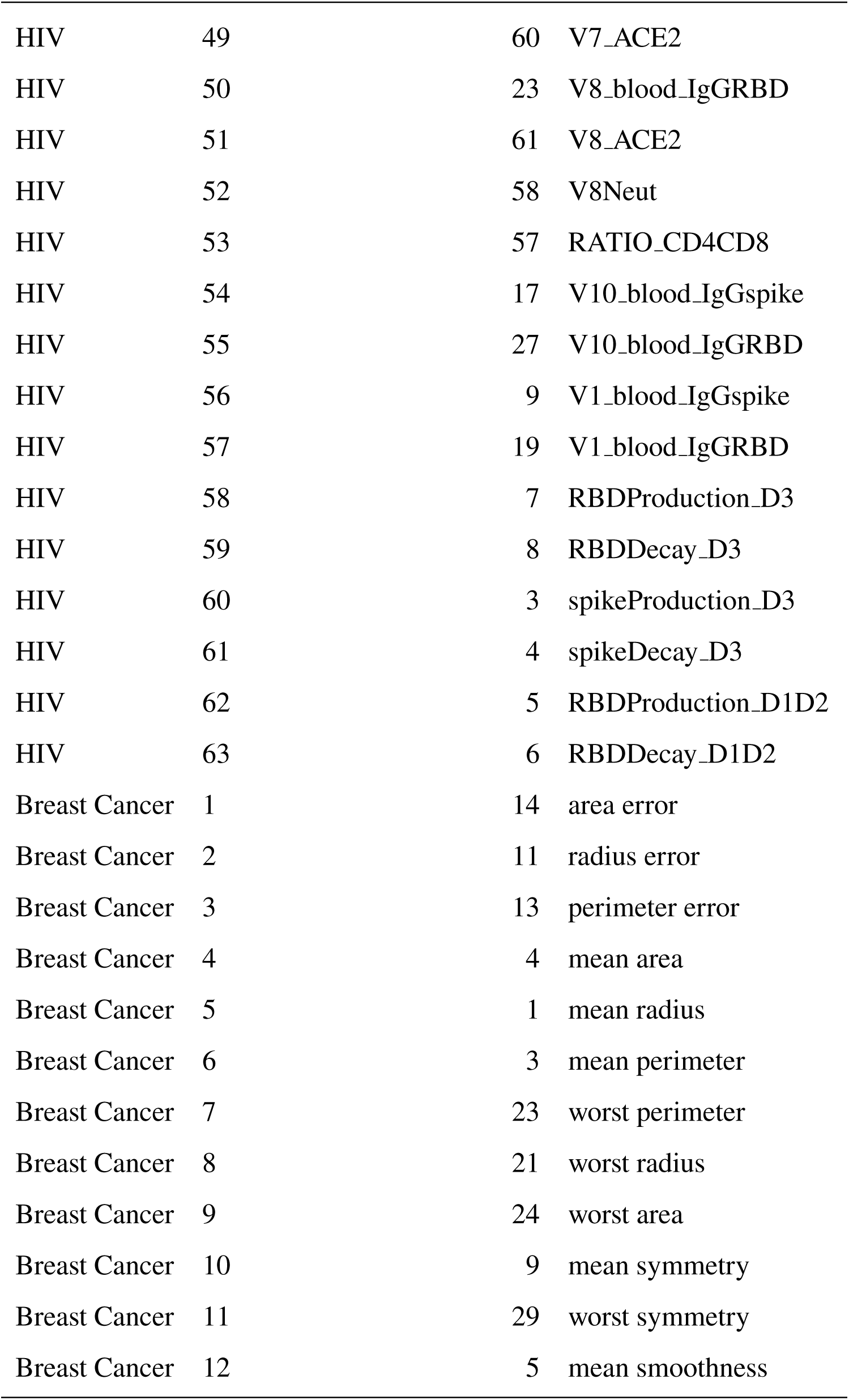
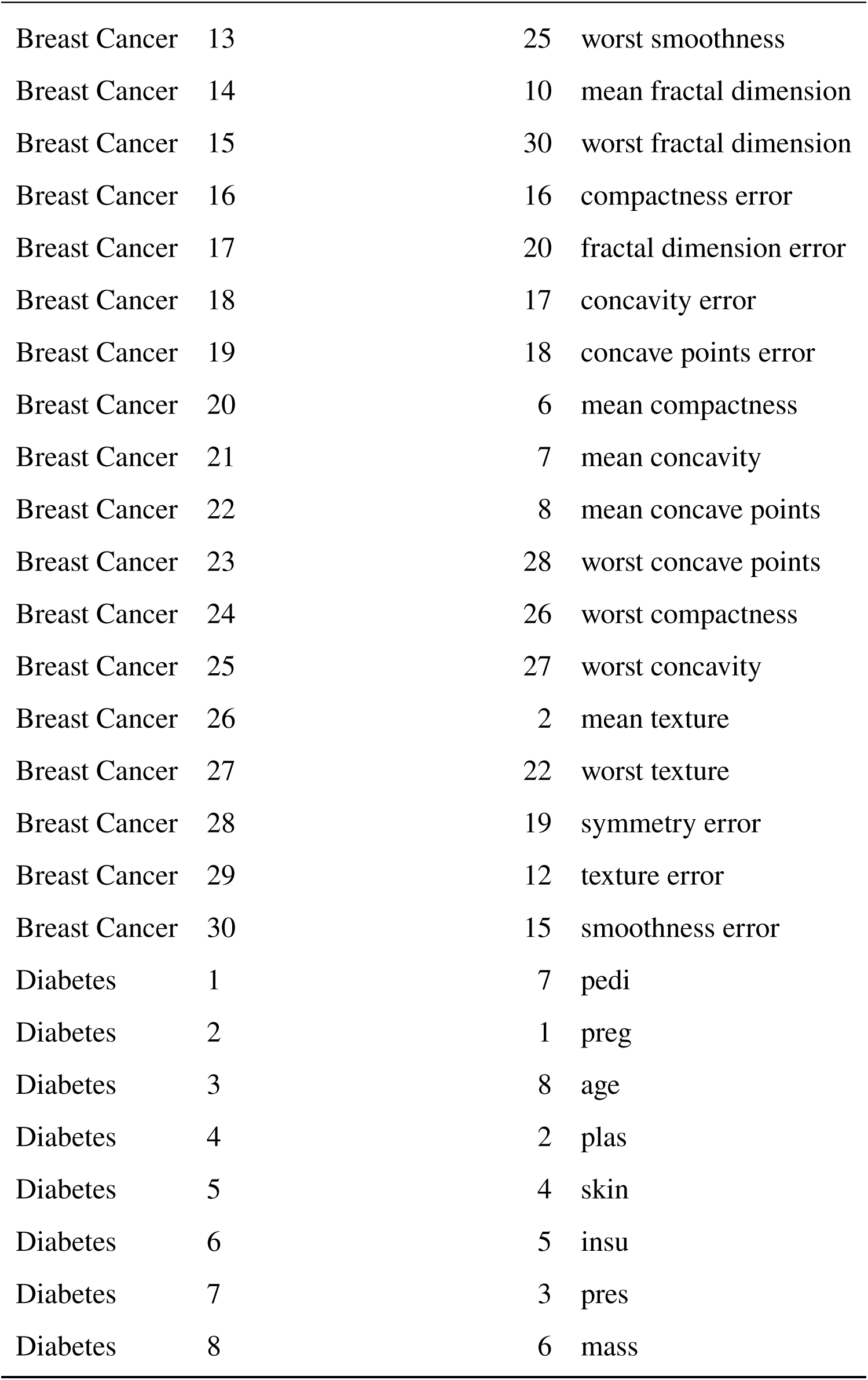
Feature ordering used in the Graphical Lasso edge-comparison matrices. For each dataset, matrix positions were obtained by average-linkage hierarchical clustering of the absolute partial-correlation structure estimated from the real data, using dissimilarity 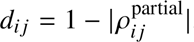. The same order was applied to every synthetic method.

### S6.3 Within-class permutation analysis

**Figure S11:**
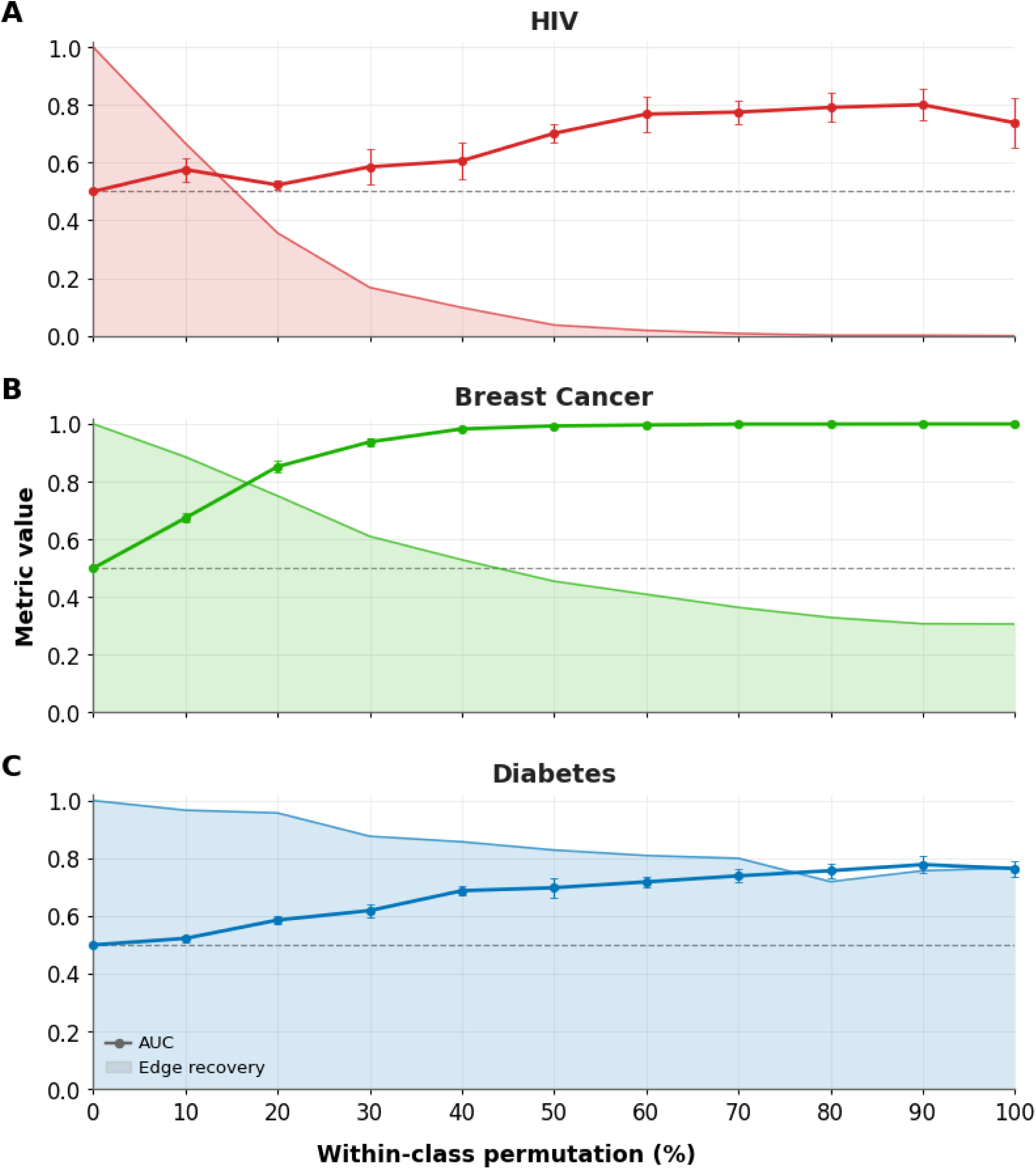
Figure

## Notes

### Competing Interest Statement

The authors have declared no competing interest.

